# Brain-heart-eye axis revealed by multi-organ imaging, genetics and proteomics

**DOI:** 10.1101/2025.01.04.25319995

**Authors:** The MULTI consortium, Aleix Boquet-Pujadas, Filippos Anagnostakis, Michael R. Duggan, Cassandra M. Joynes, Arthur W. Toga, Zhijian Yang, Keenan A. Walker, Christos Davatzikos, Junhao Wen

## Abstract

Multi-organ research investigates interconnections among multiple human organ systems, enhancing our understanding of human aging and disease mechanisms. Here, we used multi-organ imaging (*N*=105,433), individual- and summary-level genetics, and proteomics (*N*=53,940) from the UK Biobank, Baltimore Longitudinal Study of Aging, FinnGen, and Psychiatric Genomics Consortium to delineate a brain-heart-eye axis via 2003 brain patterns of structural covariance^1^ (PSC), 82 heart imaging-derived phenotypes^2^ (IDP) and 84 eye IDPs^3–5^. Cross-organ phenotypic associations highlight the central autonomic network between the brain and heart and the central visual pathway between the brain and eye. Proteome-wide associations of the PSCs and IDPs show both within-organ specificity and cross-organ interconnections, verified by the RNA and protein expression profiles of the 2923 plasma proteins. Pleiotropic effects of common genetic variants are observed across multiple organs, and key genetic parameters, such as SNP-based heritability, polygenicity, and selection signatures, are comparatively evaluated among the three organs. A gene-drug-disease network shows the potential of drug repurposing for cross-organ diseases. Colocalization and causal analyses reveal cross-organ causal relationships between PSC/IDP and chronic diseases, such as Alzheimer’s disease, heart failure, and glaucoma. Finally, integrating multi-organ/omics features improves prediction for systemic disease categories and cognition compared to single-organ/omics features. This study depicts a detailed brain-heart-eye axis and highlights future avenues for modeling human aging and disease across multiple scales. All results are publicly available at https://labs-laboratory.com/medicine/.

## Main

The brain, heart, and eye are key organs of human physiology, collectively orchestrating intricate and vital processes essential for humans^6,7^. Recent multi-organ research endeavors^8,9,10,11,12,13,14,15,16,17^ have unveiled complex and synergistical relationships between different human organ systems. These approaches hold the potential to unveil cross-organ connections, elucidate the complex tapestry of human health and disease, and potentially pave the road toward precision medicine^18^.

Imaging genetics^19^ integrates *in vivo* imaging data like magnetic resonance imaging (MRI) with genetics, shedding light on disease mechanisms along underlying causal pathways^20^ spanning from genetics to imaging-derived phenotypes (IDPs) and disease manifestations.

Additional omics data like transcriptomics, proteomics, and metabolomics^21^ can further enrich our understanding of this causal pathway. The burgeoning development of machine learning has led researchers to apply this technology to analyze brain MRI, yielding insights into brain aging and disease. These include diagnosing and predicting diseases such as Alzheimer’s disease (AD)^22^, evaluating personalized treatment effectiveness like multiple sclerosis^23^, and stratifying patient populations into pathologically homogeneous subgroups^24^, as well as in typical aging^25^. Both conventional and machine learning-derived brain imaging-derived phenotypes (IDPs) were used in genome-wide association studies (GWAS)^26–29^, which were correlated with common and rare single-nucleotide polymorphisms (SNPs). Likewise, cardiac MRI is pivotal in preventing, diagnosing, and treating cardiovascular conditions related to the heart, such as ischaemic heart disease^30^. For example, recent studies^2,31–35^ have analyzed large-scale heart MRI data using machine learning to extract heart IDPs to quantify cardiac structure and function. Heart imaging GWAS was also performed to delineate the genetic architecture of these heart IDPs^36–38^. For example, Aung and colleagues performed GWAS to identify 25 genomic loci associated with the right ventricular structure and function of the heart^37^. Finally, when integrated with machine learning, optical coherence tomography (OCT) imaging has revolutionized diagnosing and managing various eye diseases. For example, Zhou et al.^39^ recently introduced RETFound, a foundational model designed for retinal images. GWAS on eye OCT IDPs have also revealed multiple genetic loci associated with the structure of the eye^40,41^.

Mutli-organ research has revealed cross-organ connections^8,9,10,11,12,13,14,15^, by allowing researchers to delve into human aging and disease with a granularity that surpasses the single-organ lens, thus better capturing the multifaceted disease etiology. A driving force behind this was the advance of large-scale data consolidation initiatives, notably the UK Biobank (UKBB)^42^, which offered the scientific community multi-organ imaging and multi-omics data. In UKBB, this collection of multi-organ imaging data encompasses brain MRI, heart MRI, eye OCT, abdominal MRI (such as liver and pancreas scans), and whole-body dual-energy X-ray absorptiometry imaging (Category ID:100003). For example, McCracken et al.^11^ used multi-organ imaging from UKBB to showcase the interrelationship between the heart, brain, and liver. Zhao et al.^8^, in a recent study, reinforced the heart-brain connection by integrating genetic data, unveiling insights into its genetic foundation. Yet, the brain IDPs utilized were predefined based on the brain’s neuroanatomy, encompassing volumetric assessments derived from specific regions of interest (ROI) in T1-weighted MRI or diffusivity metrics related to white matter tracts obtained from diffusion MRI. Machine learning has been integrated into imaging genetics and has shown great potential to provide additional power in identifying new genomic loci^1,43^. For instance, Patel et al.^43^ utilized a 3D convolutional autoencoder for training on T1-weighted and T2-FLAIR brain MRIs from 6,130 UKBB participants, generating a 128-dimensional representation termed ENDOs and showing additional discovery power compared to previous predefined atlas-based brain IDPs. Similarly, in a prior study^1^, we introduced sopNMF, a stochastic optimization method for non-negative matrix factorization. This approach parcellates the human brain into data-driven structural networks called patterns of structure covariance^44^ (PSCs). Biologically, PSCs signify coordinated alterations across brain areas and individuals in a data-driven approach – areas like Broca’s cortex and Wernicke’s area might collectively impact individuals with motor speech dysfunction.

This study used individual-level imaging, genetic, and proteomic data from UKBB^42^, Baltimore Longitudinal Study of Aging (BLSA^45^), and GWAS summary statistics from FinnGen^46^ and the Psychiatric Genomics Consortium^47^ (PGC) to delineate the brain-heart-eye axis (**Method 1**). To depict the neuroanatomical structures of the three organs, we used 2003 brain PSCs obtained from 39,567 brain T1-weighted MRI scans in our previous study^1^, 82 heart IDPs from 39,676 heart MRI scans analyzed by Bai et al.^2^, and 84 eye IDPs from 64,317 eye OCT images^3–5^ from UKBB (**Method 2** and **Supplementary Table 1**). We performed a phenome-wide association study (PWAS, **Method 3**) to establish cross-organ phenotypic landscapes between the 2003 brain PSCs, 82 heart IDPs, and 84 eye IDPs. A proteome-wide association study (ProWAS, **Method 4**) was conducted to link the PSCs/IDPs to 2923 plasma proteins (Olink) from UKBB and generate their expression profiles using organ/tissue-specific RNA and protein data^48^; the significant signals were scrutinized in an independent imaging proteomics data (SomaScan) from BLSA. We then conducted GWASs to link the brain PSCs, heart, and eye IDPs with common SNPs. Subsequently, we performed several post-GWAS analyses to partially validate the genetic signals, including estimating key genetic parameters (i.e., SNP-based heritability, polygenicity, and selection signature). A gene-drug-disease network, genetic correlation, colocalization, and causal links between the brain PSCs, heart IDPs, eye IDPS, and the respective diseases of each organ, were also established (**Method 5**).

Lastly, we evaluated the predictive capacity of the brain PSCs, heart and eye IDPs, as well as their corresponding polygenic risk score (PRS), for 14 systemic disease categories and 8 cognitive scores (**Method 6**). Our analytic framework is illustrated in **Fig. 1**.

**Figure 1:**
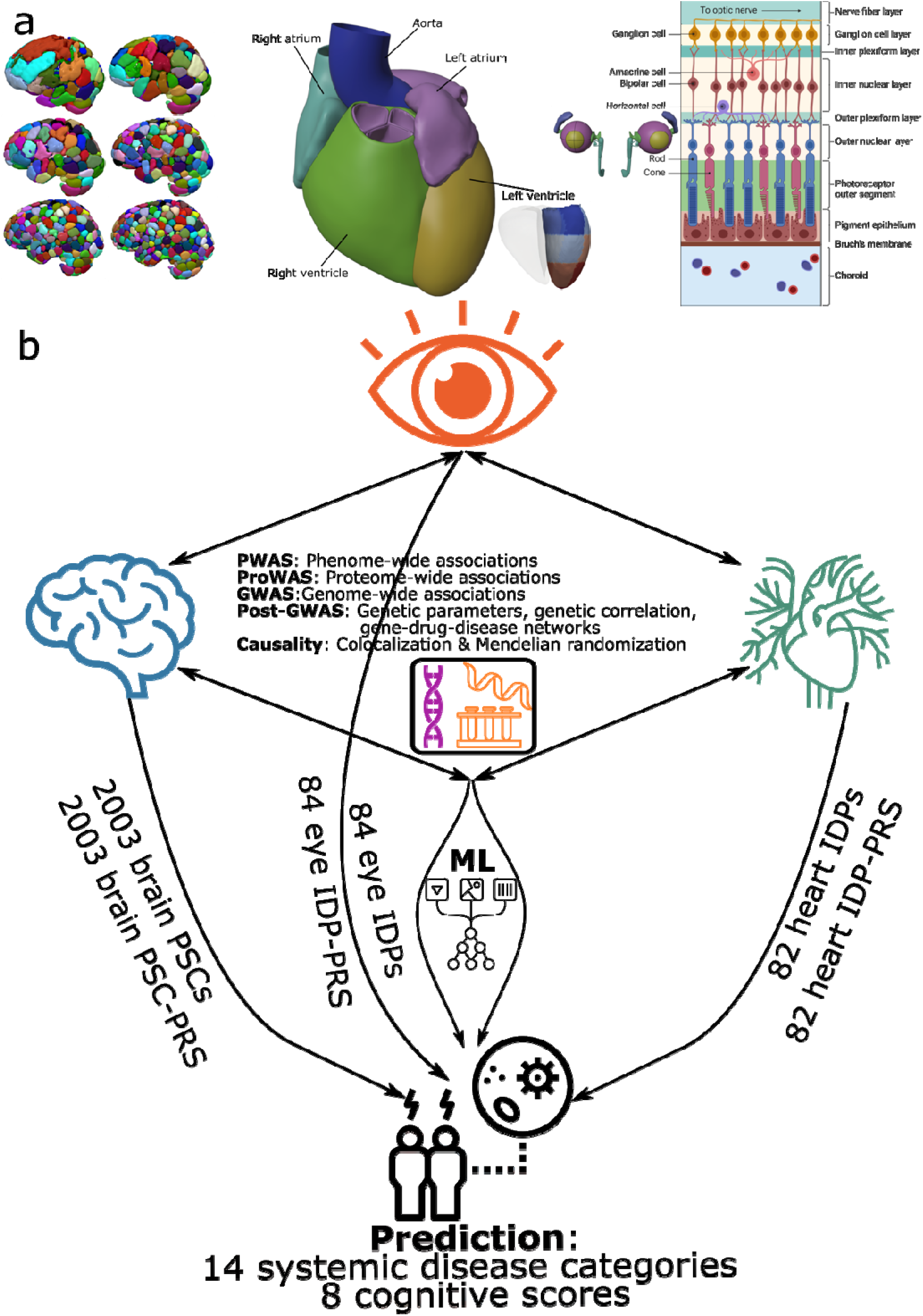
Study workflow **a**) The 2003 multi-scale patterns of structural covariance (PSCs), 82 heart imaging-derived phenotypes (IDPs) from magnetic resonance imaging (MRI), and 84 eye IDPs from optical coherence tomography (OCT). **b**) Three main sets of analyses were conducted to demonstrate the brain-heart-eye axis: *i*) phenotype-wide associations (PWAS) were performed between the 2003 brain PSCs, 82 heart IDPs, and 84 eye IDPs; *ii*) genome-wide association studies (GWAS) and post-GWAS analyses were conducted between the brain PSCs, heart IDPs, eye IDPs, and common genetic variants (SNPs), and *iii*) prediction ability of the brain PSCs, heart IDPs, eye IDPs, and their PRS were assessed with AI to predict 14 systemic disease categories and 8 cognitive scores. All analyses used the Genome Reference Consortium Human Build 37 (GRCh37). Readers can visualize the 2003 brain PSCs via the BRIDGEPORT knowledge portal: https://labs-laboratory.com//bridgeport. Detailed interpretations of the 84 eye IDPs and 82 heart IDPs are publicly available at https://labs-laboratory.com/medicine/eye and https://labs-laboratory.com/medicine/cardiovascular.

## Results

### Phenotypic landscape of the 2003 brain PSCs, 82 heart IDPs, and 84 eye IDPs

To demonstrate the cross-organ phenotypic associations of the brain-heart-eye axis, we conducted three primary PWASs linking the pair-wise imaging features of the three organs.

For the brain-heart PWAS, we found 16,158 significant associations (P-value<0.05/2003/82; effective sample size: 21,948<*N*<23,548; −0.23<*r*<0.52) after applying the Bonferroni correction (**Fig. 2a** and **Method 3a**). The significant brain PSCs largely encompassed deep subcortical structures, the bilateral anterior temporal pole, and the prefrontal cortex. For example, C32_1 (visualization example: https://labs-laboratory.com/bridgeport/music/C32_1), C64_1, and C128_1 delineated subcortical structures, such as the bilateral thalamus. Brain PSCs like C32_3 exemplified the involvement of the bilateral anterior temporal pole. Brain PSC (C128_2: https://labs-laboratory.com/bridgeport/music/C128_2) of the anterior insula was significantly associated with several heart IDPs, such as the myocardial mass of the left ventricle [LVM, *r*=0.27, −log_10_(P-value)=26.62]. The observed brain imaging patterns largely align with the central autonomic network^49^, comprising the prefrontal cortex, amygdala, insular cortex, anterior cingulate cortex, and brainstem. For the 82 heart IDPs, we identified 1313 significant associations with the LVM (9%), 87 significant associations with the right ventricular end-diastolic volume (RVEDV, 8%), and 55 associations with left ventricular mean myocardial wall global thickness (WT_global, 7%). The detailed statistics of the brain-heart PWAS are presented in **Supplementary eFile 1**. We conducted two sensitivity check analyses (**Supplementary eText 1a, eFile 2-3**) to validate the main PWAS results (**Method 3**). We obtained moderate concordance rates (CR) regarding P-values in split-sample (0.72< CR-P <0.99) and sex-stratified (0.38< CR-P <0.91) analyses. Moreover, the β values were highly correlated between the two random splits (*r-β*=0.97) and female vs. male (*r-β*=0.95) PWASs. The brain-heart PWAS results highlighted subcortical structures, the temporal pole, the insula cortex, the prefrontal cortex of the brain, and the left ventricular mass of the heart.

**Figure 2:**
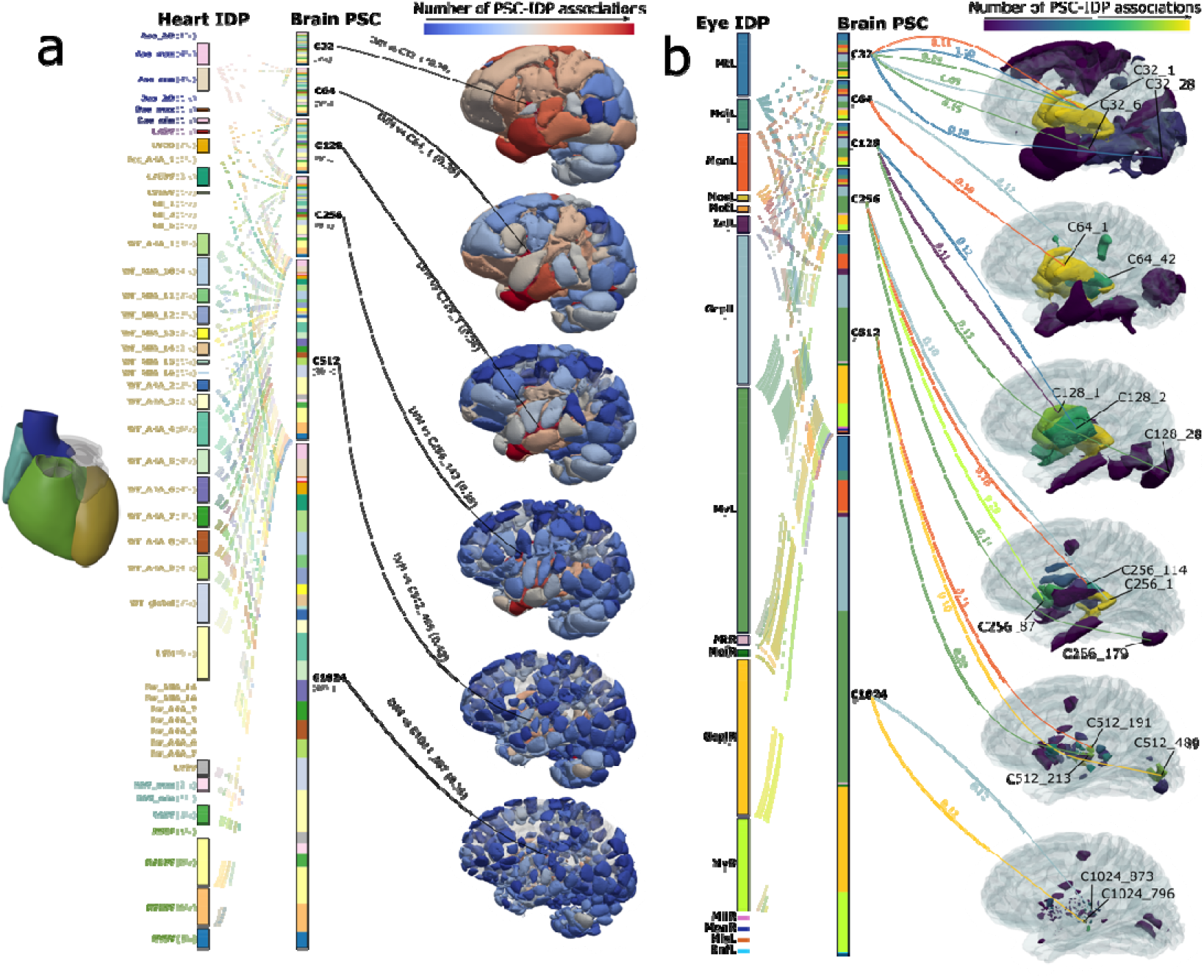
Phenotypic associations of the 2003 brain PSCs, 82 heart IDPs, and 84 eye IDPs. **a**) The phenotypic associations between the 82 heart IDPs (left panel) and 2003 brain PSCs (middle panel) are shown after the Bonferroni correction (P-value<0.05/2003/82; effective sample size: 21,948<*N*<23,548; −0.23<*r*<0.52). For each PSC/IDP category, the percentage of the significant associations is displayed. In the right panel, we charted the significant PSC-IDP associations in the 3D brain space, providing the Pearson correlation coefficient (*r*) for representative brain PSCs with the heart IDP (LVM: left ventricle myocardial mass). **b**) The phenotypic associations between the 2003 brain PSCs (middle panel) and 84 eye IDPs (left panel) are shown. Significant results that survived the Bonferroni correction are presented (P-value<0.05/2003/84; effective sample size: 1284<*N*<4472; −0.09<*r*<0.24). We showed the high-level PSC/IDP categories for visualization purposes. Abbreviations: MtL: overall macular thickness (left); MoiL: macular thickness at the outer inferior subfield (left); MonL: macular thickness at the outer nasal subfield (left); MosL: macular thickness at the outer superior subfield (left); InlL: the average thickness of the inner nuclear layer (left); GcplL: the average thickness of ganglion cells in the inner plexiform layer (left); MvL: total macular volume (left); MR: overall macular thickness (right); GcplR: the average thickness of ganglion cells in the inner plexiform layer (right); MvR: total macular volume (right); InlR: the average thickness of the inner nuclear layer (right); RnfL: the average thickness of the retinal nerve fiber layer. MiiR: macular thickness at the inner inferior subfield (right); MonL: macular thickness at the outer nasal subfield (right); MisL: macular thickness at the inner superior subfield (left). For the phenotypic association between the 82 heart IDPs and 84 eye IDPs, the results did not survive the Bonferroni correction (P-value<0.05/84/82; effective sample size: 1002<*N*<3977; −0.11<*r*<0.15). Readers can visualize the 2003 brain PSCs via the BRIDGEPORT knowledge portal: https://labs-laboratory.com//bridgeport. Detailed interpretations of the 84 eye IDPs and 82 heart IDPs are publicly available at https://labs-laboratory.com/medicine/eye and https://labs-laboratory.com/medicine/cardiovascular.

For the brain-eye PWAS, We discovered 469 significant associations after applying the Bonferroni correction (P-value<0.05/2003/84; effective sample size: 1284<*N*<4472; −0.09<*r*<0.24) (**Fig. 2b**). The prominent brain PSCs encompassed deep subcortical structures (e.g., the thalamus) and the occipital cortex. For example, C32_1, C64_1, and C128_1 delineated subcortical areas, including the lateral geniculate nucleus (LGN) and the hypothalamus. These regions are crucial in the central projection of the retina, playing an essential role in normal visual processing. Specifically, the LGN in the thalamus is a pivotal element of the mammalian visual pathway, establishing connections with the optic nerve/fibers^50^. Moreover, brain PSCs like C32_28, C128_28, C256_179, and C512_489 exemplified primary cerebral cortex involvement in the occipital lobe – the well-known visual cortex regions. For the 84 eye IDPs, we pinpointed 137 significant links with the left total macular volume (MvL, 29%), 87 and 83 significant associations with the left/right average thickness of ganglion cells in the inner plexiform layer (GcplR, 19%; GcplL, 18%), and 55 associations with the right total macular volume (MvR, 11%). Retinal ganglion cells, the projection neurons of the vertebrate retina, are pivotal in conveying essential information from other retinal neurons to various brain regions within the visual pathway^51^. In two sensitivity check analyses, we obtained moderate CRs regarding P-values in split-sample (CR-P=0.67) and sex-stratified (CR-P=0.78) analyses, potentially due to the limited sample sizes (613<*N*<2348). This was corroborated by the high correlation between the β values of the PWASs across different splits (*r-β*=0.99) or sexes (*r-β*=0.97). Detailed results for sensitivity check analyses can be found in **Supplementary eText 1b** and **eFile 4-6**. The brain-eye PWAS revealed a distinct brain-eye phenotypic landscape that aligns closely with the central visual pathway^52^.

For the heart-eye PWAS, the results did not survive the Bonferroni correction (P-value<0.05/84/82; effective sample size: 1002<*N*<3926; −0.11<*r*<0.17). The most prominent association was achieved between the RV stroke volume (RVSV) and macular thickness at the right inner temporal subfield (MitR) (*N*=3821; Pearson’s *r*=0.16; −log_10_(P-value)=5.09). Overall, the heart-eye phenotypic associations were less significant than those of the other two organs. Detailed results can be found in **Supplementary eFile 7**.

We also performed a secondary PWAS (**Method 3b**) to link the PSCs/IDPs to 117 phenotypes of other organ systems. Detailed results can be found in **Supplementary eFigure 1** and **eFile 8-10**. Finally, we mapped our brain PSCs with 119 conventional MUSE ROIs^53^ using three approaches to interpret the neuroanatomical structures of the brain PSCs. We also discussed our brain PSC PWAS results compared to previous studies by Zhao et al.^8^ and Jaggi et al.^16^ using conventional brain and heart IDPs and performed genetic correlation between our 2003 brain PSCs with 3874 conventional brain IDPs from Smith et al.^27^ **Supplementary eText 2** presents detailed results and discussion for these analyses, providing insights into the added value of our brain PSCs.

### Proteomic map of the 2003 brain PSCs, 82 heart IDPs, and 84 eye IDPs

To strengthen the cross-organ connection further, we performed three ProWASs delineating the pair-wise associations between 2923 Olink plasma proteins and the 2003 brain PSCs, 82 heart IDPs, and 84 eye IDPs (**Method 4a**). We then illustrated the organ-specific and cross-organ interactions by assessing the proteins and RNA expression of the significant proteins using organ/tissue-specific RNA and protein data (**Method 4b**).

For the brain ProWAS, we found 1282 significant associations between 558 brain PSCs and 27 proteins ( P-value<0.05/2003/2923; effective sample size: 51<*N*<4520; −0.42<*r*<0.55) after applying the Bonferroni correction (**Fig. 3a** and **Method 4a**). The most significant brain PSCs (C32_2) showed associations with 10 different proteins. In contrast, the MOG protein had the highest prominence, linking to 410 PSCs. We observed patterns of within-organ specificity and cross-organ interactions. For instance, the KLK6 protein was associated with the brain PSC and exhibited elevated expression levels in both RNA and protein data from brain tissues, demonstrating within-organ specificity. In contrast, the MOG protein, also linked to the brain PSC, was overexpressed in RNA and protein data from the kidney. Detailed statistics of the brain ProWAS are presented in **Supplementary eFile 11**. Additionally, we found that 45 significant brain PSC-protein pairs were replicated in BLSA^45,54^ with a stringent P-value threshold (<0.05/558/27), although the BLSA has a smaller sample size (*N*=924) and used a different proteomic platform (SomaScan); these two sets of estimated β values were significantly correlated (*r*=0.31; P-value=0.03) and showed the same direction of associations (**Supplementary eText 3** for more details).

**Figure 3:**
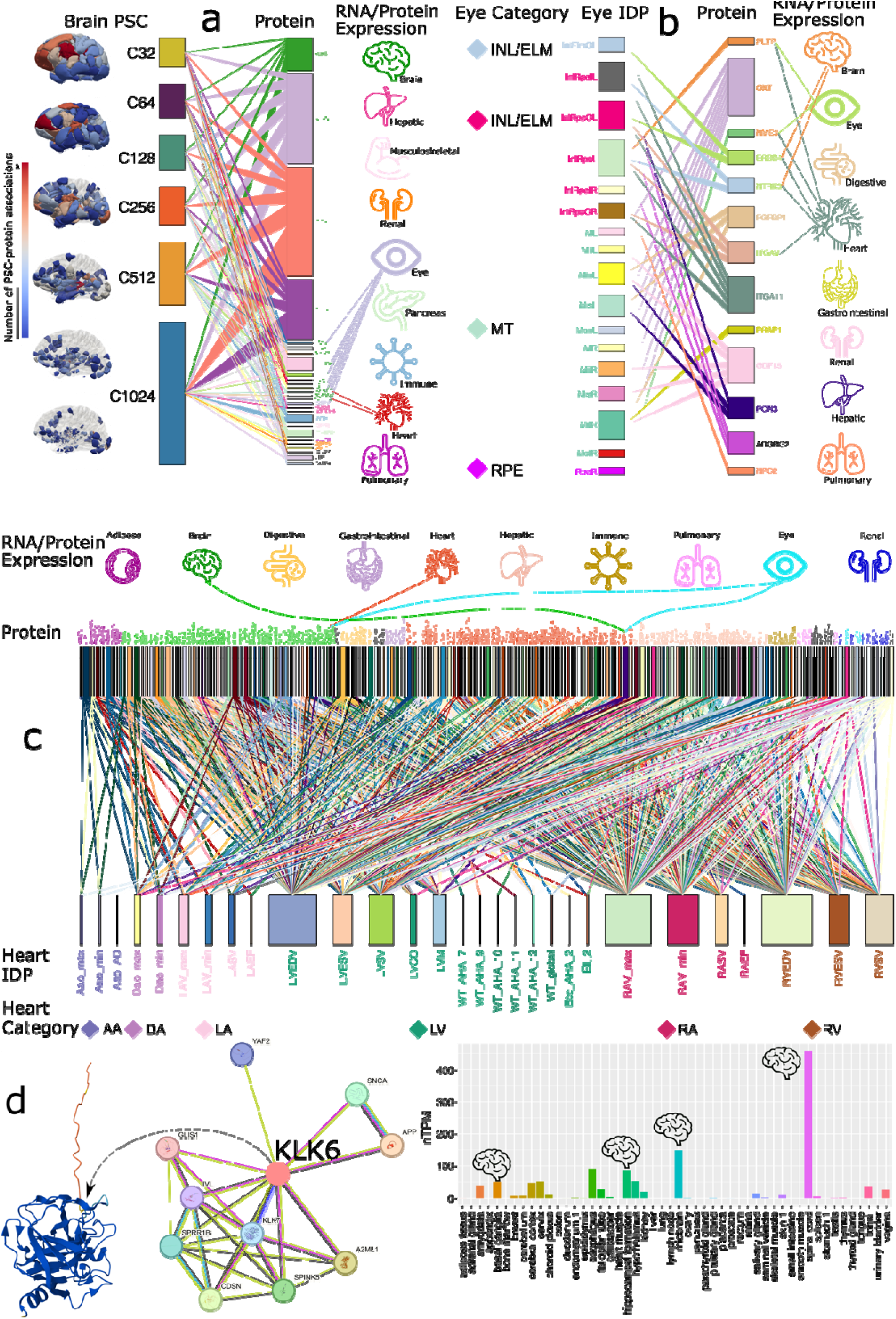
Proteomics associations of the 2003 brain PSCs, 82 heart IDPs, and 84 eye IDPs. **a**) Proteome-wide associations between the 2003 brain PSCs (left panel) and 2923 plasma proteins (middle panel) are shown after the Bonferroni correction (P-value<0.05/2923/2003; effective sample size: 51<*N*<4520; −0.42<*r*<0.55). For each PSC or protein, the percentage of significant associations is proportional to the rectangle’s height. In the right panel, we annotated the protein expression profile in a specific tissue/organ by respective colors of the protein and organ; dotted lines show secondary expression across different organs. **b**) The proteomics associations between the 84 eye IDPs (left panel) and 2923 plasma proteins (middle panel) are shown after the Bonferroni correction (P-value<0.05/2923/84; effective sample size: 48<*N*<4250; −0.47<*r*<0.48). For each IDP or protein, the percentage of significant associations is proportional to the rectangle’s height. In the right panel, we annotated the protein expression in a specific tissue/organ by respective colors of the protein and organ; dotted lines show secondary expression across different organs. **c**) The proteomics associations between the 82 heart IDPs (lower panel) and 2923 plasma proteins (middle panel) are shown after the Bonferroni correction (P-value<0.05/2923/82; effective sample size: 51<*N*<3462; −0.48<*r*<0.61). The percentage of significant associations for each IDP or protein is proportional to the rectangle’s width. In the upper panel, we annotated the protein expression level in a specific tissue/organ by respective colors of the protein and organ; dotted lines showcase secondary expression across different organs. **d**) An example of annotating the protein expression level using GTEx RNA-seq data in 50 different tissues for the KLK6 protein. We used the AlphaFold model to predict the protein structure, followed by a protein-protein interaction network analysis. Additionally, we analyzed the protein expression profiles across 50 diverse tissues to estimate their expression levels. Readers can visualize the 2003 brain PSCs via the BRIDGEPORT knowledge portal: https://labs-laboratory.com//bridgeport. Detailed interpretations of the 84 eye IDPs and 82 heart IDPs are publicly available at https://labs-laboratory.com/medicine/eye and https://labs-laboratory.com/medicine/cardiovascular.

For the eye ProWAS, We discovered 38 significant associations between 17 eye IDPs and 13 proteins after applying the Bonferroni correction (P-value<0.05/2923/84; effective sample size: 48<*N*<4250; −0.47<*r*<0.48) (**Fig. 3b**). The eye IDP measuring the average thickness of the left INL/RPE layer (InlRpeL) was significantly associated with 5 proteins, such as the PLTP protein, which showed high expression values using protein and RNA data from eye-related tissues. The detailed statistics of the eye ProWAS are presented in **Supplementary eFile 12**.

For the heart ProWAS, we identified 866 significant associations that survived the Bonferroni correction between 29 heart IDPs and 196 proteins (P-value<0.05/2923/82; effective sample size: 51<*N*<3462; −0.48<*r*<0.61). The heart IDP (RVEDV: the end-diastolic volume of the right ventricle) was associated with 128 proteins, such as the TGFA protein (**Fig. 3c**). Detailed results can be found in **Supplementary eFile 13**.

We present the organ-specific expression at RNA gene expression levels for one significant protein (i.e., the KLK6 protein) in **Fig. 3d** and provide a detailed discussion of these ProWAS findings in the Discussion section. KLK6 was significantly associated with 123 brain PSCs. For example, C32_1 was positively associated with KLK6 (*β*=77.82±10; P-value=1.10×10^−14^). We then performed a protein-protein interaction analysis^55^, showing its direct and indirect interactions with other proteins, such as the KLK7 protein from the same family, which may play similar roles in amyloid precursor protein, myelin basic protein, gelatin, casein, and extracellular matrix proteins. Finally, we showed the RNA tissue-specific enrichment of this protein in 50 different tissues using the HPA and GTEx data (**Method 4b**), with the most prominent enrichment in the spinal cord (nTPM=475.6), midbrain (nTPM=148.9), and hippocampus (nTPM=85.8) (**Fig. 3d**).

### Genetic architecture of the 2003 brain PSCs, 82 heart IDPs, and 84 eye IDPs

Using genome-wide common genetic variants, we aimed to elucidate the genetic architecture of the brain-heart-eye axis by conducting GWAS to identify shared genetic signals across the three organs. Subsequently, we performed several post-GWAS analyses to validate these genetic signals.

For the three primary GWASs (**Method 5a**) using European ancestry populations, we identified 5854 (P-value<5×10^−8^/2003), 214 (P-value<5×10^−8^/82), and 1888 (P-value<5×10^−8^/84) genomic locus-PSC/IDP associations for the 2003 brain PSCs, 82 heart, and 84 eye IDPs, respectively. We denoted the genomic loci using their top lead SNPs (**Supplementary eMethod 1**) defined by FUMA; the genomic loci are presented in **Supplementary eFile 14-16**. We found that 32, 94, and 29 common cytogenetic regions (based on the GRCh37 cytoband) were jointly linked to the brain-heart, brain-eye, and heart-eye GWAS, respectively (**Fig. 4a-c**).

**Figure 4:**
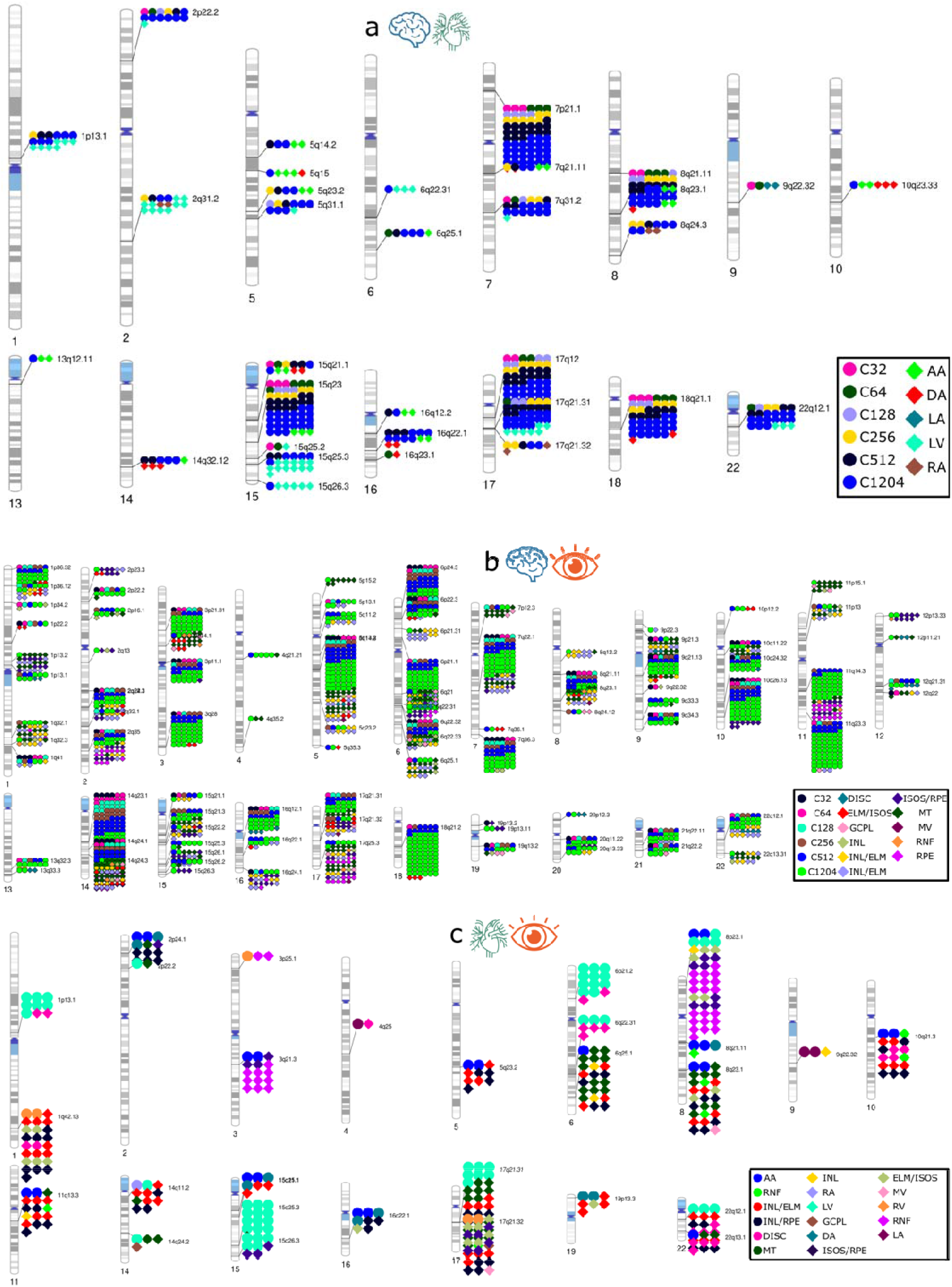
Genome-wide associations of the 2003 brain PSCs, 82 heart IDPs, and 84 eye IDPs **a**) Cytogenetic regions where the genomic region was jointly linked to the heart IDPs and brain PSCs. Bonferroni correction was applied to denote significant genomic loci associated with the brain PSCs (P-value<5×10^−8^/2003) and heart IDPs (P-value<5×10^−8^/82). **b**) Cytogenetic regions where the genomic region was jointly linked to the eye IDPs and brain PSCs. Bonferroni correction was applied to denote significant genomic loci associated with the brain PSCs (P-value<5×10^−8^/2003) and eye IDPs (P-value<5×10^−8^/84). **c**) Cytogenetic regions where the genomic region was jointly linked to eye IDPs and heart IDPs. Bonferroni correction was applied to denote significant genomic loci associated with the heart IDPs (P-value<5×10^−8^/82) and eye IDPs (P-value<5×10^−8^/84). Readers can visualize the 2003 brain PSCs via the BRIDGEPORT knowledge portal: https://labs-laboratory.com//bridgeport. Detailed interpretations of the 84 eye IDPs and 82 heart IDPs are publicly available at https://labs-laboratory.com/medicine/eye and https://labs-laboratory.com/medicine/cardiovascular.

The primary GWAS conducted among populations of European ancestry demonstrated robustness across several sensitivity analyses and underscored the necessity of collecting data from non-European populations. We first calculated the genomic inflation factor (λ) and LDSC^56^ intercept (*b*) for the 2003 brain PSC [λ=1.095 (1.007-1.233); *b*=1.011 (0.982-1.0467)], 82 heart IDP GWASs [λ=1.0649 (1.0105-1.1459); *b*=1.004 (0.9815-1.0233)], and 84 eye IDP GWASs [λ=1.119 (1.028-1.200); *b*=1.019 (0.991-1.045)]. All LDSC intercepts were close to 1, indicating no substantial genomic inflation. We already scrutinized the brain GWAS in our previous study^1^. In the current study, we checked the robustness of the heart and eye IDP GWASs in five additional sensitivity check analyses (**Method 5a**). For the heart IDP GWAS, we observed high CRs based on the P-value and Pearson’s *r* for the two sets of β values (*r-*β) in split-sample (0.82<CR-P<0.97; *r-*β=0.94) and sex-stratified (0.76<CR-P<0.94; *r-β*=0.78) sensitivity analyses. Furthermore, our GWAS β values were highly correlated with those from a prior heart IDP GWAS by Zhao et al.^8^ (*r*-*β*=0.90). However, our GWAS’s generalizability to non-European ancestries was limited due to limited sample sizes in non-European populations (0.02<CR-P<0.50) based on P-values, underscoring the need to increase sample sizes for future GWAS in underrepresented ethnic groups (*r*-*β*=0.84) (**Supplementary eText 4a** and **eFile 17-20**). For the eye IDP GWAS, we observed high CRs based on the P-values and Pearson’s *r-β* values across split-sample (0.86<CR-P<0.98; *r-β*=0.97±0.04), sex-stratified (0.80<CR-P<0.95; *r-β*=0.96±0.11), and GWAS method-specific (CR-P=1 between PLINK linear model and fastGWA^57^ linear mixed model; *r-β*=0.99±0.0008) sensitivity analyses. In addition, our GWAS reproduced discoveries from a previous eye IDP GWAS study by Zhao et al.^58^ (CR-P=0.72; *r-β*=0.996). The generalizability of our GWAS to non-European ancestries was restricted (0.11< CR-P< 0.63) according to the P-values, possibly due to limited sample sizes. This was supported by the high correlation (*r-*β=0.95) observed between the two sets of β estimates (**Supplementary eText 4b** and **eFile 21-25**). Manhattan and QQ plots are publicly available at the MEDICINE knowledge portal: https://labs-laboratory.com/medicine/. All subsequent post-GWAS analyses utilized the primary GWAS results from European ancestry cohorts.

To uncover the phenome-wide associations of the heart and eye IDP-linked genomic loci in the previous literature, we performed a PheWAS (**Method 5b** and **Supplementary eText 5**) look-up analysis using the GWAS Atlas^59^ platform for these top lead SNPs. This identified 2939 and 8493 previous SNP-trait associations in prior GWASs for the heart and eye IDP GWASs, respectively, as detailed in **Supplementary Figure 2, eText 5**, and **eFile 26-27**.

### SNP-based heritability estimates of the 2003 brain PSCs, 82 heart IDPs, and 84 eye IDPs

We computed the SNP-based heritability to quantify the proportion of phenotypic variance attributable to common genetic variants across the genome and organs. Among the 2003 brain PSCs, the GCTA^60^ software revealed significant SNP-based heritability in 1897 PSCs (*h*^2^=0.30±0.16; P-value<0.05/2003) (**Method 5c**). Among the 82 heart IDPs, 80 demonstrated significant heritability estimates (*h*^2^=0.25±0.12; P-value<0.05/82) (**Fig. 5b**), and all 84 eye IDPs demonstrated significant heritability estimates (*h*^2^=0.44±0.12; P-value<0.05/84) (**Fig. 5a-c**).

**Figure 5:**
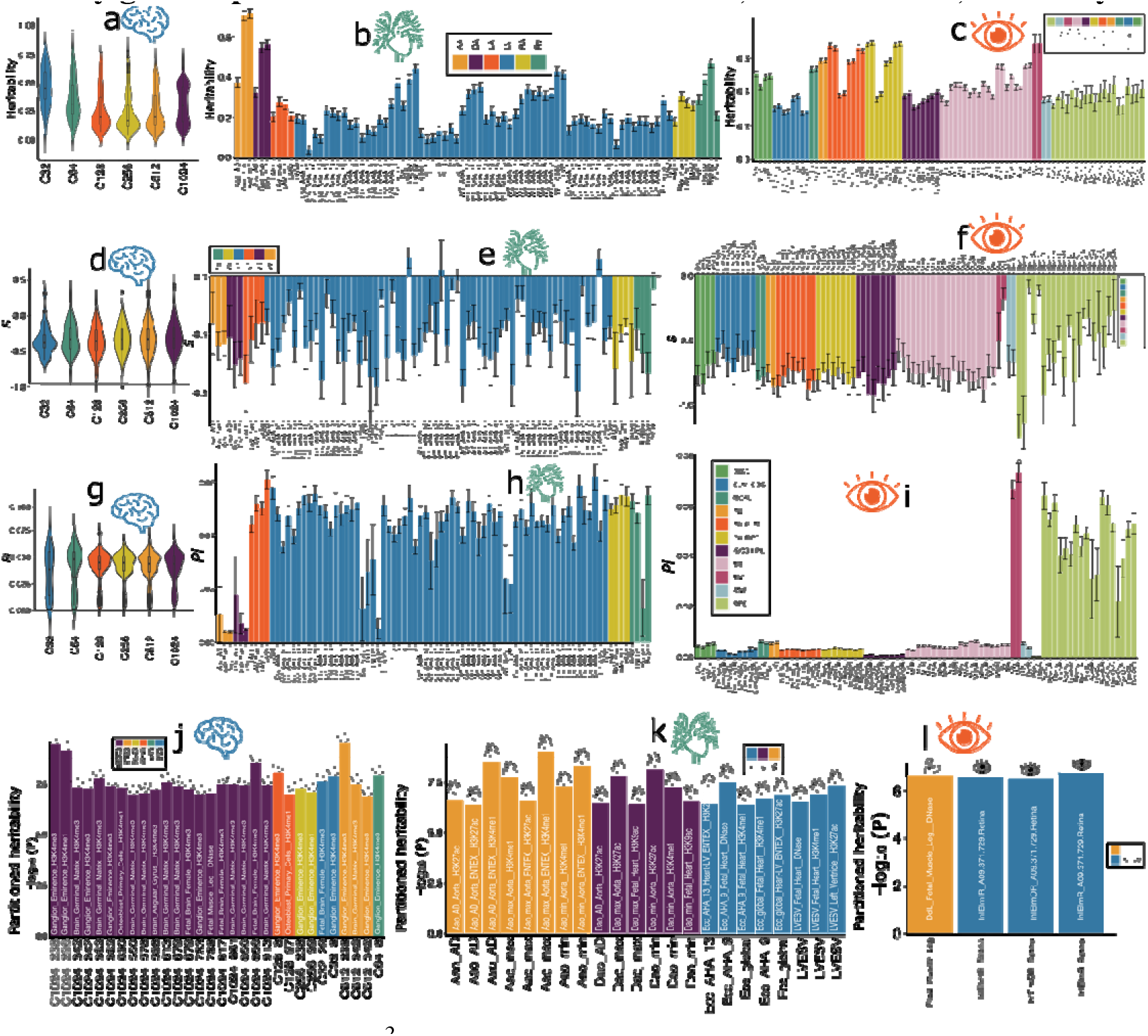
Key genetic parameters of the 2003 brain PSCs, 82 heart IDPs, and 84 eye IDPs **a-c**) The SNP-based heritability (*h^2^*) was estimated using the GCTA software for the 2003 brain PSCs (C32-1024), 82 heart IDPs, and 84 eye IDPs. **d-f**) The selection signature (*S*) was estimated using the SBayesS software for the 2003 brain PSCs, 82 heart IDPs, and 84 eye IDPs. **g-i**) The polygenicity (*Pi*) was estimated using the SBayesS software for the 2003 brain PSCs, 82 heart IDPs, and 84 eye IDPs. **j-l**) The tissue-specific partitioned heritability using gene expression and chromatin data was estimated using the LDSC software for the 2003 brain PSCs, 82 heart IDPs, and 84 eye IDPs. We further categorized brain PSCs into different scales of the *C* parameters. The 82 heart IDPs are divided into 6 segmented heart regions: the left ventricle (LV), right ventricle (RV), left atrium (LA), right atrium (RA), descending aorta (DA), and ascending aorta (AA). The error bar denotes the standard error of the estimate of the mean. The 84 eye IDPs were classified into 11 high-level phenotype categories. Readers can visualize the 2003 brain PSCs via the BRIDGEPORT knowledge portal: https://labs-laboratory.com//bridgeport. Detailed interpretations of the 84 eye IDPs and 82 heart IDPs are publicly available at https://labs-laboratory.com/medicine/eye and https://labs-laboratory.com/medicine/cardiovascular.

For the brain PSCs, The heritability estimates spanned from a coarse to a fine scale (C32 to C1024), with the highest *h*^2^ observed at the C32 scale (*h*^2^=0.45±0.16; P-value<1.57×10^−8^) and the lowest *h*^2^ at the C256 scale (*h*^2^=0.24±0.16; P-value<2.27×10^−5^). The heritability estimates varied across the 82 heart IDPs for the 6 high-level categories, with the highest *h^2^* observed for the ascending aorta (AA, *h^2^*=0.60±0.20; P-value<1.22×10^−56^) and the lowest *h^2^*for the IDPs of the left ventricle (LV, *h^2^*=0.21±0.09; P-value<1.42×10^−5^). The heritability estimates also varied across the 84 eye IDPs for the 11 high-level categories, with the highest *h^2^* observed for macular volume (MV, *h^2^*=0.68±0.0008; P-value<1.47×10^−36^) and the lowest *h^2^* for the external limiting membrane layer (ELM) and inner & outer segment (ISOS) layers (ELM/ISOS, *h^2^*=0.30±0.04; P-value<1.24×10^−81^).

The detailed statistics of the SNP-based heritability estimates are presented in **Supplementary eFile 28-29**. The results generated by LDSC are detailed in **Supplementary eFile 30-31** for the heart and eye IDPs, respectively. Overall, LDSC-derived results using summary-level data were lower than (e.g., heart PSCs: *h^2^*=0.14±0.07; *h^2^*=0.22±0.06) but highly correlated with (*r*=0.95; P-value=4.18×10^−42^; *r*=0.88; P-value<3.79×10^−28^) the GCTA estimates, which used individual-level data. This was consistent with previous observations on brain IDP GWASs^61,26^ which may be attributed to the different model assumptions and LD information of different approaches^62^. Furthermore, our GCTA-based *h^2^* estimates for the heart and eye IDPs were highly correlated with those obtained from Zhao et al.^8^ (heart IDPs: *N*=82 overlapping IDPs, *r*=0.91, P-value<1×10^−10^) and Zhao et al.^58^ (eye IDPs: *N*=46 overlapping IDPs, *r*=0.99, P-value<1×10^−10^), both using GCTA (**Supplementary eFigure 3**).

### Selection signatures and polygenicity estimates of the 2003 brain PSCs, 82 heart IDPs, and 84 eye IDPs

To understand the evolutional processes of these PSCs/IDPs, specifically regarding how traits evolve and adapt over time through natural selection and genetic variation, we used the SBayesS^63^ method to compute the selection signature (*S*) and polygenicity (*Pi*) for the 2003 brain PSCs, 82 heart IDPs, and 84 eye IDPs (**Method 5d**). A signature of negative (or purifying) selection prevents mutations with significant harmful effects from becoming prevalent in the population, while positive selection promotes the propagation of those with beneficial effects.

We obtained selection signature estimates of *S*=−0.32±0.22, *S*=−0.40±0.20, and *S*=−0.69±0.20 for the 2003 brain PSCs, 82 heart IDPs, and 84 eye IDPs (pairwise estimates P-value < 0.001). Similarly, we observed polygenicity estimates of *Pi*=0.040±0.015, *Pi*=0.043±0.014, and *Pi*=0.014±0.021 for the 2003 brain PSCs, 82 heart IDPs, and 84 eye IDPs. These findings offer a comparative assessment of critical genetic metrics to elucidate the genetic code governing the three organs (**Fig. 5d-f**). Detailed statistics are presented in **Supplementary eFile 32-34**.

### Tissue-specific enrichment of partitioned heritability of the 2003 brain PSCs, 82 heart IDPs, and 84 eye IDPs

To provide additional biological validation for our GWAS discoveries of this brain-heart-eye axis, we conducted partitioned heritability analyses^64^ (**Method 5e**) to estimate the heritability enrichment of genetic variants regarding 205 tissue-specific gene expression data (e.g., lung, brain, and liver)^65^ and 489 tissue-specific chromatin annotation data (e.g., heart-atrial_H3K4me3)^66,67^ (**Fig. 5j-l**).

For the brain PSCs, we found 30 significant heritability enrichment (P-value < 0.05/205/489) in various brain tissues (e.g., angular gyrus and ganglion eminence) and musculoskeletal tissues (e.g., fetal muscle leg) at different chromatin states (**Fig. 5j**). For example, C32_8 showed significant heritability enrichment (P-value=1.33×10^−8^) in ganglion eminence derived from primary culture neurospheres of peaks for trimethylated lysine 4 on histone H3 (H3K4me3). For the heart IDPs, 12 significant heritability enrichment was found in multiple heart tissues in the H3K4me1 (e.g., P-value=9.06×10^−5^ for the fetal heart and the end-systolic volume of the LV), and acetylated lysine 27 on histone H3 (H3K27ac, P-value=1.78×10^−8^ for the aorta and the descending aorta maximum area; **Fig. 5k**). For the eye IDPs, we found that 4 significant heritability enrichment in retina-related tissues and fetal muscle (**Fig. 5l**). Detailed statistics are presented in **Supplementary eFile 35-37**. Those organ-specific enrichment patterns largely provide biological validation of our primary GWAS results.

### Gene-drug-disease network highlights drug repurposing potential for cross-organ diseases

We conducted a gene-drug-disease enrichment analysis^68^ using genes associated with the 2003 brain PSCs, 82 heart IDPs, and 84 eye IDPs within the targeted gene sets of drug categories from the DrugBank database^69^. This analysis constructed a gene-drug-disease network to identify potentially repurposable drugs, which has been shown to increase drug development success in the literature^70,71^ (**Method 5f**).

For the brain PSCs, the gene-drug-disease network identified 1207 significant pairs of gene-drug-disease interactions between 57 unique brain PSCs, 27 genes, and 78 ICD disease categories. For the eye IDPs, we found 327 significant interactions between 4 unique IDPs, 8 genes, and 33 ICD disease categories. Finally, for the heart IDPs, we identified 8 significant interactions between one gene, two IDPs, and 1 ICD disease category (**Fig. 6**). Here, we showcased several drugs and/or small molecules. For example, the *GALR3* gene linked to the brain PSC (C32_2) served as the target gene for *HT-2157* (NCT number: NCT01413932; status: completed). This drug, a selective non-peptide antagonist for the GAL-3 receptor, was tested for treating major depressive disorder (ICD: F00-09). Another example was the *MAPT* gene associated with both brain PSC (e.g., C128_106) and eye IDP (e.g., GCPL), which was the target gene for *AL-408* (small molecule drug) treating cerebrovascular disorders and cognition disorders (G40-47 and G10-14). The *MAPT* gene was also used to develop another drug, davunetide, which has been investigated in clinical trials aimed at treating various diseases related to the central nervous system, including progressive nonfluent aphasia, progressive supranuclear palsy, corticobasal degeneration syndrome, and frontotemporal dementia with parkinsonism. The *CCL11* gene associated with the brain PSC (C1024_275) was used to develop the drug bertilimumab, treating multiple eye diseases, including glaucoma (ICD-code: H).

**Figure 6:**
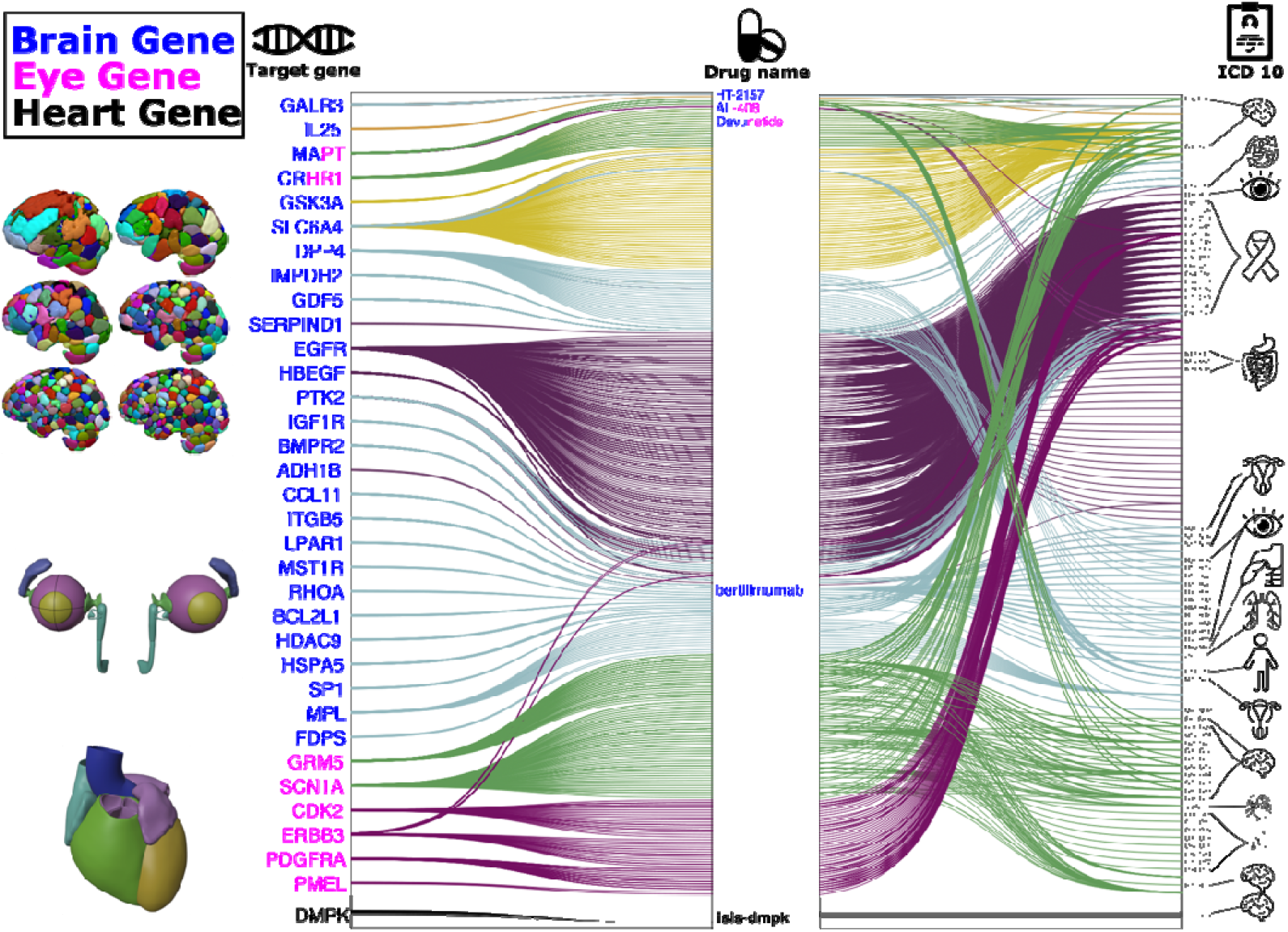
The gene-drug-disease network of the 2003 brain PSCs, 82 heart IDPs, and 84 eye IDPs The gene-drug-disease network reveals a broad spectrum of gene, drug, and disease interactions across the 2003 brain PSCs, 84 eye IDPs, 82 heart IDPs, and diseases beyond the three organs. The ICD code icons symbolize disease categories linked to the primary organ systems. All presented genes passed the FDR-corrected P-value threshold and were pharmaco-genetically associated with drug categories in the DrugBank database. Abbreviation: ICD: International Classification of Diseases. We showcased four representative drugs from the genes linked to the three organs.

Finally, the heart IDP-related (LA maximum/minimum volum) gene *DMPK* was enriched in several drugs, including *isis-dmpk,* for treating diseases of the myoneural junction and muscle (ICD-code: G70-G73). Detailed results are presented in **Supplementary eFile 38-40**.

### Genetic correlation between the 2003 brain PSCs, 82 heart IDPs, and 84 eye IDPs

We estimated the pairwise PSCs/IDPs genetic correlation^56^ (*g_c_*, **Method 5g**) between the three organs. This analysis further supports the cross-organ interconnections and echos their phenotypic associations, supporting Cheverud’s Conjecture^72^.

For the brain-heart correlation, we found 30 significant genetic correlations [*g_c_*=0.31 (−0.60, 0.38)] after applying the Bonferroni correction (P-value < 0.05/2003) (**Fig. 7a** and **Supplementary eFile 41**). The significant genetic correlations largely encompassed deep subcortical structures, the inferior frontal cortex, the insula cortex, and the temporal pole. For example, the largest negative correlation was found between the circumferential strain AHA_2 (Ecc_AHA_2) of the LV and C256_57 (*g_c_*=−0.60±0.13; P-value=7.16×10^−6^) and the strongest positive correlation between the descending aorta distensibility (Dao_AD) and C1024_827 (*g_c_*=0.38±0.09; P-value=1.32×10^−5^). For the heart IDPs, the most enriched associations were linked to the ascending and descending aorta (97%).

**Figure 7:**
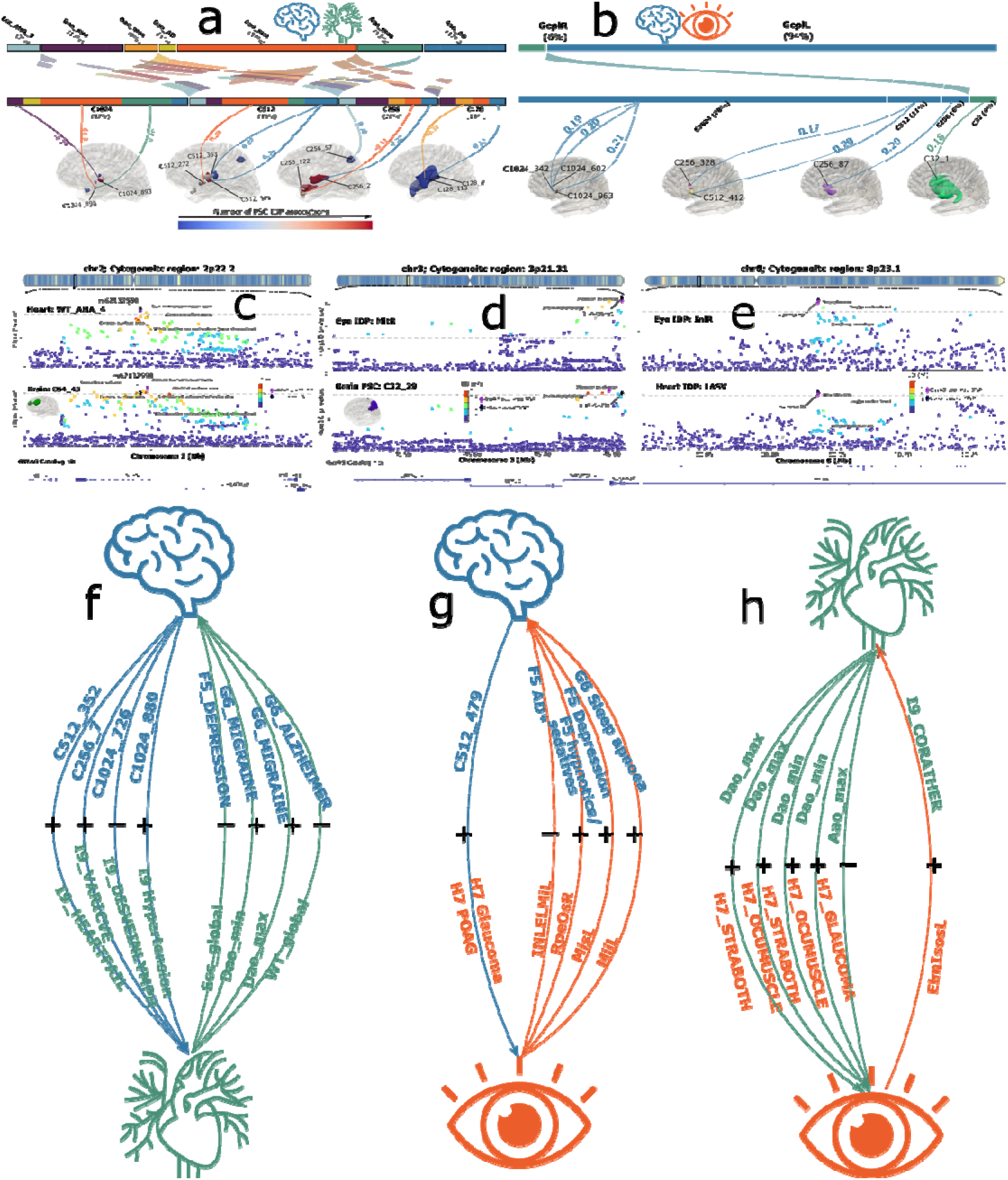
Genetic correlation and causality of the 2003 brain PSCs, 82 heart IDPs, and 84 eye IDPs **a**) The genetic correlation (*g_c_*) between the 2003 brain PSCs and 82 heart IDPs. Significant results after the Bonferroni correction (P-value<0.05/2003) are shown. With the application of a more stringent correction threshold (P-value < 0.05/2003/82), no statistical significance was observed. **b**) Significant genetic correlations (*g_c_*) between the 2003 brain PSCs and 84 eye IDPs after the Bonferroni correction (P-value<0.05/2003). With the application of a more stringent correction threshold (P-value < 0.05/2003/84), no statistical significance was observed. **c**) Genetic colocalization was evidenced at one genomic locus (2p22.2) between the mean myocardial wall thickness aha4 (WT_AHA_4) and C64_45 (PP.H4.ABF=0.98). **d**) Genetic colocalization was evidenced at one locus (3p21.31) between the right macular thickness at the inner temporal subfield (MitR) and C32_29. **e**) Genetic colocalization was evidenced at one locus (8p23.1) between the right inner nuclear layer thickness at the inner temporal subfield (InlR) and the LA stroke volume (LASV). **f**) The *Brain2Heart* causal relationship tested 2003 brain PSCs as exposure variables and 45 heart diseases (code: I9) as outcome variables; the *Heart2Brain* causal link is established between 82 heart IDPs as exposure variables and 41 brain diseases as outcome variables. **g**) The *Brain2Eye* causal relationship tested 2003 brain PSCs as exposure variables and 32 eye diseases (code: H7) as outcome variables; the *Eye2Brain* causal link is established between 84 eye IDPs as exposure variables and 41 brain diseases as outcome variables. **h**) The *Heart2Eye* causal relationship tested 82 heart IDPs as exposure variables and 32 eye diseases (code: H7) as outcome variables; the *Eye2Heart* causal link is established between 84 eye IDPs as exposure variables and 45 heart diseases as outcome variables. Solid arrow lines represented significant results with more stringent Bonferroni correction (i.e., P-value<0.05/*N*, where *N* represents the larger number of IDPs/PSCs/diseases), and dotted arrow lines for a less stringent correction (i.e., P-value<0.05/*M*, where *M* represents the smaller number of IDPs/PSCs/diseases). The symbols + and – signify positive and negative causal associations between the traits tested. Readers can visualize the 2003 brain PSCs via the BRIDGEPORT knowledge portal: https://labs-laboratory.com//bridgeport. Detailed interpretations of the 84 eye IDPs and 82 heart IDPs are publicly available at https://labs-laboratory.com/medicine/eye and https://labs-laboratory.com/medicine/cardiovascular.

For the brain-eye correlation, we found 18 significant positive genetic correlations (*g_c_*=0.19±0.01) after applying the Bonferroni correction (P-value < 0.05/2003) (**Fig. 7b** and **Supplementary eFile 42**). These correlations included GcplL of the eye and brain regions covering deep subcortical structures. Examples were between GcplL and C1024_963 (*g_c_*=0.21±0.04; P-value=1.65×10^−6^) and between GcplR and C32_1 (*g_c_*=0.18±0.04; P-value=4.98×10^−5^). The PSCs of the visual cortex exhibited signals of lesser significance, which did not withstand the Bonferroni correction. This could be attributed to potential limitations in power when using GWAS summary statistics via LDSC compared to the phenotypic associations derived from raw phenotype data. However, our results evidenced a substantial alignment between the phenotypic and genetic correlations for the pairwise PSC-IDP. For example, GcplL showed a significant phenotypic correlation (*r*= 0.11; P-value=1.10×10^−12^) with C32_1, echoing their genetic correlation mentioned above (*g_c_*=0.18±0.04; P-value=4.95×10^−5^).

For the heart-eye correlation, we found only 3 significant positive genetic correlations after Bonferroni correction (P-value < 0.05/84). The left macular thickness at the outer temporal subfield (MotL) was genetically correlated with the RA stroke volume (RASV) (*g_c_*=0.36±0.09; P-value=4.00×10^−5^) and RA maximum volume (RAV_max) (*g_c_*=0.32±0.08; P-value=6.00×10^−5^). The left macular thickness at the inner temporal subfield (MitL) was genetically correlated with the RA maximum volume (RAV_max) (*g_c_*=0.30±0.08; P-value=3.00×10^−4^) (**Supplementary eFile 43**).

### The genetic colocalization between the 2003 brain PSCs, 82 heart IDPs, and 84 eye IDPs

To identify potential shared causal variants, we applied the Approximate Bayes Factor colocalization^73^ method between each PSC-IDP and IDP-IDP pair of the three organs (**Method 5h**).

For the brain-heart colocalization, we found 1822 colocalization signals using the suggested posterior possibility threshold^73^ (PP.H4.ABF>0.8, which tests the hypothesis H4: a shared causal variant is associated with both traits) (**Supplementary eFile 44**). Genetic colocalization was observed at the genomic locus (2p22.2) between the mean myocardial wall thickness AHA_4 (WT_AHA_4) of the LV and C64_45 of the brain (PP.H4.ABF=0.9804). This discovery highlights the potential causal variant (rs62132550), aligning with the physical position of the *STRN* gene. SNPs in strong linkage disequilibrium with this causal SNP have previously shown associations with diverse traits documented in the GWAS Catalog, encompassing brain morphological features like white matter diffusivity and cortical surface area, as well as traits such as red cell distribution width and alanine transaminase levels (**Fig. 7c**).

For the brain-eye colocalization, we found 5368 colocalization signals (**Supplementary eFile 45**). We illustrate this at the locus (3p21.31) between the right macular thickness at the inner temporal subfield (MitR) and C32_29 of the brain, which outlines the posterior cortex of the brain (PP.H4.ABF=0.9966, **Fig. 7d**). This discovery highlights the potential causal variant located at chr3:45795731, aligning with the physical position of the *SLC6A20* gene. SNPs in strong linkage disequilibrium with this causal SNP have previously been linked to various traits in the GWAS Catalog^74^. Notably, the top lead SNP (rs17279437) was associated with macular measurement^75^ and retinal disorders^76^, and another SNP (rs73058498; mapped gene: *SACM1L*) was associated with brain morphology^77^. Other SNPs within this locus were also previously associated with body mass index^78^ and systolic blood pressure^79^.

For the heart-eye colocalization, we identified 538 colocalization signals (**Supplementary eFile 46**). For example, we showed one locus (8p23.1) between the right inner nuclear layer thickness at the inner temporal subfield (InlR) and the LA stroke volume (LASV; PP.H4.ABF=0.9980, **Fig. 7e**). This discovery highlights the potential causal and top lead variant (rs2975648), mapping to the *MSRA* gene. SNPs in strong linkage disequilibrium with this causal SNP have been linked to neuroticism, triglycerides level, and smoking initiation in previous literature.

### Cross-organ causal network between the brain, heart, and eye

We established 6 bi-directional causal networks between the PSC/IDPs and disease endpoints of the three organs using FinnGen^46^; PGC^47^ data were used as replications for the brain-related causal relationships. We established these networks by employing two-sample Mendelian randomization analyses^80^ (**Method 5i**).

Within the *Brain2Heart* network, we found potential causal relationships from C512_352 to heart failure (I9_HEARTFAILURE) [P-value=8.29×10^−5^<0.05/603 PSCs; OR (95% CI)=1.28 (1.14, 1.46); number of IVs=9], C256_7 to varicose veins (I9_VARICAE) [P-value=2.70×10^−5^; OR (95% CI)=1.17 (1.08, 1.25); number of IVs=21], C1024_726 to diseases of vein, lymphatic vessels, and lymph nodes (I9_DISVEINLYMPH) [P-value=1.20×10^−5^; OR (95% CI)=0.83 (0.77, 0.90); number of IVs=9], and C1024_880 to hypertension (I9_HYPTENSESS) [P-value=1.50×10^−5^; OR (95% CI)=1.09 (1.05, 1.13); number of IVs=28] (**Fig. 7f**).

Within the *Heart2Brain* network, we found several significant causal signals (P-value=0.05/41 unique brain disease endpoints). We found potential causality from the global circumferential strain of the LV (Ecc_global) to recurrent depression (F5_DEPRESSION) [P-value=5.60×10^−5^; OR (95% CI)=0.78 (0.70, 0.88); number of IVs=11], the Dao_min to migraine with aura (G6_MIGRANE_WITH_AURA) [P-value=1.50×10^−4^; OR (95% CI)=1.27 (1.12, 1.43); number of IVs=16], the Dao_max to migraine with aura (G6_MIGRANE_WITH_AURA) [P-value=1.90×10^−4^; OR (95% CI)=1.28 (1.12, 1.45); number of IVs=16], and the global mean of the myocardial wall thickness of LV (WT_global) to AD [P-value=1.22×10^−3^; OR (95% CI)=0.70 (0.56, 0.87); number of IVs=8] (**Fig. 7f**).

Within the *Brain2Eye* network, we identified potential causal relationships from C512_479 (medial orbital frontal region) to glaucoma [P-value=6.16×10^−6^<0.05/603 PSCs; OR (95% CI)=1.35 (1.18, 1.53); number of IVs=7] and primary open-angle glaucoma (POAG) [P-value=1.12×10^−5^; OR (95% CI)=1.67 (1.33, 2.11); number of IVs=7] (**Fig. 7g**).

Within the *Eye2Brain* network, we found potential causal signals from the right retinal pigment epithelium (RPE) thickness at the outer superior subfield (RpeOsR) to the use of hypnotics and sedatives [P-value=2.26×10^−4^<0.05/75 IDPs; OR (95% CI)=1.09 (1.04, 1.13); number of IVs=9], the left macular thickness at the inner superior subfield (MisL) to depression [P-value=4.08×10^−4^; OR (95% CI)=1.14 (1.06, 1.22); number of IVs=27], the left macular thickness at the inner inferior subfield (MiiL) to sleep apnoea [P-value=4.53×10^−4^; OR (95% CI)=1.11 (1.05, 1.17); number of IVs=29], and the left INL/ELM thickness at the inner subfield (INLEMLiL) to AD [P-value=1.21×10^−3^<0.05/41; OR (95% CI)=0.83 (0.74, 0.93); number of IVs=56]. Notably, this INLEMLiL-AD causal relationship was independently replicated using the AD case-control GWAS^81^ from the PGC [P-value=8.31×10^−3^; OR (95% CI)=0.93 (0.88, 0.98); number of IVs=64] (**Fig. 7g**).

For the *Heart2Eye* network, we found potential causal relationships from the descending aorta maximum area (Dao_max) to strabismus (H7_STRABOTH) [P-value=5.10×10^−4^<0.05/32 eye diseases; OR (95% CI)=1.27 (1.11, 1.45); number of IVs=16] and disorders of ocular muscles, binocular movement, accommodation and refraction (H7_OCUMUSCLE) [P-value=5.40×10^−4^; OR (95% CI)=1.17 (1.07, 1.28); number of IVs=16]. Causation was also established from the descending aorta minimum area (Dao_min) to strabismus (H7_STRABOTH) [P-value=8.80×10^−4^; OR (95% CI)=1.24 (1.09, 1.40); number of IVs=16] and disorders of ocular muscles, binocular movement, accommodation and refraction (H7_OCUMUSCLE) [P-value=5.80×10^−4^; OR (95% CI)=1.16 (1.06, 1.26); number of IVs=16].

Finally, ascending aorta maximum area (Aao_max) to glaucoma (H7_GLAUCOMA) [P-value=8.60×10^−4^; OR (95% CI)=0.89 (0.84, 0.95); number of IVs=38] (**Fig. 7h**).

For the *Eye2Heart* network, we identified potential causal relationship from the lef average elmisos thickness (ElmIsosL) to coronary atherosclerosis (I9_CORATHER) [P-value=5.40×10^−4^<0.05/75 eye IDPs; OR (95% CI)=1.17 (1.07, 1.28); number of IVs=21] (**Fig. 7h**). Details of the results, including all five different Mendelian randomization estimators, are shown in **Supplementary eFile 47-52.** Detailed quality check analyses for each significant causal relationship are presented in **Supplementary eFigure 4-23**. **Supplementary eText 6** compares our causal analyses with those conducted in the previous studies by Zhao et al.^8^ and Lin et al^82^.

### Multi-organ features improve prediction for 14 systemic diseases and 8 cognitive scores

We assessed the enhanced predictive capabilities of integrating the brain PSCs, heart IDPs, eye IDPs, and their respective PRSs (**Method 5j**), compared to single-organ/omics features. This assessment aimed to predict 14 systemic disease categories based on the ICD-10 code and 8 cognitive scores in both univariate and multivariate machine learning and statistical models. The definition of patient and healthy control groups is detailed in **Method 6** and **Supplementary eTable 2**. Of note, these tasks aim to test our hypothesis that multi-organ, multi-omics data can improve model performance compared to single-organ/omics data. The overall model performance is modest, considering the inherent difficulty of the tasks, the heterogeneity of the patient groups, and the use of baseline machine learning models. In addition, we did not include proteomics data because of their high missing value rates across individuals and proteins.

We first evaluated the prediction power (i.e., the incremental *R^2^*; **Method 6a**) of the individual PSC/IDP/PRS to predict the 14 disease categories. Overall, we found that the most predictive features corresponded to the diseases of their respective primarily affected organs. For example, the 10 representative brain PSCs and 10 brain PSC-PRSs demonstrated the strongest predictive capacity for mental and behavioral disorders (ICD-10 code: F). Detailed results and interpretation are presented in **Supplementary eText 7, eFigure 24**, and **eFile 53-55**.

We then evaluated the predictive capacity of cross-organ imaging features (IDPs or PSCs) and their respective PRSs in classifying the 14 disease categories by combining different sets of features for different organs (**Method 6b**). Several observations were made. Firstly, integrating multi-omics features, such as PRSs with PSCs/IDPs, enhanced predictions for conditions like mental and behavioral disorders (ICD: F) and metabolic diseases (ICD: E), specifically for brain PSCs and PRSs. Secondly, cross-organ imaging features demonstrated greater predictive power than single-organ features, exemplified by improved predictions for mental and behavioral disorders using brain PSCs and heart IDPs. Lastly, the best performance was achieved by combining cross-organ imaging features and PRSs together, such as using brain PSCs, eye IDPs, and their PRSs to predict mental and behavioral disorders (**Fig. 8a**).

**Figure 8:**
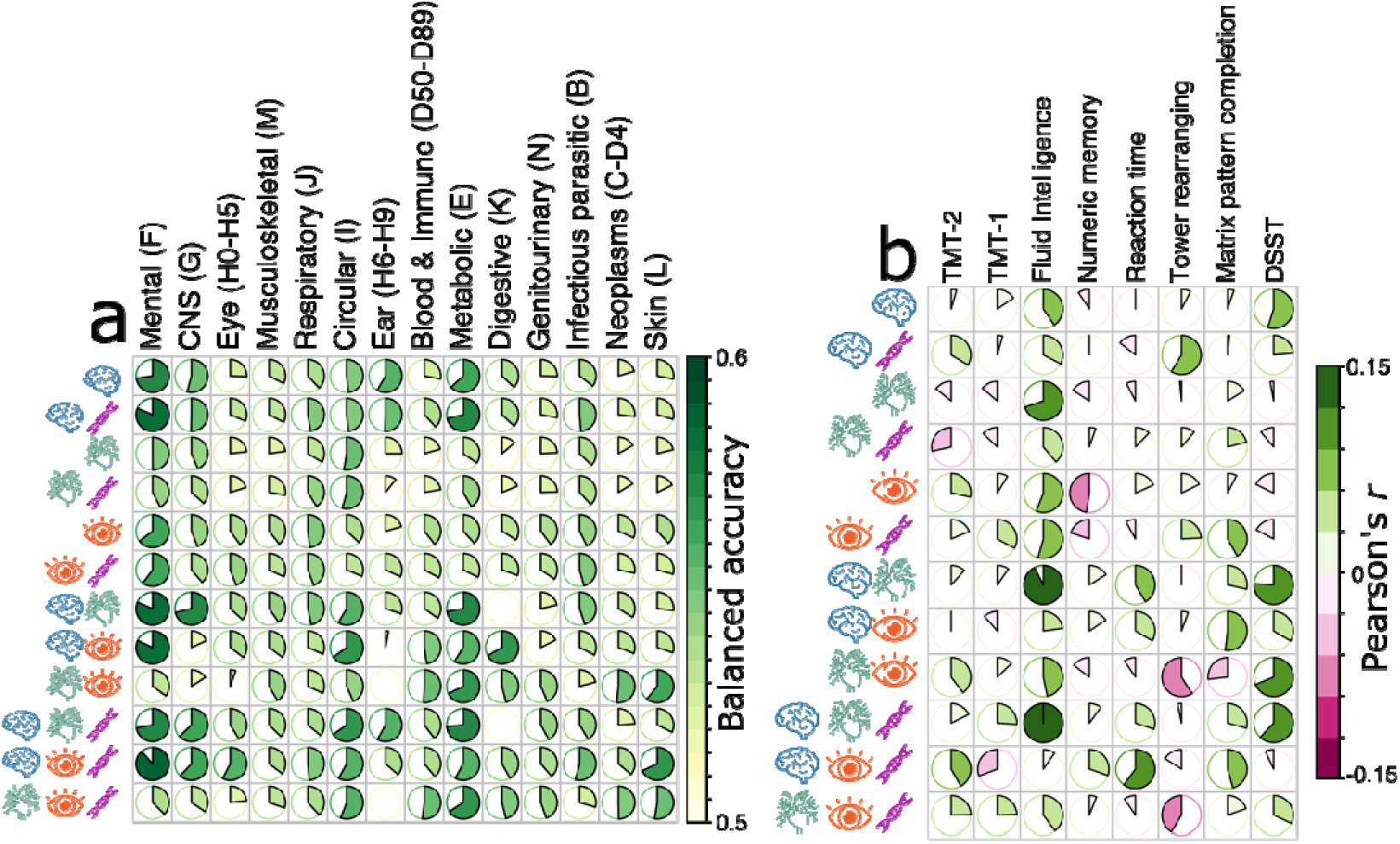
Prediction ability of the 2003 brain PSCs and 82 heart IDPs for predicting 14 systemic diseases and 8 cognitive scores **a**) The classification for the 14 ICD-based disease categories was assessed using brain, heart, and eye features and their respective PRSs via support vector machines in a nested cross-validation (CV) procedure (i.e., training/validation/test datasets). We present the balanced accuracy from the CV results. **b**) Cognition prediction was assessed via support vector regression using brain, heart, and eye features, as well as their respective PRSs. Pearson’s *r* coefficient reflects the relationship between predicted values and the ground truth from the CV test datasets. Additional metrics, results using alternative machine learning methods, and outcomes from the independent test dataset can be found in **Supplementary eFile 56-58**. The size of the pie charts reflects the balanced accuracy achieved using specific feature sets. A more filled-in pie chart signifies higher accuracy.

Importantly, there was no evidence of overfitting when comparing the results obtained from the nested CV test datasets (*CV accuracy*, shown in **Fig. 8a**) to those derived from the independent test dataset (*Ind. accuracy*, detailed in **Supplementary eFile 56-58**). We refrained from statistically comparing the classification performance between machine learning models due to the complexities involved within a complex nested cross-validation setup^83^ and the liberty in standard statistical tests like a two-sample t-test. Instead, we present the mean and standard deviation of the CV results. Notably, neural networks exhibited comparable results to support vector machines in the independent test dataset (**Supplementary eFile 59** for brain and heart features). **Supplementary eFigure 25** compares our brain PSCs with conventional atlas-based brain IDPs (i.e., the MUSE ROIs), demonstrating data-driven PSCs with higher balanced accuracy than MUSE ROIs in certain disease categories, especially for mental and behavioral disorders.

Overall, the predictive capacity for predicting the 8 cognitive scores was low using the 2003 PSCs, 82 heart IDPs, 84 eye IDPs, and their derived PRSs regarding Pearson’s *r* (<0.15). When combining the features across the two organs, using support vector regression, the prediction performance substantially increased for some cognitive scores, especially for fluid intelligence and DSST. Brain PSCs achieved better prediction results for the tower rearranging test than the other two organs (**Fig. 8b**). Additionally, integrating age and sex as supplementary features notably improved the prediction performance (**Supplementary eFigure 26**). Detailed metrics regarding the predictions using different features and machine learning models are shown in **Supplementary eFile 60-63**.

## Discussion

This study established a comprehensive brain-heart-eye axis by integrating multi-omics and multi-organ data from UKBB, BLSA, FinnGen, and PGC. We examined this axis across multiple scales to reveal its intricate phenotypic landscape, proteomic map, and genetic architecture. Our findings highlighted both within-organ specificity and cross-organ interactions among the three organs, further dissecting the association toward causality in chronic diseases. Additionally, our machine learning-based predictions underscored the importance of incorporating features from multiple organs and omics data, promoting a multi-organ approach for modeling human aging and disease.

### The phenotypic landscape of the brain-heart-eye axis highlights relevant imaging biomarkers

The brain-eye PWAS highlights the central visual pathway marked by distinct brain regions and ocular features, providing insights into the underlying neurobiological basis and potential new imaging biomarkers. The central visual pathway – linking the retina to the midbrain, thalamus, and the primary visual cortex – represents an intricate neural network that establishes a critical connection between the eye and the brain, orchestrating the transmission and processing of visual information^52^. Our PWAS results emphasize the pivotal contribution of ganglion cells (GcplR and GcplL of the eye IDPs) within the brain-eye nexus. These cells in the retina possess extensive axons that project into the brain, forming the optic nerve. This initiates the primary pathway for transmitting visual information from the eye to the brain, extending through the retina, optic nerve, optic chiasm, and optic tract. The degeneration of retinal ganglion cells has been associated with various ocular pathologies, encompassing glaucoma, hereditary optic neuropathies, ischaemic optic neuropathies, and demyelinating diseases^85,86^, as well as diseases related to the CNS^87^. The central visual pathway in the brain progresses to the thalamus’s LGN before reaching the occipital lobe’s primary visual cortex. This progression is supported by the distinct patterns observed in PSCs that spatially encompass these regions (closest-matching PSCs for thalamus: https://labs-laboratory.com/bridgeport/music/thalamus). Prior research has probed into glaucoma pathology, focusing on the retina and optic nerve heads, showcasing degenerative brain alterations encompassing the intracranial optic nerve, lateral geniculate nucleus, and visual cortex^88,89^. Multiple MRI studies have also investigated the correlation between brain structure alterations and eye conditions^90,91^. For example, in patients with neurodegenerative conditions, such as AD^92–95^, the eye serves as a portal to assess brain health.

The brain-heart PWAS emphasizes related brain areas, such as PSCs for deep subcortical structures, the prefrontal and insular cortex, alongside heart features, such as the myocardial mass and wall thickness of the left ventricle (LVM), and the end-diastolic volume of the right ventricle (RVEDV). This supports the “neuro-cardiac axis^96^”, which refers to the intricate connection and communication pathways between the brain and the heart. This axis involves the bidirectional interaction of neurological signals and cardiac responses, influencing each other’s function and regulation. It also encompasses various brain regions responsible for autonomic nervous system control, emotional processing, and cognitive functions, which affect cardiac activity, rhythm, and cardiovascular health. For example, the limbic system, including the emotional coding center^97^, amygdala (closest-matching PSCs: https://labs-laboratory.com/bridgeport/music/amygdala), and the cerebral cortex, contribute to autonomic nervous system modulation, particularly in response to emotions, stress, and cognitive processes^98^. More broadly, the pathophysiological interplays of the nervous and cardiovascular systems, evidenced by constant communication between the heart and the brain, have proved invaluable to interdisciplinary fields of neurological and cardiac diseases^49,99–101^. Consistent with a previous study using conventional brain IDPs^8^, which highlighted numerous associations between subcortical structures and global eye wall thickness, our PSC PWAS validates this pattern, which reveals the most prominent brain PSC and heart IDP associations within subcortical PSCs in a purely data-driven fashion.

In our PWAS analysis, the heart IDPs and eye IDPs showed weaker associations than the other two pairs of organs. This can be attributed to several factors. First, the brain shows a strong functional specialization to the other two organs. The brain has direct regulatory control over both the heart and the eye through neural pathways. The central autonomic network, as evidenced by our results, directly influences heart function, while the brain’s visual processing areas are closely linked to eye function. This direct control and specialization may lead to stronger associations. Secondly, this also leads to the fact that physiological pathways connecting the brain to the heart and eye are more direct than those connecting the heart and eye. Another possibility is that many neurological conditions can manifest with both cardiovascular and ocular symptoms, creating stronger brain-heart and brain-eye associations. However, a recent review^102^ also highlighted that identifying retinal microvascular disease and retinal biomarkers could offer valuable insights into the progression of microvascular disease in other systemic vascular regions over time.

### The proteomics map of the brain-heart-eye axis depicts organ-specific and cross-organ interconnections

Our ProWAS offers additional evidence supporting the brain-heart-eye axis at the protein level. We observed predominantly organ-specific protein expression profiles within individual organs, while notable cross-organ interactions were also obvious. We further discuss the potential implications of the ProWAS results by exemplifying significant proteins over-expressed in tissues from respective organs.

In the brain ProWAS, the MOG protein, encoded by the *MOG* gene located on chromosome 6p22.1 (**Supplementary eFigure 27a**), represents myelin oligodendrocyte glycoprotein. This protein primarily facilitates homophilic cell-cell adhesion. As a minor component of the myelin sheath, MOG plays a role in the completion and maintenance of the myelin sheath and cell-cell communication. This protein was significantly linked to many PSCs, including the C32_1 (*N*=4235; *β*=89.88±9.34; −log_10_(P-value)=20.96), encompassing the deep subcortical structures (**Supplementary eFigure 27b**). For organ/tissue-specific expression analysis, this protein showed high expression in several brain tissues and gastrointestinal tracts like the small intestine (**Supplementary eFigure 27c**). To further depict specific brain regions showing high expression of this protein, we calculated the normalized RNA expression levels (nTPM) for 13 brain tissues. White matter tissue showed the highest nTPM (**Supplementary eFigure 27d**). This aligns with our understanding that white matter is abundant in oligodendrocytes, which are essential for myelination. Additionally, this protein is classified in cluster 9, ‘Oligodendrocytes – Myelination’, according to protein expression clustering analysis (**Supplementary eFigure 27e**), where it shows a high correlation with myelin basic protein (MBP) (*r*=0.9982). Moreover, this protein also exhibits cell type-specific enrichment in cardiomyocytes from heart muscle (**Supplementary eFigure 27f**). Cancer-type specific enrichment showed 5.9 FPKM in glioma among 17 cancer types using RNA-seq data from the TCGA dataset^103^ (**Supplementary eFigure 27g**).

Another example is the PLTP (phospholipid transfer protein) protein (**Supplementary eFigure 28a**), which was significantly linked to the average thickness of the INL/RPE layer (InlRpeL: https://labs-laboratory.com/medicine/average_inlrpe_thickness_left_f28536_0_0), and showed a high expression in various human tissues/organs, including the brain, eye, and heart (**Supplementary eFigure 28b-c**). This protein mainly facilitates the transfer of phospholipids and free cholesterol from triglyceride-rich lipoproteins (such as low-density lipoproteins or LDL and very low-density lipoproteins or VLDL) into high-density lipoproteins and also enables the exchange of phospholipids among triglyceride-rich lipoproteins. Tissue cell-type-specific enrichment analysis revealed a significant presence of macrophages across various tissues (**Supplementary eFigure 28d**), which plays a pivotal role in the innate immune response.

Single-cell type analysis in the retina showed substantial expression of this protein in a type of retinal glial cells (Müller glial cell; **Supplementary eFigure 28e**). Our protein-level analyses exemplify complex implications for the PLTP protein in specific retinal mechanisms. These findings suggest potential links between PLTP function in the retina and its roles in other brain regions and peripheral organs.

In the heart ProWAS, we highlight the TGFA (transforming growth factor alpha; **Supplementary eFigure 29a**) protein, which showed a high expression in tissues from various organs (**Supplementary eFigure 29b-c**). TGFA is a mitogenic polypeptide that exhibits binding affinity for the epidermal growth factor receptor. TGFA demonstrates synergistic activity with transforming growth factor beta (TGF-β) in promoting anchorage-independent cell proliferation. The TGFA protein was significantly linked to multiple heart IDPs, such as the maximum volume of the RA (RAV_max: https://labs-laboratory.com/medicine/ra_maximum_volume_f24114_2_0). Regional brain expression analysis revealed that white matter exhibited the highest enrichment of this protein, with an nTPM value of 53.2 (**Supplementary eFigure. 29d**).

### Genetic evidence of the brain-heart-eye axis dissects causal relationships within a multi-organ network

Our GWAS and post-GWAS analyses provide comprehensive genetic evidence that may shape this brain-heart-eye axis’s observed phenotypic landscape and proteomics map.

Our GWAS uncovered shared genetic variants commonly associated with brain PSCs, eye IDPs, and heart IDPs. These findings have several important implications and suggest potential shared biological pathways. First, the joint associations between genetic variants and imaging features across the brain, eye, and heart emphasize cross-organ pleiotropic effects^104^. Second, chronic diseases affecting these three organs might share similar pathological and biological pathways. For instance, complex diseases like Alzheimer’s, cardiovascular diseases, and certain ocular conditions often have multifactorial causes. Identifying common genetic variants helps elucidate the genetic contributions to these diseases and their effects on different organs. Third, cross-organ causality may also contribute to the shared genetic variants, supported by our Mendelian randomization and genetic colocalization analyses (**Fig. 7c-e**).

We calculated several key genetic parameters to depict the genetic architecture of the three organs. We observed that SNP-based heritability estimates were higher for brain PSCs and eye IDPs compared to heart IDPs, with a P-value of 0.10 when comparing brain PSCs to heart IDPs and a P-value of 4.12×10^−19^ for eye IDPs versus heart IDPs. This aligns with our previous study, where we assessed the heritability of biological age across the three organs using machine learning techniques^10^. In that study, we examined the underlying factors contributing to this phenomenon, such as allele frequency, sample sizes, and the quality of the imaging features. We then compared the natural selection signature (*S*) and polygenicity (*Pi*) across the three organs. Our results showed a widespread signature of negative selection of the three organs, with the eye IDPs showing the largest magnitude (P-value<0.001) compared to the other two organs (**Fig. 5 d-f**). These findings are consistent with previous literature^105^, where Zeng et al. found signatures of negative selection in 28 human complex traits. The retina is a highly conserved structure across different species, having been subject to significant purifying selection pressures to preserve its essential role in vision. This strong selection pressure ensures the removal of deleterious mutations, maintaining the retina’s functional integrity over evolutionary time. For instance, a prior multi-omics analysis study identified 178 ultrarare variants within 84 ultraconserved non-coding regions (UCRs) linked to 29 disease genes during retinal development^106^; the negative selective pressure on these regions is much stronger than on protein-coding regions. In contrast, the brain’s adaptability and plasticity allow it to meet diverse functional demands, potentially reducing the selection pressure on specific brain traits. This complexity often involves a myriad of genes with small individual effects, which can diffuse the impact of purifying selection typically seen in traits with fewer (i.e., polygenicity), likely more significant genetic determinants. Our polygenicity estimates support this, where the eye IDPs obtained the lowest *Pi* estimates (P-value<1×10^−10^, **Fig. 5 g-i**).

The tissue-specific heritability enrichment results emphasize notable heritability enrichment within organ-specific tissues (**Fig. 5j-l**). For example, the heritability enrichment of ganglia, which also aggregates outside the brain – like those in the retina^107^ and cardiac ganglia^108^ – offers cellular evidence supporting this inter-organ communication. Additionally, we noticed a greater significance in the enrichment analyses (indicated by significant P-values) when employing chromatin data compared to gene expression data. This observation aligns with the original heritability enrichment analysis carried out by Finucane et al.^109^, wherein the authors identified more significant enrichment signals in chromatin data than gene expression data in disease-relevant tissues and cell types. The genetic correlation between the PSCs/IDPs of each pair of organs largely mirrors their phenotypic associations, supporting the long-standing Cheverud’s Conjecture^72^.

Finally, Mendelian randomization analyses established 6 causal networks between the three organs (**Fig. 7f-h**). It highlighted potential causal relationships between imaging-derived endophenotypes^110^ (PSCs/IDPs) and various chronic disease endpoints across different organs. For example, we identified multiple heart IDPs that were causally linked to three brain diseases: depression, migraine, and AD. In a previous study, Zhao et al. also found similar causal evidence between psychiatric disorders and heart IDPs, but their analysis was limited to pre-selected, well-powered traits from multiple publicly available resources. Our study extends previous research^8,58^ by implementing bi-directional causal analysis across the brain, heart, and eye. This approach revealed additional insights, including causal links between several heart IDPs and ocular diseases, such as glaucoma.

### Multi-organ features improve prediction for systemic disease categories and cognition

We demonstrated the clinical potential of these brain PSCs, heart and eye IDPs, and their respective PRSs using machine learning for precision diagnostics. Notably, integrating multi-scale data from different organs and omics enhanced predictive power.

Previous research has shown the great potential of these *in vivo* imaging biomarkers for advancing precision medicine. However, one limitation of these machine learning methods emerges from models trained solely on features derived from individual organ systems or omics data. This approach might inadequately capture the multifaceted pathological effects of underlying conditions, especially considering the co-occurring comorbidities across different systems, such as the cardiovascular contributions for epilepsy^111^ and dementia^112^. Importantly, while the individual impact of brain PSCs, heart IDPs, and their PRSs is limited in predicting systemic disease categories and cognitive scores, their supplementary power beyond basic demographics like age and sex, compared to features from a single organ, emphasizes the necessity for a multi-organ approach in future research. Another challenge in applying machine learning for precision medicine is the reproducibility crisis^113,114^, as demonstrated in our prior research on AD classification using convolutional neural networks^22^. Consequently, open science, permitting the community to examine the model, source code, and reuse the GWAS summary statistics, is crucial to advancing the field toward potential clinical translation. To address this, we made all our resources and results publicly available at the MEDICINE knowledge portal: https://labs-laboratory.com/medicine/ and the BRIDGEPORT knowledge portal: https://labs-laboratory.com/bridgeport.

### Integrating machine learning and artificial intelligence in multi-organ research

Machine learning and artificial intelligence have been widely applied in biomedical research. For example, the foundational model proposed by Zhou et al.^39^ may potentially enhance its capabilities by leveraging multi-organ imaging data during training. Recent studies^115,9^ focused on determining the biological age of nine distinct human organ systems and identifying the clinical and genetic factors that underlie the multifaceted aging process. These findings underscore the clinical promise of utilizing these AI-derived endophenotypes to gauge and understand the collective health of multiple organs. Machine learning and artificial intelligence have gained significant attention due to several factors. First, they are adept at handling high-dimensional biomedical data, such as brain MRI and genetics. They can distill the complex data into lower-dimensional yet clinically rich imaging and genetic signatures. Second, these data-driven models are considered more clinically and biologically relevant because they can capture underlying neurobiological effects related to aging and diseases. Additionally, integrating disease-related information from multiple modalities, such as imaging and genetics^116^, may enhance the power of discoveries in subsequent genetic discoveries.

### Limitation

Our study presents several limitations. Primarily, our genetic analysis concentrated solely on common genetic variants, prompting future inquiries into the potential impact of rare variants within this brain-heart-eye axis. Additionally, our GWAS analyses were predominantly based on participants of European ancestry. Future research efforts should encompass underrepresented racial and ethnic groups and disease-specific populations. Additionally, future research should investigate the impact of environmental factors and gene-environment interactions on the phenotypic variance of the three organs. Furthermore, the prevalence of missing proteomics data presents significant challenges for implementing multivariate AI/ML approaches. Future research will address this issue through improved data collection strategies and the development of advanced imputation methodologies^117^ for missing data imputation. Finally, the multi-scale brain PSCs across different scales are interconnected, even though our previous work^1^ demonstrated that integrating cross-scale features enhances disease classification. Future methodological advancements could focus on conditioning model training across scales to ensure that the derived PSCs are more independent.

### Outlook

In summary, this study integrates machine learning, imaging genetics, and proteomics to map the brain-heart axis across multiple scales. We expect these findings to broaden future research avenues, encouraging the adoption of machine learning techniques in multi-organ, multi-omics studies for precision medicine.

## Methods

### Method 1: The MULTI consortium

The MULTI consortium is an ongoing initiative to integrate and consolidate multi-organ data, such as brain and heart MRI and eye OCT, with multi-omics data at individual and summary levels, including imaging, genetics, and proteomics. Building on existing consortia and studies, such as those listed below, MULTI aims to curate and harmonize the data to model human aging and disease across the lifespan.

### UK Biobank

UKBB^42^ is a population-based research initiative comprising around 500,000 individuals gathered from the United Kingdom between 2006 and 2010. Ethical approval for the UKBB study has been secured, and information about the ethics committee can be found here: https://www.ukbiobank.ac.uk/learn-more-about-uk-biobank/governance/ethics-advisory-committee.

This study collectively analyzed 39,567 brain MRI scans (referred to as the *brain population*), 39,676 heart MRI scans (representing the *heart populations*, varying from 33,866 to 39,286 per specific IDP), and 64,316 eye OCT images (i.e., the *eye populations* varied from 15,997 to 61,732 per IDP) at baseline. The combined sample size across the three organ populations totaled 104,509. The T1-weighted MRI data underwent processing at the University of Pennsylvania, employing the MuSIC atlas to produce the 2003 brain PSCs^1^. Concurrently, the 82 heart IDPs were obtained directly from the UKBB website and derived from a prior study^2^ (Category ID: 157). Meanwhile, the 84 eye IDPs, derived from OCT imaging, were directly downloaded from the UKBB website and returned by previous studies^3–5^ (Category ID: 100079). The sample sizes employed in the brain-heart PWAS, exploring overlapping populations between brain PSCs and heart IDPs, ranged from 21,948 to 23,548 individuals, depending on the analyzed features. Of note, the brain and heart MRIs were scanned at the same visit/session. The sample sizes for the overlapping populations between the brain PSCs and eye IDPs used in PWAS varied from 1284 to 4472 individuals. The small sample sizes were because the brain MRI and eye OCT were scanned at different time/session points. For comprehensive information, including the complete list of phenotypes and their respective sample sizes, refer to **Supplementary eTable 1**.

For the genetic data, we conducted a quality check on the imputed genotype data^42^ for the entire UKBB population (approximately ~500k individuals). Subsequently, we merged the processed data with the *eye & heart population* for genetic analyses; the genetic quality check was performed on the brain population for the brain PSCs, as indicated in our previous study^1^.

Refer to **Method 5** for details. Our primary focus was on populations of European ancestry, with non-European ancestry populations included in sensitivity check analyses. The preprocessing for the proteomics data is detailed below.

### Baltimore Longitudinal Study of Aging

BLSA^45,54^ data (https://www.blsa.nih.gov/) were used to replicate the brain ProWAS results from the UKBB study. We finally included measurements of 7268 plasma proteins from 924 participants quantified with the SomaScan v4.1 platform after quality checks and merging with the imaging populations. Age (years), sex (male/female), race (white/non-white), and education level (years) were defined based on participant self-reports.

### FinnGen

The FinnGen^46^ study investigates combined genetic information alongside health registry data to unravel the origins and mechanisms behind various disease endpoints. Its primary emphasis lies in understanding the genetic foundations of diseases prevalent in the Finnish population, surpassing 500,000 individuals. This is accomplished through extensive GWAS and thorough analysis of vast genomic data in collaboration with multiple research entities. For the benefit of research, FinnGen generously made their GWAS findings accessible to the wider scientific community (https://www.finngen.fi/en/access_results). This research utilized the publicly released GWAS summary statistics (version R9), which became available on May 11, 2022, after harmonization by the consortium. No individual data were used in the current study.

FinnGen published the R9 version of GWAS summary statistics via REGENIE software (v2.2.4)^118^, covering 2272 disease endpoints, including 2269 binary traits and 3 quantitative traits. The GWAS model encompassed covariates like age, sex, the initial 10 genetic principal components, and the genotyping batch. Genotype imputation was referenced on the population-specific SISu v4.0 panel. Specifically, we included GWAS summary statistics for 45 heart, 32 eye diseases, and 41 brain diseases in our analyses.

### Psychiatric Genomics Consortium

PGC^47^ is an international collaboration of researchers studying the genetic basis of psychiatric disorders. PGC aims to identify and understand the genetic factors contributing to various psychiatric disorders such as schizophrenia, bipolar disorder, major depressive disorder, and others. The GWAS summary statistics were acquired from the PGC website (https://pgc.unc.edu/for-researchers/download-results/), underwent quality checks, and were harmonized to ensure seamless integration into our analysis. No individual data were used from PGC. Each study detailed its specific GWAS models and methodologies, and the consortium consolidated the release of GWAS summary statistics derived from individual studies. In the current study, we included 4 brain diseases.

### Method 2: Imaging analyses

#### (a) Patterns of structural covariance of the brain

Our earlier work^1^ applied the sopNMF method to a large-scale cohort of brain MRIs (*N*=50,699). This resulted in 2003 multi-scale brain PSCs, wherein the scale C ranged from 32 to 1024, expanding exponentially by a factor of 2. Of note, 13 PSCs from C1024 vanished during modeling, resulting in 2003 PSCs.

PSCs represent data-driven structural networks that encapsulate coordinated neuroanatomical changes in brain morphology stemming from a mixture of normal aging, pathology, and unmodeled factors, such as environmental and genetic influences.

Mathematically, the sopNMF algorithm is a deep learning-like stochastic approximation constructed and extended based on opNMF^44,119^. Consider a dataset comprising *n* MR images, each containing *d* voxels. The imaging data are represented as a matrix ***X***, where each column corresponds to a flattened image: 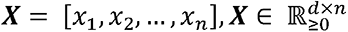. The sopNMF algorithm factorizes ***X*** into two low-rank matrices 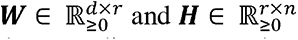, subject to the constraints of *i*) non-negativity and *ii*) column-wise orthonormality. More mathematical details are presented in **Supplementary eMethod 2** and the original references^1,44,119^.

#### (b) Imaging-derived phenotypes of the heart

The 82 heart IDPs were directly downloaded from the UKBB website (Category ID: 157). Bai et al.^2^ studied a broad spectrum of structural and functional characteristics for the heart and aorta, measured through heart MRI data sourced from UKBB, employing an automated analysis pipeline based on machine learning. They investigated the correlations of these heart IDPs with factors such as sex, age, key cardiovascular risk elements, and other non-imaging traits. As demonstrated in our present study, this exploration unveils new research avenues for scrutinizing disease mechanisms and developing image-based biomarkers across diverse organ systems assessed through advanced imaging techniques. For the 82 heart IDPs included in our analyses, we further categorized them into 6 different IDP groups for visualization purposes, as shown in **Supplementary eTable 1**.

#### (c) Imaging-derived phenotypes of the eye

For the 88 eye IDPs, we downloaded the derived OCT measurements from the UKBB website (Category ID: 100079) and used the imaging quality scores (Field ID: 28552 and 28553) for quality check. We excluded 12,044 individuals whose scores were lower than 45 (Category ID: 100116). Additionally, we restricted our analysis to IDPs with sample sizes surpassing 10,000 within the European ancestry, excluding four IDPs.

OCT imaging is an advanced, non-invasive technology that generates three-dimensional images of the macula, crucial for detailed central vision. Abnormalities in macular thickness and morphology captured by OCT imaging are sensitive biomarkers of diabetic retinopathy, age-related macular degeneration, glaucoma, sleeping, and various neurodegenerative diseases^120,121^. Three prior studies^3–5^ processed the OCT images to derive the 84 eye IDPs used in our analyses, and the results were subsequently returned to UKBB, making them accessible to the community. Ko et al.^4^ and Patel et al.^3^ from the UKBB Eyes and Vision Consortium (https://www.ukbiobankeyeconsortium.org.uk/; Return code: 1873 and 1875) analyzed OCT images from over 60,000 individuals in the UKBB. They derived variables related to the thickness of the retinal pigment epithelium (excluding outliers and individuals with diseases affecting macular thickness). They associated the thickness measures with age, myopia, ethnicity, smoking, and intraocular pressure. A subsequent investigation by Han et al.^5^ extracted optic nerve head morphology measures and performed a GWAS on these additional eye IDPs.

For the 84 eye IDPs included in our analyses, we further categorized them into 11 different IDP groups for visualization purposes (**Supplementary eTable 1**).

### Method 3: Phenome-wide associations between the three organs

#### (a) Primary PWAS

We performed three sets of primary PWAS that correlated *i*) the 2003 brain PSCs with the 82 heart IDPs, *ii*) the 2003 brain PSCs with the 84 eye IDPs, and *iii*) the 84 eye IDPs and 82 heart IDPs, employing a linear regression model where each organ-specific feature served as the dependent variable while the other functioned as the independent variable.

For the brain-heart PWAS, we accounted for various covariates, including age (Field ID: 21003), sex (Field ID: 31), brain positioning in the scanner (lateral, transverse, and longitudinal; Field ID: 25756-25758), head motion (Field ID: 25741), intracranial volume, body weight (Field ID: 21002), height (Field ID: 50), waist circumference (Field ID: 48), BMI (Field ID: 23104), smoking status (Field ID: 20116), diastolic (Field ID: 4079), and systolic (Field ID: 4080) blood pressure, assessment center (Field ID: 54), body surface area (Field ID: 22427), average heart rate (Field ID: 22426), along with the first 40 genetic principal components. For the brain-eye PWAS, we controlled for age (Field ID:21003 for both brain and eye assessment), sex (Field ID: 31), brain position in the scanner (lateral, transverse, and longitudinal; Field ID: 25756-25758), head motion (Field ID: 25741), intracranial volume, body weight (Field ID: 21002 for both brain and eye assessment), height (Field ID: 50 for both brain and eye assessment), waist circumference (Field ID: 48 for both brain and eye assessment), and first 40 genetic principal components as covariates. For the heart-eye PWAS, we included covariates for age (Field ID:21003 for both heart and eye assessment), sex (Field ID: 31), body weight (Field ID: 21002 for both heart and eye assessment), height (Field ID: 50 for both heart and eye assessment), waist circumference (Field ID: 48 for both heart and eye assessment), and first 40 genetic principal components, smoking status (Field ID: 20116), diastolic (Field ID: 4079), and systolic (Field ID: 4080) blood pressure, assessment center (Field ID: 54), and BMI (Field ID: 23104).

We conducted two sensitivity analyses to check the robustness of our main PWAS: *i*) sex-specific PWAS for males and females and *ii*) split-sample PWAS by randomly dividing the entire population into two groups, ensuring that there were no significant differences in sex and age between the two splits.

#### (b) Secondary PWAS

A secondary PWAS was conducted to link the brain PSCs, heart, and eye IDPs with other 117 phenotypes (excluding the heart and eye IDPs used here) accessible in our downloaded UK Biobank database (**Supplementary eFile 4** and **5**). The same linear regression model mentioned above was used. We explicitly excluded the multimodal brain IDPs^26^ (Category code: 100) from this PWAS to prevent circular bias with our brain PSCs^122^.

### Method 4: Proteome-wide associations of the brain PSCs, heart, and eye IDPs

#### (a) Proteome-wide associations

We performed three sets of ProWAS that correlated *i*) the 2003 brain PSCs, *ii*) the 82 heart IDPs, and *iii*) 84 eye IDPs with 2923 unique proteins (10,018<*N*<39,489) using the Olink platform. The original data were analyzed and made available to the community by the UK Biobank Pharma Proteomics Project. The initial quality check was detailed in the original work^48^; we performed additional quality check steps as below. We focused our analysis on the first instance of the proteomics data (“instance”=0). Subsequently, we merged the Olink files containing coding information, batch numbers, assay details, and limit of detection (LOD) data (Category ID: 1839) to match the ID of the proteomics dataset. We eliminated Normalized Protein eXpression (NPX) values below the protein-specific LOD. Furthermore, we restricted our analysis to proteins with sample sizes exceeding 10,000. Of note, we observed a high prevalence of missing values across individuals for the 2923 proteins, posing challenges for employing multivariate machine learning models.

For the brain ProWAS, we accounted for various covariates, including age (Field ID: 21003), sex (Field ID: 31), brain positioning in the scanner (lateral, transverse, and longitudinal; Field ID: 25756-25758), head motion (Field ID: 25741), intracranial volume, body weight (Field ID: 21002), height (Field ID: 50), waist circumference (Field ID: 48), BMI (Field ID: 23104), assessment center (Field ID: 54), protein batch number (Category ID: 1839), limit of detection (LOD; Category ID: 1839), along with the first 40 genetic principal components. For the eye ProWAS, we controlled for age (Field ID:21003 for eye assessment instance), sex (Field ID: 31), body weight (Field ID: 21002 for eye assessment), height (Field ID: 50 for eye assessment), waist circumference (Field ID: 48 for eye assessment), protein batch number (Category ID: 1839), limit of detection (LOD; Category ID: 1839), and first 40 genetic principal components as covariates. For the heart ProWAS, we included covariates for age (Field ID:21003), sex (Field ID: 31), body weight (Field ID: 21002), height (Field ID: 50), waist circumference (Field ID: 48), and first 40 genetic principal components, smoking status (Field ID: 20116), diastolic (Field ID: 4079), systolic (Field ID: 4080) blood pressure, assessment center (Field ID: 54), and BMI (Field ID: 23104), protein batch number (Category ID: 1839), and LOD (Category ID: 1839). Multiple comparisons were performed using Bonferroni corrections based on the number of proteins and the number of PSC or IDP for each organ. We also used the time interval as an alternative covariate in the model for image data from organs that were not collected at the same visit as the proteomics data.

#### (b) Tissue/organ-specific map of the expression of the human proteome

To integrate our ProWAS results within the framework of multi-organ connections, we annotated the tissue/organ-specific expression patterns of significant proteins using the Human Protein Atlas^123^. This integrated the RNA and protein level data from multiple sources, including the HPA, GTEx^124^, and FANTOM5^125^ datasets, to comprehensively assess tissue “over-expression” profiles for the significant proteins. The protein data encompasses 15,303 genes for which antibodies are available. The RNA expression data is obtained from deep sequencing of RNA (RNA-seq) across different tissue types. The methodology determining the expression of the protein is detailed in the original publication^123^. Importantly, proteins are then simultaneously over-expressed in various tissues or organs (i.e., lack of organ-specificity). Our main objective was to determine if the tested protein exhibited expression in the brain, eye, and heart. If it was not over-expressed in any of these three organs, we highlighted its expression in other tissues with the highest evidence of over-expression.

### Method 5: Genetic analyses

We used the imputed genotype data for all genetic analyses, and our quality check pipeline resulted in 33,541 participants (8,469,833 SNPs passing quality check) for the *brain population*, 33,743 participants (6,477,810 SNPs passing quality check) for the *heart population*, and 48,016 participants (6,477,810 SNPs passing quality check) for the *eye population* with European ancestry. We summarize our genetic quality check steps. First, we excluded related individuals (up to 2^nd^-degree) from the full UKBB sample using the KING software for family relationship inference.^126^ Importantly, in the GWAS for heart and eye IDPs using fastGWA^57^, we skipped this step, as the linear mixed model inherently addresses population stratification, encompassing additional cryptic population stratification factors. We then removed duplicated variants from all 22 autosomal chromosomes. Individuals whose genetically identified sex did not match their self-acknowledged sex were removed. Other excluding criteria were: *i*) individuals with more than 3% of missing genotypes; *ii*) variants with minor allele frequency (MAF) of less than 1%; *iii*) variants with larger than 3% missing genotyping rate; *iv*) variants that failed the Hardy-Weinberg test at 1×10^−10^. To adjust for population stratification,^127^ we derived the first 40 genetic principle components using the FlashPCA software^128^. Details of the genetic quality check protocol are described elsewhere^1,9,10,129^. The quality check for the brain PSC GWAS was conducted within the brain imaging genetic populations^1^, whereas the heart and eye IDP GWAS was carried out across the entire 500k UKBB population, leading to the different numbers of valid SNPs.

#### (a) GWAS

##### Brain PSC GWAS

In our prior study^1^, we performed linear regression using Plink^130^ for each brain PSC within a subset of the UKBB brain population (discovery set; *N*=18,052; European ancestry). Following this, we replicated the findings in another dataset (replication set; *N*=15,243). This study presents GWASs from the combined discovery and replication sets (*N*=33,541 European ancestry). The brain PSC GWAS controlled for confounding factors, including age (Field ID:21003), age-squared, sex (Field ID:31), age x sex interaction, age-squared x sex interaction, the first 40 genetic principal components, and total intracranial volume, guided by earlier brain imaging GWASs^26,61^. We employed a stringent genome-wide P-value threshold (5×10^−8^/2003) using Bonferroni correction based on the number of PSCs (*N*=2003) to ensure stringent statistical rigor.

##### Heart IDP GWAS

We applied a linear mixed model regression to the European ancestry populations using fastGWA^57^ implemented in GCTA^60^. The model included age (Field ID: 21003), age-squared, sex (Field ID: 31), age x sex interaction, age-squared x sex interaction, the first 40 genetic principal components, body weight (Field ID: 21002), height (Field ID: 50), and waist circumference (Field ID: 48), BMI (Field ID: 23104), smoking status (Field ID: 20116), assessment center (Field ID: 54), heart rate (Field ID: 12673), diastolic (Field ID: 12675), and systolic (Field ID: 12674) blood pressure, peripheral pulse pressure (Field ID: 12676), central pulse pressure (Field ID: 12678), body surface area (Field ID: 22427), average heart rate (Field ID: 22426), as covariates. Likewise, based on the number of heart IDPs, we applied the Bonferroni correction on top of the genome-wide significant threshold (5×10^−8^/82).

##### Eye IDP GWAS

We applied a linear mixed model regression to the European ancestry populations using fastGWA^57^ (12,120<*N*<45,897 across the 84 eye IDPs). The model included age (Field ID: 21003), age-squared, sex (Field ID: 31), age x sex interaction, age-squared x sex interaction, the first 40 genetic principal components, body weight (Field ID: 21002), height (Field ID: 50), and waist circumference (Field ID: 48) as covariates. Similarly, based on the number of eye IDPs, we applied the Bonferroni correction on top of the genome-wide significant threshold (5×10^−8^/84).

We scrutinized the robustness of the brain PSC GWAS in our previous sutdy^1^. Here, we assessed the reliability of our heart and eye IDP GWAS results through several sensitivity analyses. These checks included: *i*) a split-sample GWAS that randomly divided the entire heart population into two groups with no significant differences in sex and age, *ii*) sex-stratified GWAS conducted separately for males and females, *iii*) a non-European GWAS to gauge the generalizability of GWAS signals identified in populations of European ancestry, *iv*) PLINK linear model GWAS to compare genetic signals with fastGWA, and *v*) validation against the prior heart and eye IDP GWAS by Zhao et al.^8,58^, which used data from UKBB with different sample sizes and valid SNP after quality check.

##### Annotation of genomic loci

For heart and eye IDP GWASs, genomic loci were annotated using FUMA^131^. For genomic loci annotation, FUMA initially identified lead SNPs (correlation *r^2^*≤ 0.1, distance < 250 kilobases) and assigned them to non-overlapping genomic loci. The lead SNP with the lowest P-value (i.e., the top lead SNP) represented the genomic locus. Further details on the definitions of top lead SNP, lead SNP, independent significant SNP, and candidate SNP can be found in **Supplementary eMethod 1**. To identify genomic loci in the brain PSC GWAS, we utilized PLINK with parameters for clumping that matched those of the FUMA online platform. This choice was made due to the resource constraints of the FUMA platform in handling tasks at this scale (*N*=2003), as described in our prior study^1^.

#### (b) PheWAS

We used the GWAS Atlas^59^ platform to conduct an online PheWAS look-up analysis for the top lead SNP within each genomic locus; linkage disequilibrium was fully considered in this case. To facilitate this, we developed an “in-house web crawler” designed to automate searches on the PheWAS webpage: https://atlas.ctglab.nl/PheWAS. The search threshold was set at a P-value of 5×10^−8^. The GWAS Atlas PheWAS categorized these traits into different broad domains/categories. This PheWAS was conducted on December 2, 2023.

#### (c) SNP-based heritability

We calculated the SNP-based heritability (*h^2^*) using the GCTA software, which employs raw individual genotype data to generate a genetic relationship matrix, addressing the “missing heritability” issue^60^. Additionally, we estimated *h^2^* using the LDSC^56^ software, leveraging only GWAS summary statistics derived from our brain PSC, heart, and eye IDP GWASs as an alternative approach. To this end, we used precomputed LD scores from the European ancestry in the 1000 Genomes dataset. Notably, disparities in SNP-based *h^2^* estimates were observed between the two software tools, which is in line with our own findings^10,12^ and those from other studies^26,61^. Consequently, we present the GCTA estimates in the main manuscript and the LDSC estimates in the supplement. Bonferroni correction based on the number of PSCs/IDPs was applied to denote statistical significance.

#### (d) Selection signature and polygenicity estimate

We used SBayesS^63^ to estimate two sets of parameters that unveil the genetic architecture of the PSCs and IDPs. SBayesS is an expanded approach capable of estimating essential parameters characterizing the genetic architecture of complex traits through a Bayesian mixed linear model^105^. This method only requires GWAS summary statistics of the SNPs and LD information from a reference sample. These parameters include polygenicity (*Pi*), and the relationship between minor allele frequency (MAF) and effect size (*S*). We used the software pre-computed sparse LD correlation matrix derived from the European ancestry by Zeng et al.^63^. More mathematical details can be found in the original paper from Zeng et al.^63^. We ran the *gctb* command^105^ using the argument *--sbayes S*, and left all other arguments by default.

#### (e) Tissue-specific partitioned heritability estimate

This analysis aims to comprehend the varying roles of distinct tissue types in contributing to the heritability of the brain PSCs and heart and eye IDPs. To achieve this, the partitioned heritability analysis through stratified LD score regression assesses the extent of heritability enrichment attributed to predefined and annotated genome regions and categories^64^. This analysis considers two sets of analyses considering different tissue types: *i*) 498 multi-tissue chromatin-based annotations from peaks from six epigenetic marks using data from ROADMAP^66^ and ENTEx^67^ and *ii*) 205 multi-tissue gene expression data estimate using data from GTEx V8^65^ and “Franke lab” dataset. Bonferroni correction is applied to all tested annotations and categories (0.05/498/205). Detailed methodologies for the stratified LD score regression are outlined in the original work^64^. LD scores and allele frequencies for European ancestry were acquired from a predefined version based on data from the 1000 Genomes project.

#### (f) Gene-drug-disease network

We defined a gene-drug-disease network by examining the enrichment of the significant genes linked to the 2003 brain PSCs, 82 heart IDPs, and 84 eye IDPs within specific drug categories from the DrugBank database^69^ using the GREP software^68^. First, the gene-level association test was performed using MAGMA^132^. Then, gene annotation was performed to map the SNPs (reference variant location from Phase 3 of 1,000 Genomes for European ancestry) to genes according to their physical positions. We then performed gene-level associations based on the SNP GWAS summary statistics to obtain gene-level P-values to define the Bonferroni-corrected significant genes linked to each PSC/IDP (P-value<0.05/18,761). Using these significant genes as input, GREP conducted Fisher’s exact tests to determine whether these genes were enriched in gene sets targeted by drugs within clinical indication categories for specific diseases or conditions (based on the ICD code). FDR correction was applied to account for multiple tests to signify significant gene-drug-disease relationships.

#### (g) Genetic correlation

We estimated the genetic correlation (*g_c_*) between each PSC-IDP pair utilizing the LDSC software. Precomputed LD scores from the 1000 Genomes of European ancestry were employed, maintaining default settings for other parameters in LDSC. It’s worth noting that LDSC corrects for sample overlap, ensuring an unbiased genetic correlation estimate^133^. Statistical significance was determined using Bonferroni correction, considering the number of PSCs or IDPs (*N*=2003, 84, or 82, whichever is the largest).

#### (h) Bayesian colocalization

We explored the genetic colocalization signals between pairwise PSC-IDP and IDP-IDP at each genomic locus defined by respective GWASs using the *coloc* package. Specifically, we employed the Fully Bayesian colocalization analysis^73^ utilizing Bayes Factors (*coloc.abf*). This method assesses the posterior probability (PP.H4.ABF: Approximate Bayes Factor) to evaluate hypothesis H4, indicating the presence of a single shared causal variant associated with both traits. To establish the significance of the H4 hypothesis, a threshold of PP.H4.ABF>0.8^73^ was set. All other parameters, such as the prior probability of p_12_, were maintained at their default values.

#### (i) Two-sample bidirectional Mendelian randomization

We postulated that the brain PSCs and heart and eye IDPs, acting as intermediate phenotypes (endophenotypes)^134^, might have causal connections to chronic disease endpoints across the three organs. To test this hypothesis, we constructed six bi-directional causal networks: *i*) *Brain2Heart*, *ii*) *Heart2Brain*, *iii*) *Brain2Eye*, *iv*) *Eye2Brain*, *v*) *Heart2Eye,* and *vi*) *Eye2Heart.* These networks used GWAS summary statistics from our analyses in the UKBB, the FinnGen^46^, and the PGC^47^ study for the brain, heart, and eye disease endpoints. The *Brain2Heart* causal network employed the 2003 brain PSCs from UKBB as exposure variables and the 45 heart diseases from FinnGen (R9) as outcome variables (disease code: I9). The *Heart2Brain* network investigated causality from the 82 heart IDPs from UKBB as exposure variables to 41 FinnGen brain diseases (disease code: F5 and G6) as outcome variables. The *Brain2Eye* causal network utilizes the 2003 brain PSCs from UKBB as exposure variables and the 32 eye diseases from FinnGen (R9) as outcome variables (disease code: H7). The *Eye2Brain* network explores causality from the 84 eye IDPs from UKBB as exposure variables to 41 FinnGen brain diseases as outcome variables (disease code: F5 and G6). Regarding significant signals related to brain diseases, we employed the GWAS summary statistics of 4 brain diseases from the PGC to replicate the signals independently (i.e., AD). Finally, The *Heart2Eye* network investigated causality from the 82 heart IDPs from UKBB as exposure variables to 41 brain diseases (disease code: F5 and G6) from FinnGen as outcome variables. The *Eye2Heart* network explored causality from the 84 eye IDPs from UKBB as exposure variables to 45 heart diseases (disease code: I9) from FinnGen as outcome variables.

The systematic quality-checking procedures to ensure unbiased exposure/outcome variable and instrumental variable (IVs) selection are detailed below.

We used a two-sample Mendelian randomization approach implemented in the *TwoSampleMR* package^80^ to infer the causal relationships within the two networks. We employed five distinct Mendelian randomization methods, presenting the results of the inverse variance weighted (IVW) method in the main text and the outcomes of the other four methods (Egger, weighted median, simple mode, and weighted mode estimators) in the supplement. The STROBE-MR Statement^135^ guided our analyses to increase transparency and reproducibility, encompassing the selection of exposure and outcome variables, reporting statistics, and implementing sensitivity checks to identify potential violations of underlying assumptions. First, we performed an unbiased quality check on the GWAS summary statistics. Notably, the absence of population overlapping bias^136^ was confirmed, given that FinnGen and UKBB participants largely represent populations of European ancestry without explicit overlap; PGC GWAS summary data were ensured to exclude UKBB participants. Furthermore, all consortia’s GWAS summary statistics were based on or lifted to GRCh37. Subsequently, we selected the effective exposure variables by assessing the statistical power of the exposure GWAS summary statistics in terms of instrumental variables (IVs), ensuring that the number of IVs exceeded 8 before harmonizing the data. Crucially, the function “*clump_data*” was applied to the exposure GWAS data, considering linkage disequilibrium. The function “*harmonise_data*” was then used to harmonize the GWAS summary statistics of the exposure and outcome variables. This yielded varying numbers of brain PSCs, heart IDPsm, and eye IDPs as effective exposure variables for the abovementioned six causal networks. Bonferroni correction was applied to all tested traits based on the number of effective PSC/IDPs or diseases, whichever was larger.

Finally, we conducted multiple sensitivity analyses. Initially, we conducted a heterogeneity test to scrutinize potential violations in the IV’s assumptions. To assess horizontal pleiotropy, which indicates the IV’s exclusivity assumption^137^, we utilized a funnel plot, single-SNP Mendelian randomization methods, and the Egger estimator. Furthermore, we performed a leave-one-out analysis, systematically excluding one instrument (SNP/IV) at a time, to gauge the sensitivity of the results to individual SNPs.

#### (j) PRS calculation

The PRS was computed using split-sample sensitivity GWASs for the heart and eye IDP-PRSs and discovery/replication GWASs for the brain PSCs. The PRS weights were established using split1/discovery GWAS data as the base/training set, while the split2/replication GWAS summary statistics served as the target/testing data. Both base and target data underwent rigorous quality control procedures involving several steps: *i*) excluding duplicated and ambiguous SNPs in the base data; *ii*) performing clumping of the base GWAS data; *iii*) pruning to remove highly correlated SNPs in the target data; *iv*) excluding high heterozygosity samples in the target data; and *v*) eliminating duplicated, mismatching, and ambiguous SNPs in the target data.

After completing the QC procedures, PRS for the split2 group was calculated using PLINK and the conventional C+T method (clumping + thresholding). To identify the most suitable PRS, we performed a linear regression considering various P-value thresholds (0.001, 0.05, 0.1, 0.2, 0.3, 0.4, 0.5), while accounting for age, sex, intracranial volume (if applicable), and the forty genetic principal components. The optimal P-value threshold for each brain PSC-PRS and heart IDP-PRS was determined based on achieving the highest incremental *R^2^*. We also explored the PRS-CS^139^ method as an alternative to derive the PRSs and compared its prediction power with the PLINK C+T approach. Overall, PRS-CS obtained a higher incremental *R^2^* than PLINK but was less generalizable to other domains (i.e., the 14 disease categories), as detailed in **Supplementary eText 8**.

### Method 6: Disease and cognition prediction

We evaluated the predictive performance of the 2003 brain PSCs, 82 heart IDPs, 84 eye IDPs, and their respective PRSs in predicting 14 systemic disease categories and 8 cognitive scores. Patient cohorts for the 14 disease categories were identified using the ICD-10 code accessible on the UKBB website (Data field: 41270 and 41202). The healthy control group consisted of participants without any ICD-10-based disease diagnoses. **Supplementary eTable 2** details the 14 ICD-10 disease categories and 8 cognitive scores.

#### (a) Pseudo R-squared (*R^2^*) statistics for individual feature-level prediction ability

We used logistic regression to determine the incremental *R^2^* of the 2003 brain PSCs, 82 heart, and 84 eye IDPs, and their respective PRSs for predicting 14 disease categories individually. The null model included age, sex, and disease status as the outcome variable, while the alternative model introduced the PSC/IDP or their corresponding PRS as an additional predictor. The incremental *R^2^* was calculated as the difference between the pseudo *R^2^* of the alternative model and that of the null model, employing the *PseudoR2* function from the *DescTools* R package (v 0.99.38). Detailed results, including other metrics like sample sizes, P-values, and β coefficients, are presented in the supplementary data.

#### (b) Systemic diseases classification

We evaluated the predictive capabilities of six distinct feature sets for each pair of organs, such as brain PSC, heart IDP, brain PSC + PSC-PRS, heart IDP + IDP-PRS, brain PSC + heart IDP, and brain PSC + PSC-PRS + heart IDP + IDP-PRS, using a support vector machine and a nested CV procedure. This approach involved CV training/validation/test datasets and separate independent test datasets to assess performance. We showed the nested CV test balanced accuracy (*CV accuracy*), which addresses sample imbalance. The nested CV procedure included an outer loop of 10-fold CV, allocating 80% for training/validation and 20% for testing. Within each iteration, an inner loop used 80% of the training/validation data for a 10-fold training/validation split to tune hyperparameters. Alongside the nested CV datasets, we reserved 250 patients and 250 healthy controls for independent test datasets (*Ind. accuracy*), whenever feasible, considering sample sizes. Combining features from different organs (e.g., brain and heart) often led to reduced sample sizes, precluding the use of independent test datasets in some tasks. Supplementary data include additional metrics (e.g., accuracy and sample sizes for the test data) for both test datasets. Additionally, the classification performance using neural networks (a five-layer fully connected network, nested CV was not applied) is detailed in supplementary data.

#### (c) Cognitive scores regression

Utilizing a similar nested CV approach, we employed the 6 feature sets from different organs to predict 8 cognitive scores (**Supplementary eTable 2**) using support vector regression and lasso regression. We used Pearson’s *r* as the main assessment metric derived from the training/validation/test datasets. Given the larger cognitive data sample sizes compared to classification tasks, we randomly sampled 1500 participants for the nested CV training/validation/test and left all other data as independent test datasets when applicable.

Supplementary data provide additional metrics, such as mean absolute error, P-values, and detailed sample sizes.

## Data Availability

The GWAS summary statistics corresponding to this study are publicly available on the MEDICINE knowledge portal (https://labs-laboratory.com/medicine/) and the BRIDGEPORT knowledge portal (https://labs-laboratory.com/bridgeport). Our study used data generated by the TCGA Research Network (https://www.cancer.gov/tcga), the human protein atlas (HPA: https://www.proteinatlas.org), and the STRING data (https://string-db.org/). The two platforms curated and consolidated publicly available (single-cell) RNA-seq and protein data, including the GTEx project (https://gtexportal.org/home/). Genomic loci annotation used data from FUMA (https://fuma.ctglab.nl/). PheWAS used data from the GWAS Atlas platform (https://atlas.ctglab.nl/PheWAS). GWAS summary data for the DEs were downloaded from the official websites of FinnGen (R9: https://www.finngen.fi/en/access_results) and PGC (https://pgc.unc.edu/for-researchers/download-results/). Individual data from UKBB can be requested with proper registration at https://www.ukbiobank.ac.uk/. The gene-drug-disease network used data from the DrugBank database (v.5.1.9; https://go.drugbank.com/). The analysis for partitioned heritability estimates used data from ROADMAP (https://egg2.wustl.edu/roadmap/web_portal/) and ENTEx (https://www.encodeproject.org/). All unrestricted data supporting the findings are also available from the corresponding author upon request.

## Code Availability

The software and resources used in this study are all publicly available:

- MLNI: https://github.com/anbai106/mlni, disease classification
- PLINK: https://www.cog-genomics.org/plink/, linear model GWAS, PRS
- FUMA: https://fuma.ctglab.nl/, gene mapping, genomic locus annotation
- GCTA: https://yanglab.westlake.edu.cn/software/gcta/#Overview, heritability estimates, and fastGWA
- LDSC: https://github.com/bulik/ldsc, genetic correlation, and heritability estimates
- TwoSampleMR: https://mrcieu.github.io/TwoSampleMR/index.html, MR
- Coloc: https://chr1swallace.github.io/coloc/, Bayesian colocalization
- PRS-CS: https://github.com/getian107/PRScs, PRS

## Authors’ contributions

Dr. Wen has full access to all the data in the study and takes responsibility for the integrity of the data and the accuracy of the data analysis.

*Study concept and design*: W.J

*Acquisition, analysis, or interpretation of data*: W.J

*Drafting of the manuscript*: W.J

*Critical revision of the manuscript for important intellectual content*: All authors

*Statistical analysis*: W.J with the help of B.P.A and D.M

## Acknowledgments

The MULTI consortium (J.W) aims to integrate multi-organ imaging with multi-omics data to advance our understanding of human aging and disease mechanisms. We want to express our sincere gratitude to the UK Biobank team for their invaluable contribution to advancing clinical research in our field (https://www.ukbiobank.ac.uk/). We also acknowledge the data sharing from the UKBB Eye and Vision Consortium (https://www.ukbiobank.ac.uk/enable-your-research/approved-research/genetic-contribution-to-vision-loss-and-disability-the-uk-biobank-eye-vision-consortium; Return ID: 1875) and UKB-PPP consortium (https://registry.opendata.aws/ukbppp/; Category code: 1838) to share the returned data with the community. We thank FinnGen (https://www.finngen.fi/en) and PGC (https://pgc.unc.edu/) for their generosity in sharing the GWAS summary statistics with the scientific community. We thank the BLSA participants and staff for their participation and continued dedication. The BLSA protocol was approved by the Institutional Review Board of the National Institute of Environmental Health Science, National Institutes of Health (03AG0325). This study used the UK Biobank resource under Application Number 35148 (D.C) under the NIH-funded iSTAGING consortium (D.C; grant number: RF1 AG054409). We thank Dr. Wenjia Bai for generously providing us access to the cardiac atlas utilized in his publication: https://wp.doc.ic.ac.uk/wbai/data/. We acknowledge the leadership of the Brain Imaging Genetics (BIG) workgroup, led by Dr. Tavia Evans, Dr. Natalia Vilor-Tejedor, and Dr. Junhao Wen, within the International Society to Advance Alzheimer’s Research and Treatment (ISTAART) community, for advocating brain imaging genetics in Alzheimer’s and aging research.

