## Supplementary materials for "Brain-heart-eye axis revealed by multi-organ imaging, genetics and proteomics"

### Online Supplementary Materials

**eMethod 1: The definition of genomic loci, independent significant SNP, lead SNP, candidate SNP**
**eMethod 2: Structural covariance patterns via stochastic orthogonally projective non-negative matrix factorization**
**eText 1: Sensitivity check analyses for the primary PWAS between the 2003 brain PSCs, 82 heart IDPs, and 84 eye IDPs**
**eText 2: Comparisons of our brain-heart PSC PWAS with 119 MUSE ROIs, Zhao et al., Jaggi et al., and Smith et al.**
**eText 3: Replication of the brain ProWAS results in the BLSA study.**
**eText 4: Sensitivity check analyses for the main GWAS of the 82 heart IDPs and 84 eye IDPs**
**eText 5: PheWAS of the genomic loci linked to the 2003 brain PSCs, 82 heart IDPs, and 84 eye IDPs**
**eText 6: Comparisons between our Brain2Herat and Heart2Brain causal networks and results from previous studies**
**eText 7: Individual prediction power of the 2003 brain PSCs, 83 heart IDPs, and 84 eye IDPs for the 14 disease categories and 8 cognitive scores**
**eText 8: Comparisons for PRS calculation between PLINK C+T approach and PRS-CS**
**eFigure 1: Secondary PWAS linking the brain PSCs, heart, and eye IDPs to 117 clinical phenotypes**
**eFigure 2: PheWAS of the loci linked to the 82 heart IDPs and 84 eye IDPs in the GWAS Atlas platform**
**eFigure 3: The correlation between the SNP-based heritability estimates of the 82 heart IDPs and 84 eye IDPs between Wen et al. and Zhao et al.**
**eFigure 4: Sensitivity check analysis for causal relationship from C512_352 to I9_HEARTFAILURE**
**eFigure 5: Sensitivity check analysis for causal relationship from C256_7 to I9_VARICVE**
**eFigure 6: Sensitivity check analysis for causal relationship from C1024_726 to I9_DISVEINLYMPH**
**eFigure 7: Sensitivity check analysis for causal relationship from C1024_880 to I9_HYPTENS**
**eFigure 8: Sensitivity check analysis for causal relationship from lv_circumferential_strai_global to F5_DEPRESSION_RECURRENT**
**eFigure 9: Sensitivity check analysis for causal relationship from descending_aorta_minimum_area to G6_MIGRANE_WITH_AURA**
**eFigure 10: Sensitivity check analysis for causal relationship from descending_aorta_maximum_area to G6_MIGRANE_WITH_AURA**
**eFigure 11: Sensitivity check analysis for causal relationship from global mean of the myocardial wall thickness of LV to G6_ALZHEIMER**
**eFigure 12: Sensitivity check analysis for causal relationship from C512_479 to H7 glaucoma**
**eFigure 13: Sensitivity check analysis for causal relationship from C512_479 to H7 POAG**
**eFigure 14: Sensitivity check analysis for causal relationship from INLEMLiL to F5 AD**

**eFigure 15: Sensitivity check analysis for causal relationship from RpeOsR to F5 the use of hypnotics and sedatives (RX_N05C)**

**eFigure 16: Sensitivity check analysis for causal relationship from MisL to F5 depression**

**eFigure 17: Sensitivity check analysis for causal relationship from MiiL to F5 sleep apnea**

**eFigure 18: Sensitivity check analysis for causal relationship from Dao_max to H7 STRABOTH**

**eFigure 19: Sensitivity check analysis for causal relationship from Dao_max to H7 OCUMUSCLE**

**eFigure 20: Sensitivity check analysis for causal relationship from Dao_min to H7 STRABOTH**

**eFigure 21: Sensitivity check analysis for causal relationship from Dao_min to H7 OCUMUSCLE**

**eFigure 22: Sensitivity check analysis for causal relationship from Aao_max to H7 GLAUCOMA**

**eFigure 23: Sensitivity check analysis for causal relationship from ElmIsosL to I9 CORATHER**

**eFigure 24: Incremental R2 for individual-level imaging features and their respective PRS for predicting the 14 systemic disease categories.**

**eFigure 25: Classification performance for predicting the 14 systemic disease categories using brain PSCs and conventional MUSE ROIs.**

**eFigure 26: The CV performance from the training/validation/test datasets for predicting the 8 cognitive scores**

**eFigure 27: The expression analyses of the MOG protein in the HPA**

**eFigure 28: The expression analyses of the PLTP protein in the HPA**

**eFigure 29: The expression analyses of the TGFA protein in the HPA**

**eFigure 30**: **Phenotypic correlations between the 119 MUSE ROIs and 128 brain PSCs at the scale of C=128**.

**eFigure 31**: **Genetic correlations between the 119 MUSE ROIs and 128 brain PSCs at the scale of C=128**.

**eFigure 32**: **Spatially overlap between C128_97 and the right orbital gyrus from the MUSE atlas.**

**eFigure 33**: **Genetic correlation between the 2003 brain PSCs and ~4000 brain IDPs from Smith et al**.

**eFigure 34: The correlation of the incremental $R^2$ of the heart IDP PRS between the methods and studies**

**ProWAS coefficients between the two sets of beta values of the brain ProWAS in the UKBB and the BLSA studies.**

**eTable 1: The 2003 brain PSCs and 82 heart IDPs**

**eTable 2: The 14 systemic disease categories and 8 cognitive scores**

**eFile 1-67: The Online Supplementary files contain large tables**

**eMethod 1: The definition of genomic loci, independent significant SNP, lead SNP, candidate SNP**

FUMA defined the significant independent SNPs, lead SNPs, candidate SNPs, and genomic risk loci as follows (https://fuma.ctglab.nl/tutorial#snp2gene):

*Independent significant SNPs*

They are defined as SNPs with $P \leq 5\times10^{-8}$ that are independent of each other at the user-defined $r^2$ (set to 0.6 in the current study). We further describe *candidate SNPs* as those in linkage disequilibrium (LD) with independent significant SNPs. FUMA then queries each candidate SNP in the GWAS Catalog to check whether any clinical traits have been reported to be associated with previous GWAS studies.

*Lead SNPs*

Lead SNPs are defined as independent significant SNPs that are also independent of each other at $r^2<0.1$. If multiple independent significant SNPs are correlated at $r^2 \geq 0.1$, then the one with the lowest individual *P*-value becomes the lead SNP. If $r^2$ threshold is set to 0.1 for the independent significant SNPs, then they would constitute the identical set as the lead SNPs. FUMA thus advises setting $r^2$ to be 0.6 or higher.

*Genomic risk loci*

FUMA defines genomic risk loci to include all independent signals physically close or overlapping in a single locus. First, independent significant SNPs dependent on each other at $r^2 \geq 0.1$ are assigned to the same genomic risk locus. Then, independent significant SNPs with less than the user-defined distance (250 kilobases by default) away from one another are merged into the same genomic risk locus - the distance between two LD blocks of two independent significant SNPs is the distance between the closest points from each LD block. Each locus is represented by the SNP within the locus with the lowest *P*-value.

**eMethod 2: Structural covariance patterns via stochastic orthogonally projective non-negative matrix factorization**

The sopNMF algorithm is a stochastic approximation built and extended based on opNMF[1,2]. We consider a dataset of $n$ MR images and $d$ voxels per image. We represent the data as a matrix $\boldsymbol{X}$ where each column corresponds to a flattened image: $\boldsymbol{X} = [x_1, x_2, \dots, x_n], \boldsymbol{X} \in \mathbb{R}_{\geq 0}^{d \times n}$. The sopNMF algorithm factorizes $\boldsymbol{X}$ into two low-rank ($r$) matrices $\boldsymbol{W} \in \mathbb{R}_{\geq 0}^{d \times r}$ and $\boldsymbol{H} \in \mathbb{R}_{\geq 0}^{r \times n}$ under the constraints of non-negativity and column-orthonormality. Using the Frobenius norm, the loss of this factorization problem can be formulated as

$$\|\boldsymbol{X} - \boldsymbol{WH}\|_F^2$$

$$\text{subject to } \boldsymbol{H} = \boldsymbol{W^T X}, \boldsymbol{W} \geq 0 \text{ and } \boldsymbol{W^T W} = \boldsymbol{I} \quad (1)$$

where $\boldsymbol{I}$ stands for the identity matrix. The columns $w_i \in \mathbb{R}^d$, $\|w_i\|^2 = 1, \forall\, i \in \{1..r\}$ of the so-called component matrix $\boldsymbol{W} = [w_1, w_2, \dots, w_r]$ are part-based representations promoting sparsity in data in this lower-dimensional subspace. From this perspective, the loading coefficient matrix $\boldsymbol{H}$ represents the importance (weights) of each feature above for a given image. Instead of optimizing the non-convex problem in a batch learning paradigm (i.e., reading all images into memory) as opNMF,[1] sopNMF subsamples the number of images at each iteration, thereby significantly reducing its memory demand by randomly drawing data batches $\boldsymbol{X_b} \in \mathbb{R}_{\geq 0}^{d \times b}$ of $b \leq n$ images ($b$ is the batch size; $b$=32 was used in the current analyses); this is done without replacement so that all data goes through the model once ($\lceil n/b \rceil$). In this case, the updating rule can be rewritten as

$$\boldsymbol{W}_{t+1} = \boldsymbol{W}_t \frac{\left(\boldsymbol{X_b X_b^T W}\right)_t}{\left(\boldsymbol{W W^T X_b X_b^T W}\right)_t} \quad (2)$$

We calculate the loss on the entire dataset at the end of each epoch (i.e., the loss is incremental across all batches) with the following expression:

$$\sum_{i=1}^{\lceil n/b \rceil} \left\|\boldsymbol{X_{b\_i}} - \boldsymbol{W W^T X_{bi}}\right\|_F^2 \quad (3)$$

We evaluated the training loss and the sparsity of $\boldsymbol{W}$ at the end of each iteration. Moreover, early stopping was implemented to improve training efficiency and alleviate overfitting. We summarize the sopNMF algorithm in **SI Algorithm 1**. An empirical comparison between sopNMF and opNMF is detailed in **SI eMethod 1**.

We applied sopNMF to the training population ($N$=4000). The component matrix $\boldsymbol{W}$ was sparse after the algorithm converged with a pre-defined maximum number of epochs (100 by default) with an early stopping criterion. To build the MuSIC atlas, we clustered each voxel (row-wise) into one of the $r$ features/PSCs as follows:

$$\boldsymbol{M}_j = \text{argmax}_k(\boldsymbol{W}_{j,k}) \ (4)$$

where $\boldsymbol{M}$ is a $d$-dimensional vector and $j \in \{1..d\}$. The $j$-th element of $\boldsymbol{M}$ equals $k$ if $\boldsymbol{W}_{j,k}$ is the maximum value of the $j$-th row. Intuitively, $M$ indicates which of the $r$ PSCs each voxel belongs to. We finally projected the vector $\boldsymbol{M} \in \mathbb{R}_{\geq 0}^{d}$ into the original image space to visualize each PSC of the MuSIC atlas (**Fig. 1**). Of note, 13 PSCs have vanished in this process for $C$=1024: all 0 for these 13 vectors.

We present the algorithm below:

**Algorithm 1:** Algorithm for sopNMF.

The source code of the Python implementation of sopNMF is available here: https://github.com/anbai106/SOPNMF

**Algorithm 1:** sopNMF

- **Input:** maximum number of epochs $e$, number of component $C$ or $r$, batch size $b$, early stopping criteria $\theta$ (i.e., the loss without decreasing for a certain epochs) ;
- **Output:** $\boldsymbol{W} \in \mathbb{R}^{d \times r}, \boldsymbol{H} \in \mathbb{R}^{r \times n}$ ;
- **Initialization:** $\boldsymbol{W}$ ;

**if** *not* $\theta$ *or epoch* $\neq e$ **then**
  **for** $p \leftarrow 0$ **to** $e$ **do**
    **for** $i \leftarrow 0$ **to** $t$ **do**
      *Read mini-batch* $\boldsymbol{X}_{bi}$
      *Update* $\boldsymbol{W}_{i+1}$ *via Eq. 2*
    **end**
    $loss = \sum_{i=1}^{\lceil \frac{n}{b} \rceil} \|\boldsymbol{X}_{bi} - \boldsymbol{W}\boldsymbol{W}^T\boldsymbol{X}_{bi}\|_F^2 (Eq.3)$
    **if** *loss in* $\theta$ **then**
      *Stop*
    **else**
      *Shuffle* $\boldsymbol{X}$
      *Continue*
    **end**
  **end**
**else**
  *Stop*
**end**

**eText 1: Sensitivity check analyses for the primary PWAS between the 2003 brain PSCs, 82 heart IDPs, and 84 eye IDPs**

***a) Brain-heart PWAS***:

We conducted two sensitivity checks to examine the reliability of our primary PWAS outcomes between the 2003 brain PSCs and 82 heart IDPs. For instance, in the split-sample PWAS, where both splits were randomly generated to maintain balanced age and sex distributions, split1 functioned as the discovery PWAS while split2 was the replication PWAS. We identified $N$ significant PSC-IDP associations in split1 PWAS that surpassed the Bonferroni-corrected threshold (P-value<0.05/2003/82). Subsequently, we assessed $N_n$ PSC-IDP associations that exceeded the nominal P-value threshold (0.05) and $N_b$ PSC-IDP associations that surpassed the Bonferroni-corrected P-value threshold (0.05/$N$). Regarding $\beta$ values, we evaluated the number of significant PSC-IDP associations ($N_{beta}$), where the $\beta$ values aligned with those derived from the $N$ PSC-IDP associations in split1 GWAS.

- The concordance rate of the P-value is defined as:
  - At the nominal threshold: CR-$P_n$=$N_n/N$
  - At the Bonferroni corrected threshold: CR-$P_b$=$N_b/N$
- The concordance rate of the $\beta$ values:
  - At the nominal threshold: CR-beta=$N_{beta}/N$
  - *r-$\beta$*=Pearson's *r* of the *$\beta$-discovery* and *$\beta$-replication SNPs from the $N_n$* PSC-IDP associations.

**Split-sample GWAS**

*P-values:*

In the split1 PWAS, we found 5974 PSC-IDP associations (P-value<0.05/2003/82). In the split2 PWAS, the mean concordance rate of the P-value was CR-$P_n$=0.99 using the nominal P-value and CR-$P_b$=0.72 for the Bonferroni-corrected threshold (<0.05/$N$).

*$\beta$ values:*

Compared to the $N$ PSC-IDP associations from the split1 PWAS, the split2 PWAS showed a mean concordance rate of CR-$\beta$=0.99. The $\beta$ values between the two splits were highly correlated (*r-$\beta$*=0.97, P-value<$1\times10^{-10}$). Detailed results of these statistics for each PSC-IDP association are presented in **Supplementary eFile 2**.

**Sex-stratified GWAS**

*P-values:*

In the female PWAS, we found 9574 PSC-IDP associations (P-value<0.05/2003/82). In the male PWAS, the mean concordance rate of the P-value was CR-$P_n$=0.91 using the nominal P-value and CR-$P_b$=0.38 for the Bonferroni-corrected threshold (<0.05/$N$).

*$\beta$ values:*

Compared to the $N$ PSC-IDP associations from the female PWAS, the male PWAS showed a mean concordance rate of CR-$\beta$=0.91. The $\beta$ values between the two splits were highly correlated (*r-$\beta$*=0.95, P-value<$1\times10^{-10}$). Detailed results of these statistics for each PSC-IDP association are presented in **Supplementary eFile 3**.

**Secondary PWAS:**

We also performed a secondary PWAS (**Method 3b**) to link the 2003 brain PSCs and 82 heart IDPs to 117 phenotypes linked to other organ systems available from our UKBB data download, excluding the multimodal brain IDPs used in Elliot et al.[3] (**Supplementary eFigure 1**) and the heart IDPs. In the heart IDP secondary PWAS, we detected 1110 significant IDP-phenotype associations (P-value<0.05/82/117). Notably, these associations highlighted connections with cardiovascular traits (15%, e.g., systolic blood pressure), immunological traits (15%, e.g., neutrophil count), metabolic traits (20%, e.g., cholesterol), and lifestyle and environmental factors (6%, e.g., alcohol consumption). **Supplementary eFile 8-9** present detailed statistics.

***b) Brain-eye PWAS***:
We conducted two sensitivity checks to examine the reliability of our primary PWAS outcomes between the 2003 brain PSCs and 84 eye IDPs. For instance, in the split-sample PWAS, where both splits were randomly generated to maintain balanced age and sex distributions, split1 functioned as the discovery PWAS, while split2 was the replication PWAS. We identified $N$ significant PSC-IDP associations in split1 PWAS that surpassed the Bonferroni-corrected threshold (P-value<0.05/2003). Subsequently, we assessed $N_n$ PSC-IDP associations that exceeded the nominal P-value threshold (0.05) and $N_b$ PSC-IDP associations that surpassed the Bonferroni-corrected P-value threshold (0.05/$N$). Regarding $\beta$ values, we evaluated the number of significant PSC-IDP associations ($N_{beta}$), where the $\beta$ values aligned with those derived from the $N$ PSC-IDP associations in split1 GWAS.

- The concordance rate of the P-value is defined as:
  - At the nominal threshold: $CR\text{-}P_n = N_n/N$
  - At the Bonferroni corrected threshold: $CR\text{-}P_b = N_b/N$
- The concordance rate of the $\beta$ values:
  - At the nominal threshold: $CR\text{-}beta = N_{beta}/N$
  - $r$-$\beta$=Pearson's $r$ of the *$\beta$-discovery* and *$\beta$-replication SNPs from the $N_n$* PSC-IDP associations.

**Split-sample GWAS**
*P-values:*
In the split1 PWAS, we found 380 PSC-IDP associations (P-value<0.05/2003). In the split2 PWAS, the mean concordance rate of the P-value was $CR\text{-}P_n$=0.67 using the nominal P-value and $CR\text{-}P_b$=0.2 for the Bonferroni-corrected threshold (<0.05/$N$).
*$\beta$ values:*
Compared to the $N$ PSC-IDP associations from the split1 PWAS, the split2 PWAS showed a mean concordance rate of CR-$\beta$=0.67. The $\beta$ values between the two splits were highly correlated ($r$-$\beta$=0.99, P-value<$1\times10^{-10}$). Detailed results of these statistics for each PSC-IDP association are presented in **Supplementary eFile 5**.

**Sex-stratified GWAS**
*P-values:*
In the female PWAS, we found 180 PSC-IDP associations (P-value<0.05/2003). In the male PWAS, the mean concordance rate of the P-value was $CR\text{-}P_n$=0.78 using the nominal P-value and $CR\text{-}P_b$=0.36 for the Bonferroni-corrected threshold (<0.05/$N$).
*$\beta$ values:*

Compared to the *N* PSC-IDP associations from the female PWAS, the male PWAS showed a mean concordance rate of CR-$\beta$=0.78. The $\beta$ values between the two splits were highly correlated ($r$-$\beta$=0.97, P-value<$1x10^{-10}$). Detailed results of these statistics for each PSC-IDP association are presented in **Supplementary eFile 6**.

**Secondary PWAS:**

We performed a secondary PWAS to link the 2003 brain PSCs and 84 eye IDPs to 117 phenotypes linked to other organ systems available from our UKBB data download, excluding the multimodal brain IDPs used in Elliot et al.[3]. In the brain PSC secondary PWAS, we uncovered 10,141 significant PSC-phenotype associations (P-value<0.05/2003/117). Within these associations, cognitive function-related phenotypes (56%, e.g., fluid intelligence score) and renal factors (17%, e.g., creatinine) demonstrated the most enriched correlations with the brain PSCs. In the eye IDP secondary PWAS, we detected 669 significant IDP-phenotype associations (P-value<0.05/84/117). Notably, these associations highlighted connections with musculoskeletal (20%, e.g., body weight), metabolic (18%, e.g., glucose), mental health (15%, e.g., health satisfaction), and cognitive function phenotypes (8%, e.g., fluid intelligence score). **Supplementary eFigure 1** and **eFile 10** present detailed statistics.

***c) Heart-eye PWAS***:

For the heart-eye PWAS, we did not perform any sensitivity analyses because they did not survive the Bonferroni corrections for multiple comparisons using the full sample sizes.

**eText 2: Comparisons of our brain-heart PSC PWAS with 119 MUSE ROIs, Zhao et al., Jaggi et al., and Smith et al.**

For the brain-heart PWAS, we have compared our results with those of two previous studies. Zhao et al.[4] performed PWAS using conventional atlas-based IDPs, and Jaggi et al.[5] performed using latent variables derived from PCA.

We used 119 MUSE GM ROIs to perform a PSC-MUSE PWAS and genetic correlation between them and 2003 PSCs. The analysis has two primary objectives. First, we aim to identify PSC-MUSE pairs exhibiting high correlation at phenotypic and genetic levels. Second, we intend to test Cheverud's conjecture, which proposes that the genetic correlation between two traits reflects their phenotypic correlation[6]. **Supplementary eFigure 30-31** show the PSC-MUSE pairs that passed the Bonferroni corrections (P-value < 0.05/2003/119) for both PWAS and genetic correlation analysis. We chose the scale of C=128 for the multi-scale brain PSCs because it's close to the resolution of the MUSE GM ROIs (*N*=119).

**Supporting evidence and exceptions for Cheverud's conjecture**:

As shown in **Supplementary eFigure 30-31**, most PSC-MUSE pairs confirm Cheverud's conjecture. However, in certain cases, the phenotypic correlation between the PSC-MUSE pairs showed opposite effects compared to their genetic correlation. For example, C128_97 (https://labs-laboratory.com/bridgeport/music/C128_97) exhibited a positive phenotypic correlation but a negative genetic correlation with the left cerebellum exterior. This discrepancy suggests multiple underlying factors, including the brain's potential environmental influences and compensatory mechanisms.

**Clinical interpretability of the brain PSCs**:

In addition, we found consistent PSC-MUSE correspondences using three independent approaches to increase the interpretability of the brain PSCs. Using C128_97 as an example, we discovered that this brain PSC: i) spatially overlapped with the known anatomical region (the right medial orbital gyrus), ii) showed a positive phenotypic correlation with this MUSE ROI, and iii) exhibited a positive genetic correlation with this MUSE ROI (**Supplementary eFigure 32** and BRIDGEPORT online functionality: https://labs-laboratory.com/bridgeport/music/right_medial_orbital_gyrus).

**Comparisons between C128_97 and right medial orbital gyrus for PWAS from MUSE and Zhao et al.**:

Based on the strong link between C128_97 and the right orbital gyrus, we sought to determine the additional insights the data-driven PSC could offer to our PWAS compared to the conventional MUSE ROI (right orbital gyrus) and from Zhao el al using Freesurfer IDP (right orbital gyrus).

In our primary brain-heart PWAS, we found that C128_97 was linked to 8 heart IDPs that passed the Bonferroni correction (0.05/2003/82), including the lv_myocardial_mass_f24105_2_0, rv_end_diastolic_volume_f24106_2_0, ra_stroke_volume_f24116_2_0, ascending_aorta_maximum_area_f24118_2_0, ascending_aorta_minimum_area_f24119_2_0, lv_mean_myocardial_wall_thickness_aha_4_f24127_2_0, lv_mean_myocardial_wall_thickness_aha_6_f24129_2_0, and lv_mean_myocardial_wall_thickness_global_f24140_2_0. In Zhao et al., their primary PWAS

(**Supplementary Table S3**) found that their Freesurfer atlas (Desikan) IDP (right.pars.orbitalis) was significantly linked to two heart IDPs, albeit with a more stringent P-value threshold (<1.33x10$^{-6}$), including the AAO_max_area (ascending_aorta_maximum_area_f24118_2_0) and AAO_min_area (ascending_aorta_minimum_area_f24119_2_0). To more fairly compare PWAS of C128_97 with conventional atlas-based IDP (ensuring similar sample sizes, covariates P-value threshold), we found that the MUSE ROI: the right medial orbital gyrus (P-value < 0.05/2003/82) was significantly associated with 6 heart IDPs, including the lv_myocardial_mass_f24105_2_0, rv_end_diastolic_volume_f24106_2_0, ra_stroke_volume_f24116_2_0, ascending_aorta_maximum_area_f24118_2_0, ascending_aorta_minimum_area_f24119_2_0, lv_mean_myocardial_wall_thickness_aha_4_f24127_2_0.

We compared this PSC and MUSE PWAS to compare this PSC vs. the MUSE ROI statistically. This involved 100 iterations of both features, each using a randomly down-sampled cohort of 20,000 individuals. A two-sample t-test revealed that PSC identified a higher number of significant heart-brain associations at the nominal level (P = 0.047).

While direct comparison with Jaggi et al.'s brain-heart PWAS was likely impossible due to the use of Principal Component Analysis (PCA), which lacks part-based representation, this data-driven approach may still offer enhanced feature extraction compared to conventional brain IDPs.

**Genetic correlations between the 2003 brain PSCs and 3874 brain IDPs from Smith et al.**

We estimated pair-wise genetic correlation between our 2003 brain PSCs vs. 3936 (38 IDPs were downloaded with errors) brain IDPs from Smith et al. shared in the BIG-40 platform (https://open.win.ox.ac.uk/ukbiobank/big40/).

LDSC was able to converge for 7,371,077 out of 7,883,808 estimations. Common failures of the estimations were due to the limited $h^2$ estimates for one of the two traits. A common failure of model convergence was due to the Jackknife estimation (https://github.com/bulik/ldsc/issues/54). We then corrected the P-value for multiple comparisons at a stringent level (P-valye<0.05/3936=1.27x10$^{-5}$), resulting in a total of 109,209 significant associations ($g_c$ [-0.69, 1.134]) between 1927 unique brain PSCs and 1966 brain IDPs. Of note, LDSC is not bounded, although theoretically, the $g_c$ should be within [-1, 1] (see https://github.com/bulik/ldsc/issues/89).

The most significant association (P-value<1.64x10$^{50}$; $g_c$=0.86±0.02) was achieved between C32_1 and the volume of subcortical grey matter derived from Freesurfer ASEG pipeline (Field ID: 26517; https://biobank.ndph.ox.ac.uk/showcase/field.cgi?id=26517), which perfectly align with our data-driven brain PSC covering the covariance network of subcortical structures (visualization: https://labs-laboratory.com/bridgeport/music/C32_1). Other conventional brain IDPs linked to C32_1 (in total 243) included volume of putamen (right hemisphere) (Field ID: 26591: https://biobank.ndph.ox.ac.uk/showcase/field.cgi?id=26591) and volume of putamen (left hemisphere) (Field ID: 26560: https://biobank.ndph.ox.ac.uk/showcase/field.cgi?id=26560).

For the 109,209 associations, we found our 1966 GM-based brain PSCs were associated with 67,979 IDPs of regional and tissue volume (e.g., Field ID: 25003: volume of ventricular cerebrospinal fluid) and 19,787 IDPs of cortical area (e.g., Field ID: 26832: area of lateral-occipital (right hemisphere), Freesurfer Desikan atlas) derived from T1-weighted MRI, and 6897 IDPs of WM tract diffusivity (e.g., Field ID: 25227: mean L1 in posterior corona radiata on FA skeleton (left)). Of note, the 3936 conventional brain IDPs cover all the brain imaging modalities available at UKBB, whereas our brain PSCs were derived only using T1-weighted MRI (i.e., GM voxel image). As expected, we obtained the highest number of associations with IDPs derived from GM measures from T1-weighted MRI. **Supplementary eFile 64** presents the detailed statistics for this large-scale genetic correlation analysis using LDSC. In **eFigure 33**, we presented the significant genetic associations of our brain PSCs and the 3936 conventional brain IDPs from the BIG-40 portal.

**Evaluate the robustness of the secondary heart PWAS by including house income as an additional covariate**:

We further analyzed the influence of house income on our secondary PWAS by using one of the strongest signals: ra_maximum_volume_f24114 vs. triglycerides_f30870. These comparative results are listed below:

- The original PWAS model reported a -log(P-value=83.55) and $\beta$ coefficient=-2.993± 0.1533 ($N$=29713).
- The new model, including house income as an additional covariate, reported a -log(P-value=83.67) and $\beta$ coefficient=-2.996± 0.1534 ($N$=29694).

**eText 3: Replication of the brain ProWAS results in the BLSA study.**

To validate the significant brain ProWAS associations, we leveraged plasma proteomics data from the Baltimore Longitudinal Study of Aging (BLSA[7,8]) study. Initially, the UK Biobank (UKBB) brain ProWAS analysis revealed 1282 significant correlations between 558 brain PSCs and 27 proteins, as assessed by the Olink platform. We subsequently sought to replicate these findings using 7,268 proteins from 924 participants in the BLSA study, who were assayed with the SomaScan v4.1 platform. We applied stringent Bonferroni-corrected P-value thresholds of 0.05/558/27, as well as a nominal P-value threshold, to verify the robustness of these associations

Applying the Bonferroni-corrected P-value threshold, we identified 45 brain PSC-protein pairs in the BLSA that exhibited concordant and correlated beta values with those observed in the UKBB cohort, as illustrated in **Supplementary eFigure 35a**. For example, we identified a positive correlation between C32_2 (https://labs-laboratory.com/bridgeport/music/C32_2) and the NCAN protein in both UKBB [-log(P-value)=23.42; $\beta$=115.71] and BLSA [-log(P-value)=9.15; $\beta$=140.98] analyses (**Supplementary eFigure 35b**).

At the nominal P-value threshold, we detected some proteins with divergent $\beta$ estimate signs between the UKBB Olink and BLSA SomaScan datasets, indicating potential discordance (**Supplementary eFigure 35c**). For example, the association between C512_295 (https://labs-laboratory.com/bridgeport/music/C512_295) and the MOG protein was positive in UKBB [-log(P-value)=8.17; $\beta$=5.10] but negative in BLSA [-log(P-value)=1.78; $\beta$=-5.14] analyses (**Supplementary eFigure 35d**). This inconsistency could be due to several factors. First, Olink and SomaScan are two distinct proteomics platforms that utilize fundamentally different technologies to measure protein levels. These differences can lead to variations in sensitivity and specificity, resulting in discordant $\beta$ signs. Secondly, although we included sufficient covariates in both analyses, the UKBB and BLSA cohorts may have unique demographic, clinical, or environmental characteristics influencing protein levels and/or their associations. Thirdly, Olink and SomaScan might detect different proteoforms arising from alternative splicing, post-translational modifications, etc., which might account for opposite patterns of associations, at least at the nominally corrected P-value threshold.

**eText 4: Sensitivity check analyses for the main GWAS of the 82 heart IDPs and 84 eye IDPs**

***a) Heart IDP GWAS***:
We conducted four sensitivity checks to examine the reliability of our primary GWAS outcomes thoroughly – these assessments aimed to gauge the consistency of genetic signals across various conditions. For instance, in the split-sample GWAS, where both splits were randomly generated to maintain balanced age and sex distributions, split1 functioned as the discovery GWAS while split2 was the replication GWAS. We identified $N$ SNPs in split1 GWAS that surpassed the genome-wide significance threshold (P-value<5x10$^{-8}$). Subsequently, we assessed $N_n$ SNPs that exceeded the nominal P-value threshold (0.05) and $N_b$ SNPs that surpassed the Bonferroni-corrected P-value threshold (0.05/$N$). Regarding $\beta$ values, we evaluated the number of SNPs ($N_{beta}$), where the $\beta$ values aligned with those derived from the $N$ SNPs in split1 GWAS.

- The concordance rate of the P-value is defined as:
    - At the nominal threshold: CR-$P_n$=$N_n/N$
    - At the Boneforroni corrected threshold: CR-$P_b$=$N_b/N$
- The concordance rate of the $\beta$ values:
    - At the nominal threshold: CR-$\beta$=$N_{beta}/N_n$
    - $r$-$\beta$=Pearson's $r$ of the *$\beta$-discovery* and *$\beta$-replication SNPs from the $N_n$ SNPs.*

**Split-sample GWAS**
*P-values:*
In the split1 GWAS, we found 75 (1, 791) IDP-SNP associations for the 41 heart IDPs (P-value < 5x10$^{-8}$). In the split2 GWAS, the mean concordance rate of the P-value was CR-$P_n$=0.97 using the nominal P-value and CR-$P_b$=0.82 for the Bonferroni-corrected threshold (<0.05/$N$).
*$\beta$ values:*
Compared to the $N_n$ SNPs from the split1 GWAS, the split2 GWAS showed a mean concordance rate of CR-$\beta$=1 and $r$-$\beta$=0.94. Detailed results of these statistics for each IDP are presented in **Supplementary eFile 17**.

**Sex-stratified GWAS**
*P-values:*
In the female GWAS, we found 77 (1, 664) IDP-SNP associations for the 37 heart IDPs (P-value < 5x10$^{-8}$). In the male GWAS, the mean concordance rate of the P-value was CR-$P_n$=0.94 using the nominal P-value and CR-$P_b$=0.76 for the Bonferroni-corrected threshold (<0.05/$N$).
*$\beta$ values:*
Compared to the $N_n$ SNPs from the female GWAS, the male GWAS showed a mean concordance rate of CR-$\beta$=1 and $r$-$\beta$=0.78. Detailed results of these statistics for each IDP are presented in **Supplementary eFile 18**.

**Non-European GWAS**
*P-values:*
In the fastGWA European GWAS, we found 316 (2, 2680) IDP-SNP associations for the 68 heart IDPs (P-value < 5x10$^{-8}$). In the fastGWA non-European GWAS, the mean concordance rate of the P-value was CR-$P_n$=0.50 using the nominal P-value and CR-$P_b$=0.02 for the Bonferroni-corrected threshold (<0.05/$N$).

*$\beta$ values:*

Compared to the $N_n$ SNPs from the European GWAS, the non-European GWAS showed a mean concordance rate of CR-$\beta$=1. The $\beta$ values of the two sets were highly correlated ($r$-$\beta$=0.84). Detailed results of these statistics for each IDP are presented in **Supplementary eFile 19**.

**Wen et al. vs. Zhao et al. heart IDP GWAS**

All 82 heart IDPs intersected between our GWAS and Zhao et al.'s study. We evaluated the consistency between these two GWAS sets utilizing UKBB data, acknowledging potential variations in sample sizes for each specific IDP (Zhao et al. did not specify the sample size of each IDP GWAS). Furthermore, a portion of the SNPs tested revealed swapped A1 and A2 alleles between the two studies, necessitating SNP harmonization by flipping them along with the $\beta$ values.

*P-values:*

In Zhao's GWAS, they found 449 (1, 3146) IDP-SNP associations for the 68 heart IDPs (P-value < $5x10^{-8}$). In our GWAS, the mean concordance rate of the P-value was CR-$P_n$=0.65 ±0.25 using the nominal P-value and CR-$P_b$=0.60 ±0.26 for the Bonferroni-corrected threshold (<0.05/$N$).

*$\beta$ values:*

Compared to the $N_n$ SNPs from Zhao's GWAS, our GWAS showed a mean concordance rate of CR-$\beta$=0.999. Furthermore, the $\beta$ values of the two sets were highly correlated ($r$-$\beta$=0.90). Detailed results of these statistics for each IDP are presented in **Supplementary eFile 20**.

Our GWAS largely aligns with the signals identified in the previous heart IDP GWAS by Zhao et al., indicating robustness. However, differences in sample sizes, GWAS models, and preprocessing methods (e.g., the number of QC'ed valid SNPs) might contribute to some discrepancies.

***b) Eye IDP GWAS***:

We conducted five sensitivity checks to examine the reliability of our primary GWAS outcomes thoroughly – these assessments aimed to gauge the consistency of genetic signals across various conditions. For instance, in the split-sample GWAS, where both splits were randomly generated to maintain balanced age and sex distributions, split1 functioned as the discovery GWAS, while split2 was the replication GWAS. We identified $N$ SNPs in split1 GWAS that surpassed the genome-wide significance threshold (P-value<$5x10^{-8}$). Subsequently, we assessed $N_n$ SNPs that exceeded the nominal P-value threshold (0.05) and $N_b$ SNPs that surpassed the Bonferroni-corrected P-value threshold (0.05/$N$). Regarding $\beta$ values, we evaluated the number of SNPs ($N_{beta}$), where the $\beta$ values aligned with those derived from the $N$ SNPs in split1 GWAS.

- The concordance rate of the P-value is defined as:
  - At the nominal threshold: CR-$P_n$=$N_n/N$
  - At the Bonferroni corrected threshold: CR-$P_b$=$N_b/N$
- The concordance rate of the $\beta$ values:
  - At the nominal threshold: CR-$\beta$=$N_{beta}/N_n$
  - $r$-$\beta$=Pearson's $r$ of the *$\beta$-discovery* and *$\beta$-replication SNPs from the $N_n$ SNPs.*

**Split-sample GWAS**

*P-values:*

In the split1 GWAS, we found 930 (7, 3057) IDP-SNP associations for the 84 eye IDPs (P-value < $5x10^{-8}$). In the split2 GWAS, the mean concordance rate of the P-value was CR-$P_n$=0.98±0.03

using the nominal P-value and CR-$P_b$=0.86±0.11 for the Bonferroni-corrected threshold (<0.05/*N*).

*β values:*

Compared to the $N_n$ SNPs from the split1 GWAS, the split2 GWAS showed a mean concordance rate of CR-*β*=1 and *r-β*=0.97±0.04. Detailed results of these statistics for each IDP are presented in **Supplementary eFile 21**.

**Sex-stratified GWAS**

*P-values:*

In the female GWAS, we found 951 (25, 3291) IDP-SNP associations for the 84 eye IDPs (P-value < $5x10^{-8}$). In the male GWAS, the mean concordance rate of the P-value was CR-$P_n$=0.95±0.09 using the nominal P-value and CR-$P_b$=0.80±0.15 for the Bonferroni-corrected threshold (<0.05/*N*).

*β values:*

Compared to the $N_n$ SNPs from the female GWAS, the male GWAS showed a mean concordance rate of CR-*β*=1 and *r-β*=0.96±0.11. Detailed results of these statistics for each IDP are presented in **Supplementary eFile 22**.

**fastGWA vs. PLINK GWAS**

*P-values:*

In the fastGWA mixed-effect linear model GWAS, we found 2452 (157, 6935) IDP-SNP associations for the 84 eye IDPs (P-value < $5x10^{-8}$). In the PLINK linear model GWAS (smaller sample sizes due to explicitly excluding related individuals in the populations), the mean concordance rate of the P-value was CR-$P_n$=1±0.00 using both the nominal P-value and the Bonferroni-corrected thresholds.

*β values:*

Compared to the $N_n$ SNPs from the fastGWA GWAS, the PLINK GWAS showed a mean concordance rate of CR-*β*=1 and *r-β*=0.99±0.0008. Detailed results of these statistics for each IDP are presented in **Supplementary eFile 23**.

While PLINK's linear model GWAS aligns well with the outcomes generated by fastGWA, particularly with a smaller sample size under the current non-strict replication procedure (not meeting the genome-wide P-value threshold), it's advisable to implement linear mixed-effect models in practical applications, possibly modeling additional cryptic population stratification.

**Non-European GWAS**

*P-values:*

In the fastGWA European GWAS, we found 2452 (157, 6935) IDP-SNP associations for the 84 eye IDPs (P-value < $5x10^{-8}$). In the fastGWA non-European GWAS, the mean concordance rate of the P-value was CR-$P_n$=0.63±0.14 using the nominal P-value and CR-$P_b$=0.11±0.12 for the Bonferroni-corrected threshold (<0.05/*N*).

*β values:*

Compared to the $N_n$ SNPs from the European GWAS, the non-European GWAS showed a mean concordance rate of CR-*β*=0.99±0.06. However, the *β* values of the two sets were highly correlated (*r-β*=0.95±0.11; P-value<$1x10^{-10}$). Detailed results of these statistics for each IDP are presented in **Supplementary eFile 24**.

**Wen et al. vs. Zhao et al. eye IDP GWAS**

46 eye IDPs intersected between our GWAS and Zhao et al.'s study. We evaluated the consistency between these two GWAS sets utilizing UKBB data, acknowledging potential variations in sample sizes for each specific IDP (Zhao et al. did not specify the sample size of each IDP GWAS). Furthermore, a portion of the SNPs tested revealed swapped A1 and A2 alleles between the two studies, necessitating SNP harmonization by flipping them along with the $\beta$ values.

*P-values:*

In Zhao's GWAS, they found 4323 (895, 9162) IDP-SNP associations for the 46 eye IDPs (P-value < $5 \times 10^{-8}$). In our GWAS, the mean concordance rate of the P-value was CR-$P_n$=0.72 ±0.10 using the nominal P-value and CR-$P_b$=0.71±0.10 for the Bonferroni-corrected threshold (<0.05/*N*).

*$\beta$ values:*

Compared to the $N_n$ SNPs from Zhao's GWAS, our GWAS showed a mean concordance rate of CR-$\beta$=1. In addition, the $\beta$ values of the two sets were highly correlated ($r$-$\beta$=0.996±0.001; P-value<$1 \times 10^{-10}$). Detailed results of these statistics for each IDP are presented in **Supplementary eFile 25**.

Our GWAS largely aligns with the signals identified in the previous eye IDP GWAS by Zhao et al., indicating robustness. However, differences in sample sizes, GWAS models, and preprocessing methods (e.g., the number of QC'ed valid SNPs) might contribute to some discrepancies.

**eText 5: PheWAS of the genomic loci linked to the 82 heart IDPs and 84 eye IDPs**

***a) Heart IDP***

To uncover the phenome-wide associations of these heart IDP-linked genomic loci, we performed a PheWAS look-up analysis using the GWAS Atlas[9] platform for these top lead SNPs. This identified 2939 previous SNP-trait associations in prior GWASs (**Supplementary Figure 2**). For the loci linked to the 82 heart IDPs, around 66% were related to IDPs derived from LV. The most enriched clinical traits were immunological traits (23%, e.g., basophil). Cardiovascular traits, like blood pressure and various heart diseases, were prevalent, accounting for 12% of the associations. Several other brain-related traits, including brain IDPs (e.g., white matter diffusivity and hippocampus volume), cognitive traits (e.g., intelligence), and psychiatric characteristics (e.g., neuroticism), also displayed considerable associations.

***b) Eye IDP***

To delineate the phenome-wide associations of these eye IDP-linked genomic loci, we performed a PheWAS look-up analysis using the GWAS Atlas[9] platform, specifically targeting these top lead SNPs while accounting for linkage disequilibrium. Of these 1888 locus-IDP pairs, 520 were not previously linked to clinical traits as of the November 25, 2023 query date. This PheWAS identified 8493 previously established SNP-trait associations in prior literature. Around 27% of associations were related to overall macular thickness. For the previously established clinical traits, the most enriched clinical traits were immunological traits (21%). Convincingly, ophthalmological conditions, like glaucoma and age-related macular degeneration, were also notably prevalent, accounting for 6% of the associations. Brain IDPs (e.g., white matter diffusivity and hippocampus volume), cognitive traits (e.g., intelligence), and psychiatric characteristics (e.g., neuroticism) also displayed considerable associations (**Supplementary eFigure 2**).

#### eText 6: Comparisons between our *Brain2Heart* and *Heart2Brain* causal networks and results from previous studies

**Comparison to Zhao et al.**

Compared to Zhao et al.[4], our Heart2Brain causal network expands this study from the following aspects. First, Zhao et al.'s causal analysis prioritized curtained traits by including 11 well-powered ($n$ > 20,000) brain-related clinical outcomes from FinnGen and six neuropsychiatric disorders from PGC. In contrast, we performed a systematic yet hypothesis-free quality check to include all available data from FinnGen and PGC, resulting in 82 heart IDPs as exposure variables and 41 brain diseases as outcome variables for the *Heart2Brain* causal network. Second, our MR analysis was inspired by the endophenotype hypothesis[10], where the imaging-derived endophenotypes serve as intermediate phenotypes that link genetics and exo-phenotypes (disease endpoints in this case): genetics → proteomics → PSC/IDP → disease endpoint. Therefore, we only tested the causal direction from the brain PSCs to heart diseases. Instead, when constructing the *Brain2Heart*, we used the brain PSCs as exposure and heart-related diseases as outcome variables. This was different from Zhao et al.'s approaches.

Our findings broadly align with those of Zhao et al., though they identified more causal signals from heart to brain than vice versa. This pattern partially supports the endophenotype hypothesis, suggesting that intermediate phenotypes like PSCs/IDPs are more likely to causally influence disease endpoints rather than the reverse. For example, we revealed a concordant causal relationship between heart IDPs and depression, consistent with Zhao et al., despite differences in data sources (FinnGen vs. SSGAC GWAS summary data for Zhao et al). Notably, our study identifies a new causal link between heart IDPs and Alzheimer's disease (AD), a connection supported by the well-established relationship between cardiovascular factors and AD. While our results complement those of Zhao et al., adding new insights into heart IDP relationships with AD and migraine, their analysis also uncovered a causal association with bipolar disorder (using PGC data) not detected in our study, likely due to differences in GWAS summary analysis power and multiple comparison thresholding.

**Comparison to Lin et al.**

Compared to Lin et al.[11], in which the authors used the six cardiovascular disease endpoints as outcome variables for the *Brain2Heart* causal results, our results were highly consistent. In Lin et al., the results related to the 6 cardiovascular disease endpoints did not survive the Bonferroni correction (P-value $< 0.05/2273 = \sim 2.20 \times 10^{-5}$). They uncovered causal relationships between specific brain regions and cardiovascular outcomes, including a causal link between i) left putamen volume (IDP 0126) and heart failure, and ii) right caudate volume (IDP 0125) and hypertension.

Our analysis revealed potential causal relationships between brain regions C512_352 and heart failure and C1024_880 and hypertension. Notably, C512_352 (https://labs-laboratory.com/bridgeport/music/C512_352), a subcortical brain structure, shows a phenotypic correlation ($r$=0.21, P<0.05) with left putamen but no genetic correlation ($g_c$=0.05, P>0.05), suggesting that the identified causal signals may represent new biological pathways that are distinct from previously reported associations (e.g., the one reported in Lin et al.); this warrants further investigation in future studies.

**eText 7: Individual prediction power of the 2003 brain PSCs, 83 heart IDPs, and 84 eye IDPs for the 14 disease categories and 8 cognitive scores.**

At the individual PSC/IDP/PRS level, we discovered that both the brain PSCs and heart IDPs significantly enhanced the predictive capability, evidenced by the incremental $R^2$ across various disease categories (**Supplementary eFigure 24**). This underscores the added predictive strength beyond common demographic factors such as age and sex. We presented the top 10 notable features showing the highest incremental $R^2$ within each feature set. Specifically, the 10 representative brain PSCs and 10 brain PSC-PRSs demonstrated the strongest predictive capacity for mental and behavioral disorders (ICD10 code: F). Likewise, the 10 most predictive heart IDPs and IDP-PRSs showed notable predictive abilities for circulatory system diseases (ICD10 code: J) and mental and behavioral disorders. The 10-representative eye IDPs and 10 eye IDP-PRSs were notably predictive for eye diseases (ICD10 code: H0-5) and mental and behavioral disorders. This underscores the clinical significance of these PSCs/IDPs/PRSs. More detailed results are shown in **Supplementary eFile 53-55**.

**eText 8: Comparisons for PRS calculation between PLINK C+T approach and PRS-CS.** We also explored the PRS-CS[12] method as an alternative to derive the PRSs. This method estimates posterior SNP effect sizes under continuous shrinkage priors using GWAS summary statistics and an LD reference panel (i.e., UKBB reference). PRS-CS does not require a clumping procedure, as the method takes LD into account through modeling. To fairly compare the predictive power with PLINK, we explicitly adopted the same P-value threshold method to ensure the inclusion of the same set of SNPs. Compared to the PLINK method, PRS-CS achieved higher incremental $R^2$ in explaining the phenotypes of the brain PSCs and heart PSCs phenotypes. Specifically, for the 82 heart IDP-PRSs, the PRSs from PLINK [$R^2$=0.0043 (0.0001, 0.029)] and PRC-CS [$R^2$=0.0080 (0.0002, 0.074)] were highly concordant and correlated (Pearson's *r* correlation=0.97, P-value=$1.69 \times 10^{-51}$, **Supplementary eFigure 34a**). Nevertheless, the predictive capacity of the PRSs obtained from PRS-CS is comparatively lower for the 14 systemic disease categories (cross-domain from PSCs/IDPs to systemic disease categories), particularly concerning the brain and heart-related categories. This observation aligns with our earlier study[13]. Even though there was a higher incremental $R^2$ in predicting the original phenotypes (i.e., the heart IDPs), the predictive power was reduced in translating to other domains. Consequently, we utilized the PRSs derived from PLINK for the prediction analyses presented in **Fig. 8**. Finally, we conducted a comparison of the 82 heart IDP-PRSs and 84 eye IDP-PRSs with those of Zhao et al.[14], revealing a strong correlation between the two sets (Pearson's *r* correlation=0.95, P-value<$1 \times 10^{-10}$, **Supplementary eFigure 34b**).

The same trend persisted for the brain and eye PRS predictions. Compared to the PLINK method, PRS-CS obtained higher incremental $R^2$ to explain the phenotypes of the two organs, but the two sets of estimation were highly correlated. For the 84 eye IDP-PRSs, the PRSs from PLINK [$R^2$=0.018 (0.001, 0.061)] and PRS-CS [$R^2$=0.041 (0.002, 0.115)] were highly concordant and correlated (Pearson's *r* correlation=0.99, P-value<$1 \times 10^{-10}$; **Supplementary eFile 55** vs. **67**) for PLINK vs. PRS-CS. However, the PRSs derived from PRS-CS are less predictive for the 14 systemic disease categories (cross-domain from PSCs/IDPs to systemic disease categories), especially for the brain and eye-related categories. For example, among the 84 eye IDP-PRSs, the most predictive eye IDP-PRS explained an incremental $R^2$=0.0612 (low power) in predicting ear diseases (i.e., disease effect non-sensitive) using PRS-CS. In contrast, this increased to an $R^2$=0.23 in predicting eye diseases with GcplL using PLINK. This general low, biologically non-sensitive generalizability of the PRS-CS, albeit with a higher incremental $R^2$ to predict the original phenotypes (i.e., the eye IDPs), enabled us to use the PRSs derived from PLINK for prediction analyses presented in **Fig. 8**. To assess our PRS-CS findings rigorously, we conducted a comparison of 46 overlapping IDP-PRSs with those of Zhao et al. using the same method, revealing a strong correlation between the two sets (Pearson's *r* correlation=0.92, P-value<$1 \times 10^{-10}$).

**eFigure 1: Secondary PWAS linking the brain PSCs, heart, and eye IDPs to 117 clinical phenotypes**

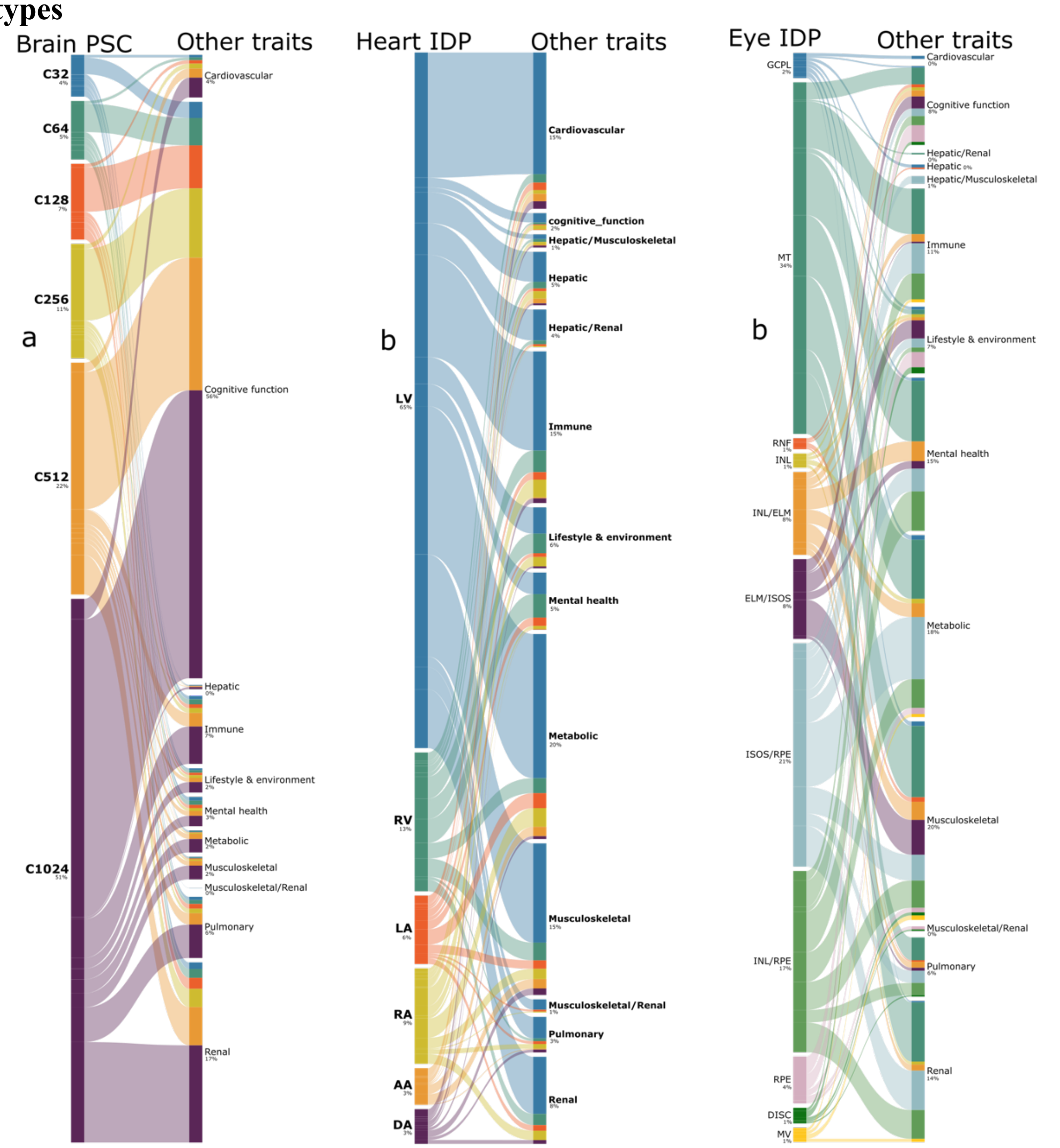


**a**) Brain PSC secondary PWAS associates the 2003 brain PSCs (shown at the high-level scales for C32-1024) to the other 117 clinical phenotypes available in UKBB. **b**) Heart IDP secondary PWAS associates the 82 heart IDPs (shown at the high-level categories) to the other 117 clinical phenotypes available in UKBB. **c**) eye IDP secondary PWAS associates the 84 eye IDPs (shown at the high-level categories) to the other 117 clinical phenotypes available in UKBB.

**eFigure 2: PheWAS of the loci linked to the 82 heart IDPs and 84 eye IDPs in the GWAS Atlas platform**

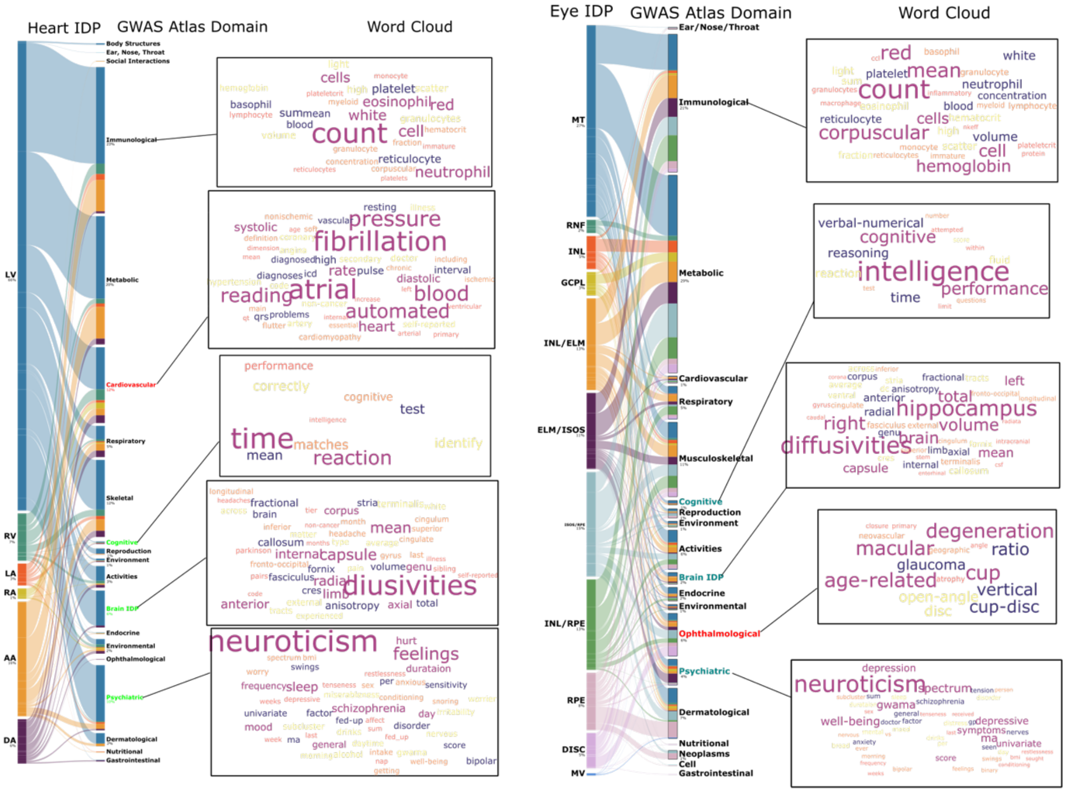


PheWAS links the top lead SNPs within the genomic loci of the 82 heart IDPs (left panel) and 84 eye IDPs (right panel) to all previously established clinical traits in the GWAS Atlas platform. We used the word cloud plots to showcase the representative traits for each GWAS Atlas trait domain.

**eFigure 3: The correlation between the SNP-based heritability estimates of the 82 heart IDPs and 84 eye IDPs between Wen et al. and Zhao et al.**

**a) heart IDPs**

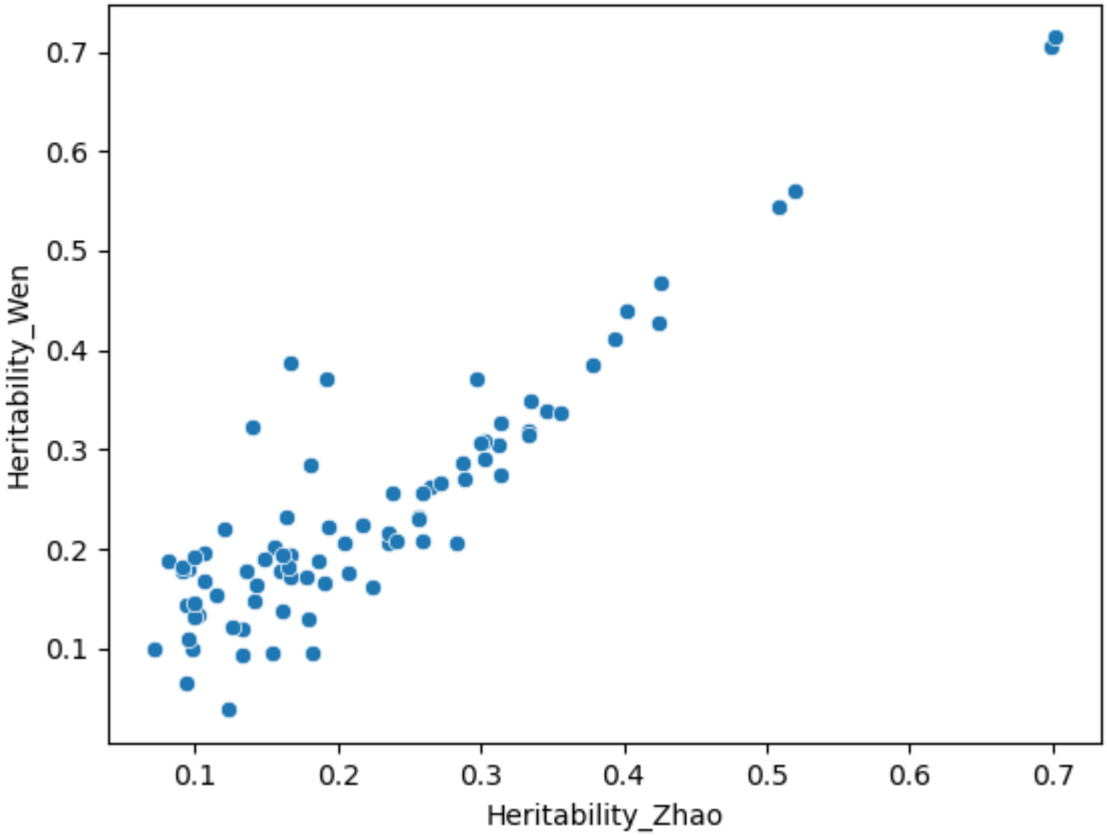

**a) eye IDPs**

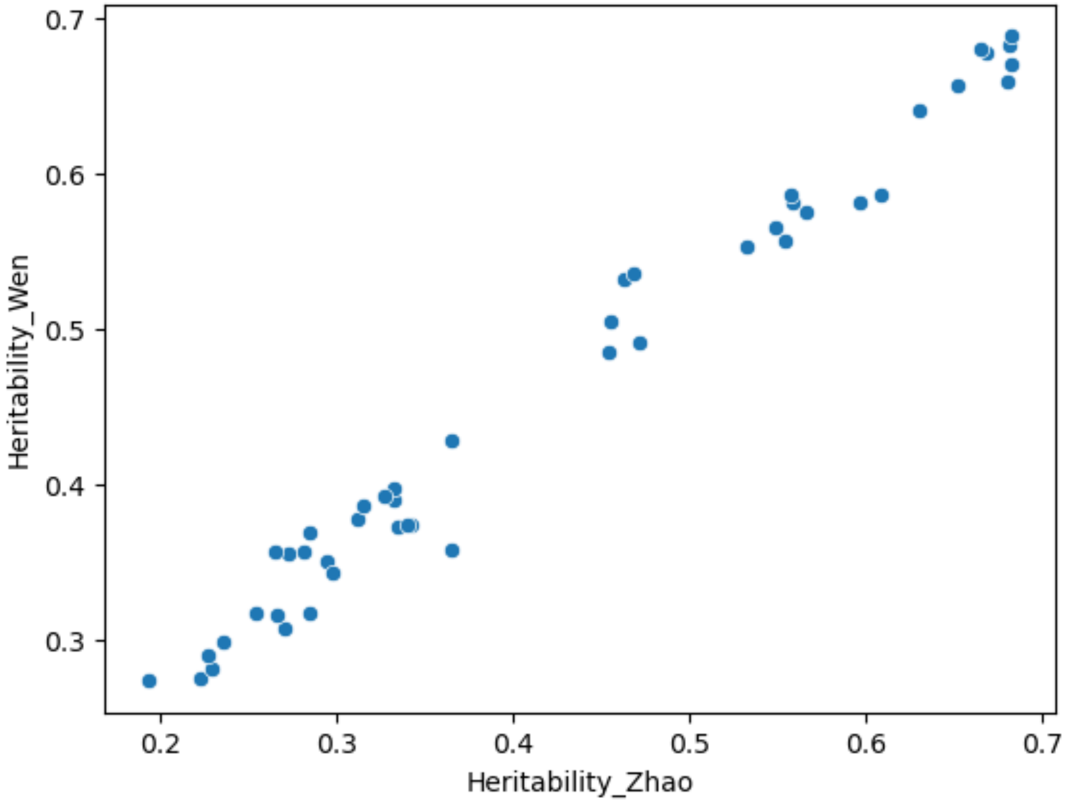


For the 82 heart IDPs (**a**) and 46 eye IDPs (**b**) between our GWAS and the GWAS conducted by Zhao et al., the two sets of SNP-based heritability were highly correlated.

**eFigure 4: Sensitivity check analysis for causal relationship from C512_352 to I9_HEARTFAILURE**

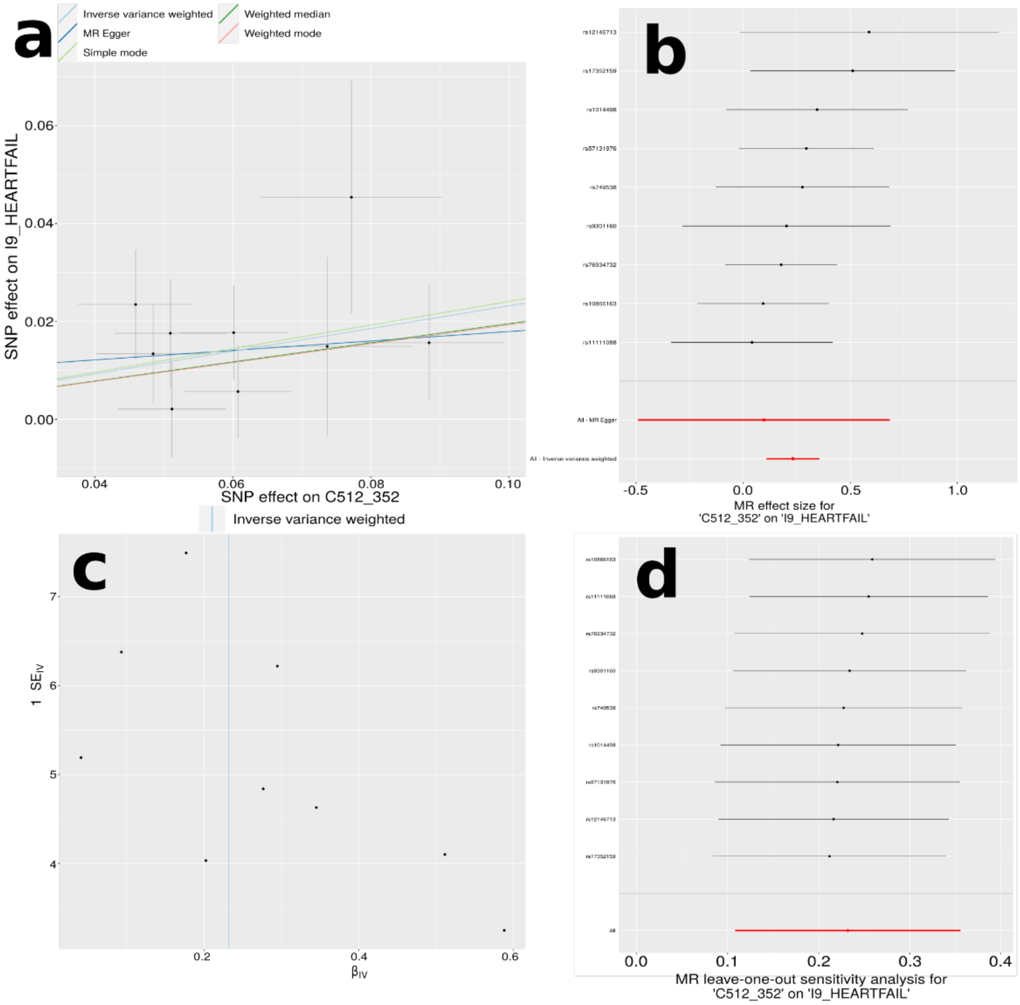


**a**) Scatter plot for the MR effect sizes of the exposure variable (*x*-axis, SD units) and the outcome variable (*y*-axis, log OR) with standard error bars. The slopes of the regression line correspond to the causal effect sizes estimated by the IVW estimator. **b**) Forest plot for the single-SNP MR results. Each line represents the MR effect (log OR) for the exposure variable on the outcome variable using only one SNP; the red line shows the MR effect using all SNPs together. **c**) Funnel plot for the relationship between the causal effect of the exposure variable on the outcome variable. Each dot represents MR effect sizes estimated using each SNP as a separate instrument against the inverse of the standard error of the causal estimate. **d**) Leave-one-out analysis of the exposure variable on the outcome variable. Each row represents the MR effect (log OR) and the 95% CI by excluding that SNP from the analysis. The red line depicts the IVW estimator using all SNPs.

**eFigure 5: Sensitivity check analysis for causal relationship from C256_7 to I9_VARICVE**

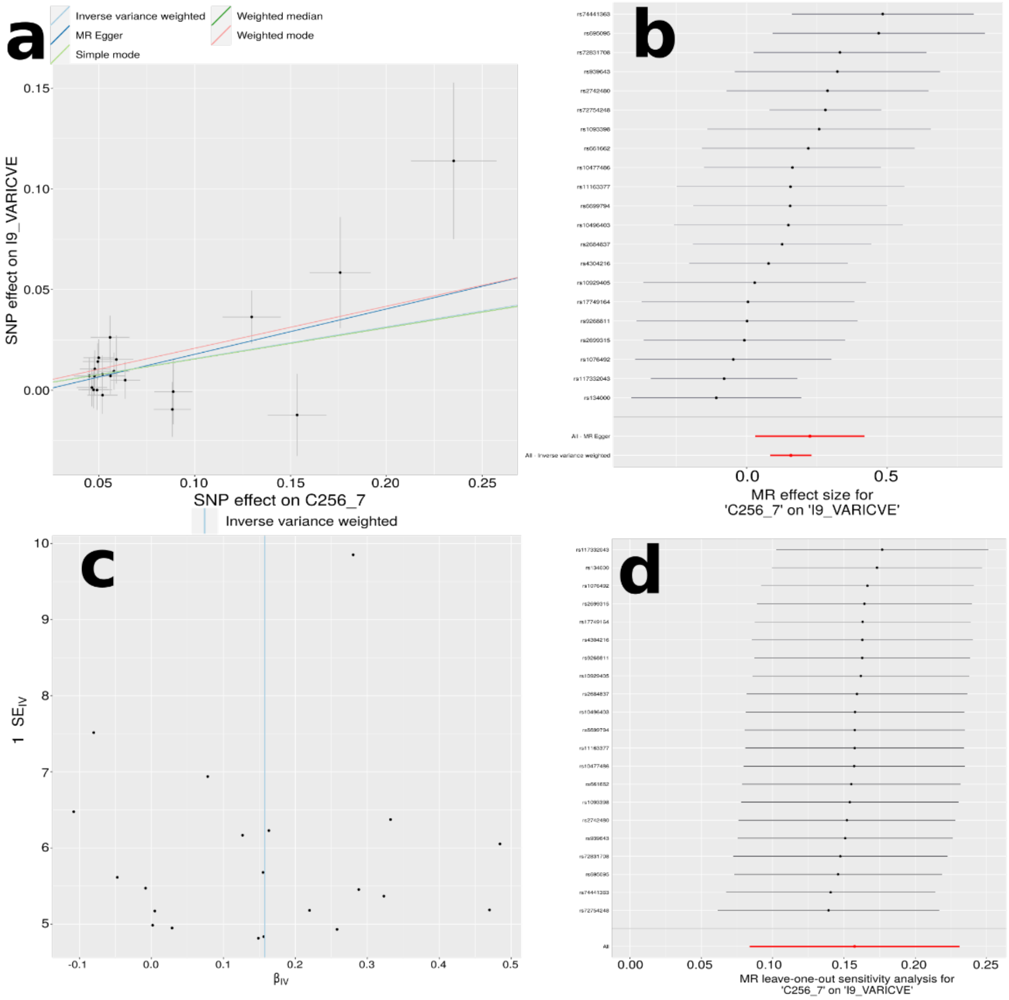


**a**) Scatter plot for the MR effect sizes of the exposure variable (*x*-axis, SD units) and the outcome variable (*y*-axis, log OR) with standard error bars. The slopes of the regression line correspond to the causal effect sizes estimated by the IVW estimator. **b**) Forest plot for the single-SNP MR results. Each line represents the MR effect (log OR) for the exposure variable on the outcome variable using only one SNP; the red line shows the MR effect using all SNPs together. **c**) Funnel plot for the relationship between the causal effect of the exposure variable on the outcome variable. Each dot represents MR effect sizes estimated using each SNP as a separate instrument against the inverse of the standard error of the causal estimate. **d**) Leave-one-out analysis of the exposure variable on the outcome variable. Each row represents the MR effect (log OR) and the 95% CI by excluding that SNP from the analysis. The red line depicts the IVW estimator using all SNPs.

**eFigure 6: Sensitivity check analysis for causal relationship from C1024_726 to I9_DISVEINLYMPH**

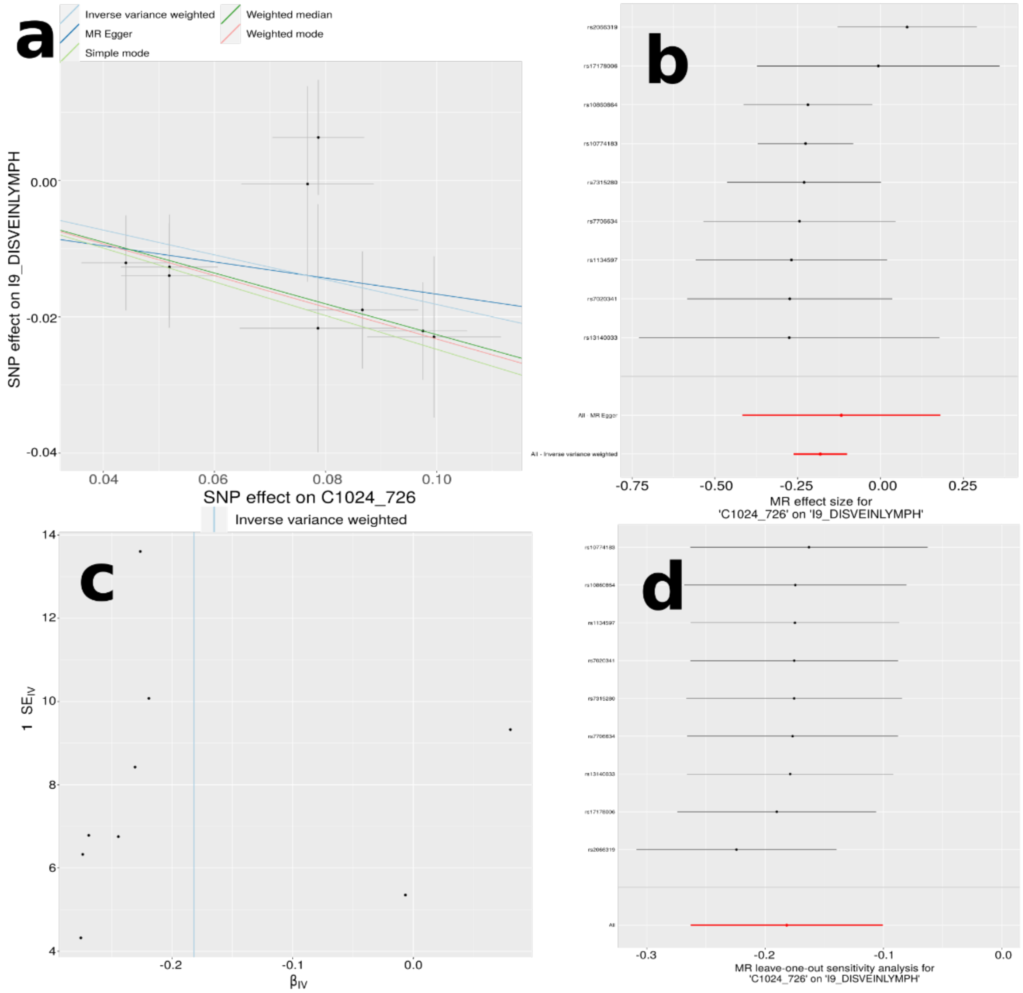


**a**) Scatter plot for the MR effect sizes of the exposure variable (*x*-axis, SD units) and the outcome variable (*y*-axis, log OR) with standard error bars. The slopes of the regression line correspond to the causal effect sizes estimated by the IVW estimator. **b**) Forest plot for the single-SNP MR results. Each line represents the MR effect (log OR) for the exposure variable on the outcome variable using only one SNP; the red line shows the MR effect using all SNPs together. **c**) Funnel plot for the relationship between the causal effect of the exposure variable on the outcome variable. Each dot represents MR effect sizes estimated using each SNP as a separate instrument against the inverse of the standard error of the causal estimate. **d**) Leave-one-out analysis of the exposure variable on the outcome variable. Each row represents the MR effect (log OR) and the 95% CI by excluding that SNP from the analysis. The red line depicts the IVW estimator using all SNPs.

**eFigure 7: Sensitivity check analysis for causal relationship from C1024_880 to I9_HYPTENS**

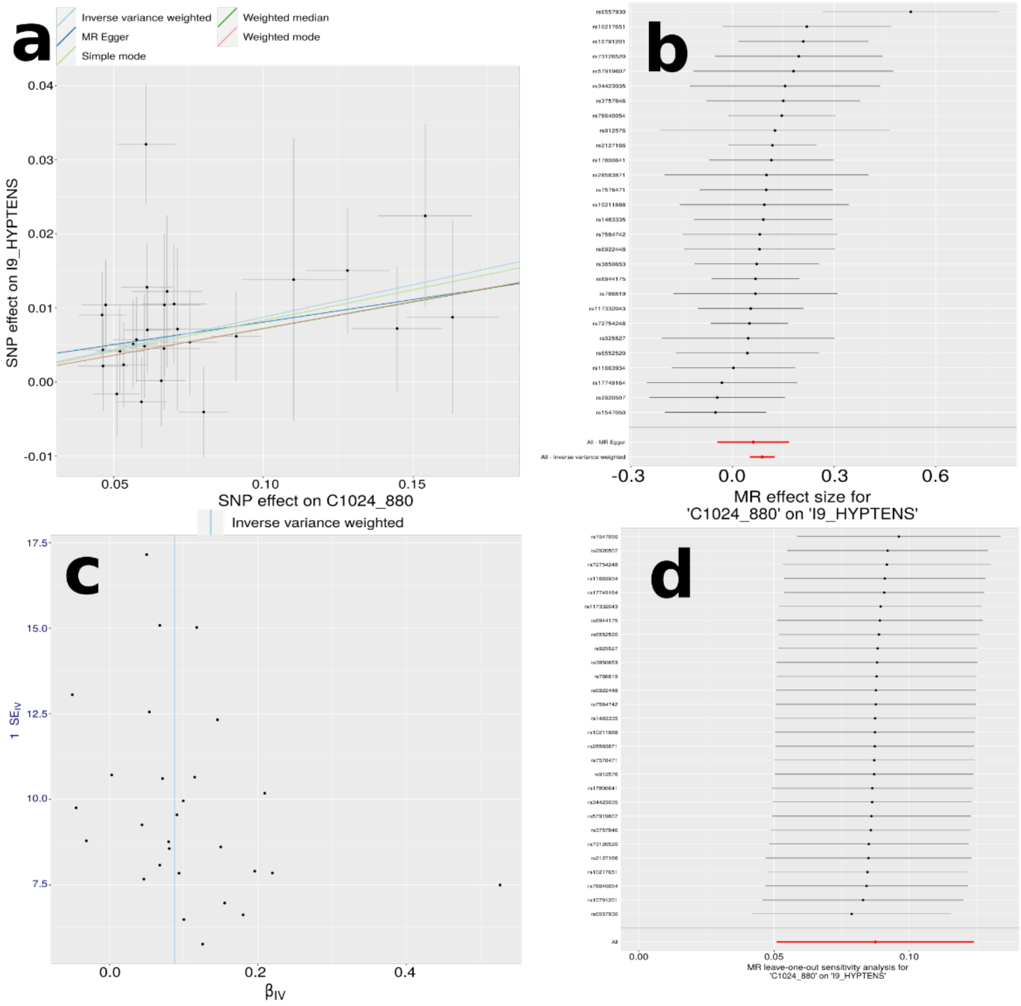


**a**) Scatter plot for the MR effect sizes of the exposure variable (*x*-axis, SD units) and the outcome variable (*y*-axis, log OR) with standard error bars. The slopes of the regression line correspond to the causal effect sizes estimated by the IVW estimator. **b**) Forest plot for the single-SNP MR results. Each line represents the MR effect (log OR) for the exposure variable on the outcome variable using only one SNP; the red line shows the MR effect using all SNPs together. **c**) Funnel plot for the relationship between the causal effect of the exposure variable on the outcome variable. Each dot represents MR effect sizes estimated using each SNP as a separate instrument against the inverse of the standard error of the causal estimate. **d**) Leave-one-out analysis of the exposure variable on the outcome variable. Each row represents the MR effect (log OR) and the 95% CI by excluding that SNP from the analysis. The red line depicts the IVW estimator using all SNPs.

**eFigure 8: Sensitivity check analysis for causal relationship from lv_circumferential_strai_global to F5_DEPRESSION_RECURRENT**

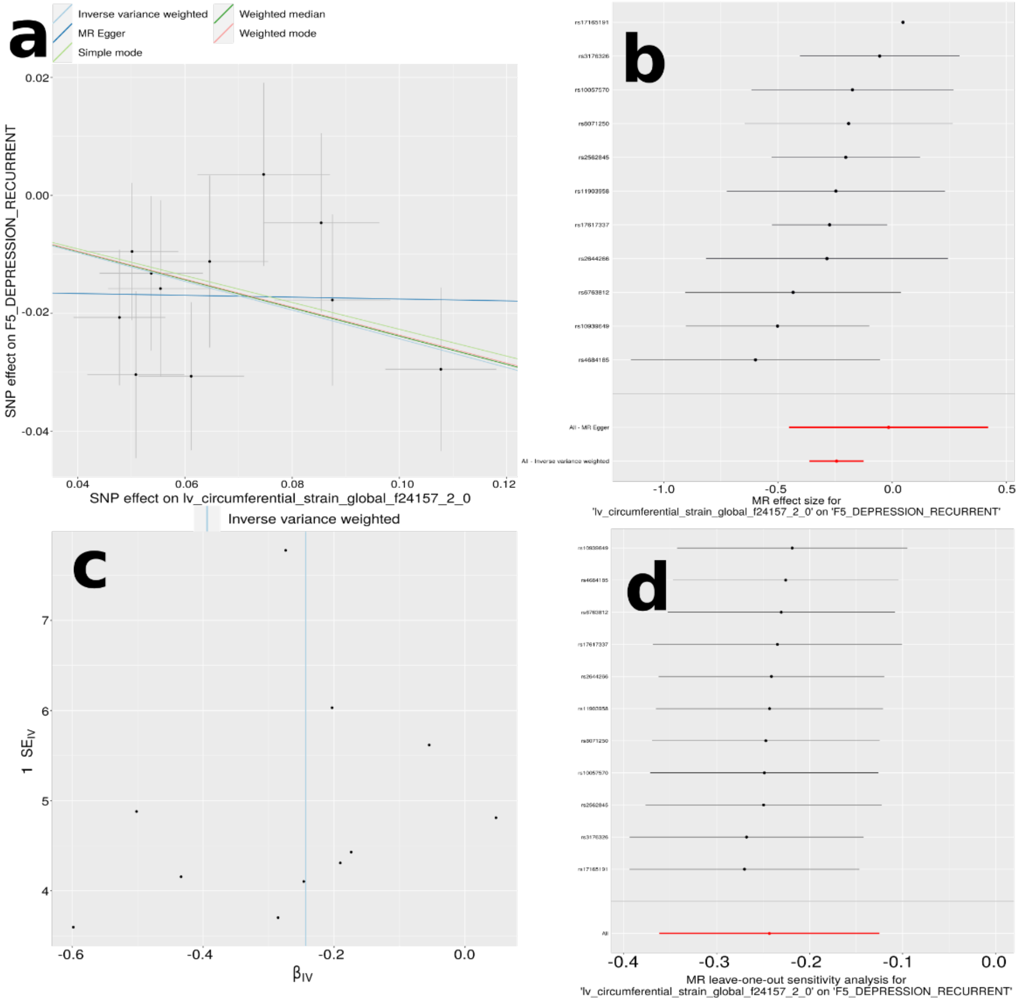


**a**) Scatter plot for the MR effect sizes of the exposure variable (*x*-axis, SD units) and the outcome variable (*y*-axis, log OR) with standard error bars. The slopes of the regression line correspond to the causal effect sizes estimated by the IVW estimator. **b**) Forest plot for the single-SNP MR results. Each line represents the MR effect (log OR) for the exposure variable on the outcome variable using only one SNP; the red line shows the MR effect using all SNPs together. **c**) Funnel plot for the relationship between the causal effect of the exposure variable on the outcome variable. Each dot represents MR effect sizes estimated using each SNP as a separate instrument against the inverse of the standard error of the causal estimate. **d**) Leave-one-out analysis of the exposure variable on the outcome variable. Each row represents the MR effect (log OR) and the 95% CI by excluding that SNP from the analysis. The red line depicts the IVW estimator using all SNPs.

**eFigure 9: Sensitivity check analysis for causal relationship from descending_aorta_minimum_area to G6_MIGRANE_WITH_AURA**

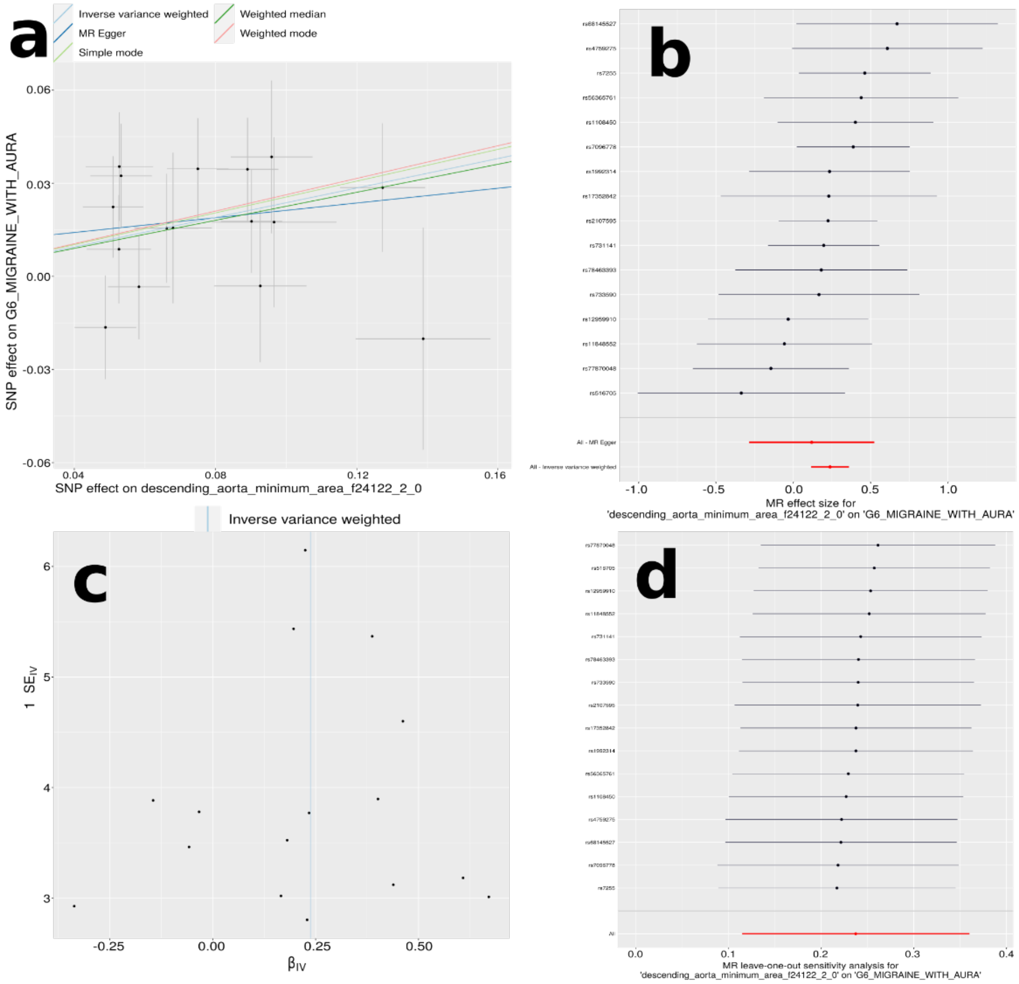


**a**) Scatter plot for the MR effect sizes of the exposure variable (*x*-axis, SD units) and the outcome variable (*y*-axis, log OR) with standard error bars. The slopes of the regression line correspond to the causal effect sizes estimated by the IVW estimator. **b**) Forest plot for the single-SNP MR results. Each line represents the MR effect (log OR) for the exposure variable on the outcome variable using only one SNP; the red line shows the MR effect using all SNPs together. **c**) Funnel plot for the relationship between the causal effect of the exposure variable on the outcome variable. Each dot represents MR effect sizes estimated using each SNP as a separate instrument against the inverse of the standard error of the causal estimate. **d**) Leave-one-out analysis of the exposure variable on the outcome variable. Each row represents the MR effect (log OR) and the 95% CI by excluding that SNP from the analysis. The red line depicts the IVW estimator using all SNPs.

**eFigure 10: Sensitivity check analysis for causal relationship from descending_aorta_maximum_area to G6_MIGRANE_WITH_AURA**

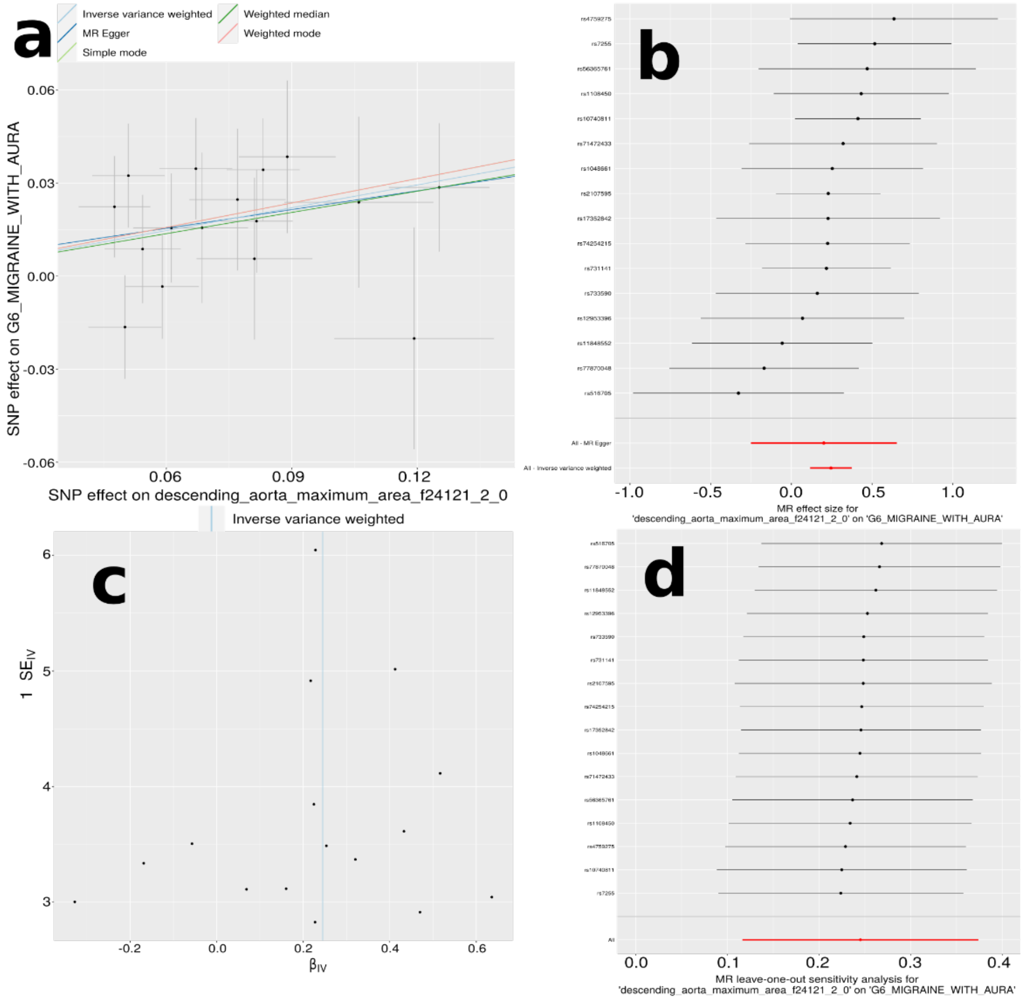


**a**) Scatter plot for the MR effect sizes of the exposure variable (*x*-axis, SD units) and the outcome variable (*y*-axis, log OR) with standard error bars. The slopes of the regression line correspond to the causal effect sizes estimated by the IVW estimator. **b**) Forest plot for the single-SNP MR results. Each line represents the MR effect (log OR) for the exposure variable on the outcome variable using only one SNP; the red line shows the MR effect using all SNPs together. **c**) Funnel plot for the relationship between the causal effect of the exposure variable on the outcome variable. Each dot represents MR effect sizes estimated using each SNP as a separate instrument against the inverse of the standard error of the causal estimate. **d**) Leave-one-out analysis of the exposure variable on the outcome variable. Each row represents the MR effect (log OR) and the 95% CI by excluding that SNP from the analysis. The red line depicts the IVW estimator using all SNPs.

**eFigure 11: Sensitivity check analysis for causal relationship from global mean of the myocardial wall thickness of LV to G6_ALZHEIMER**

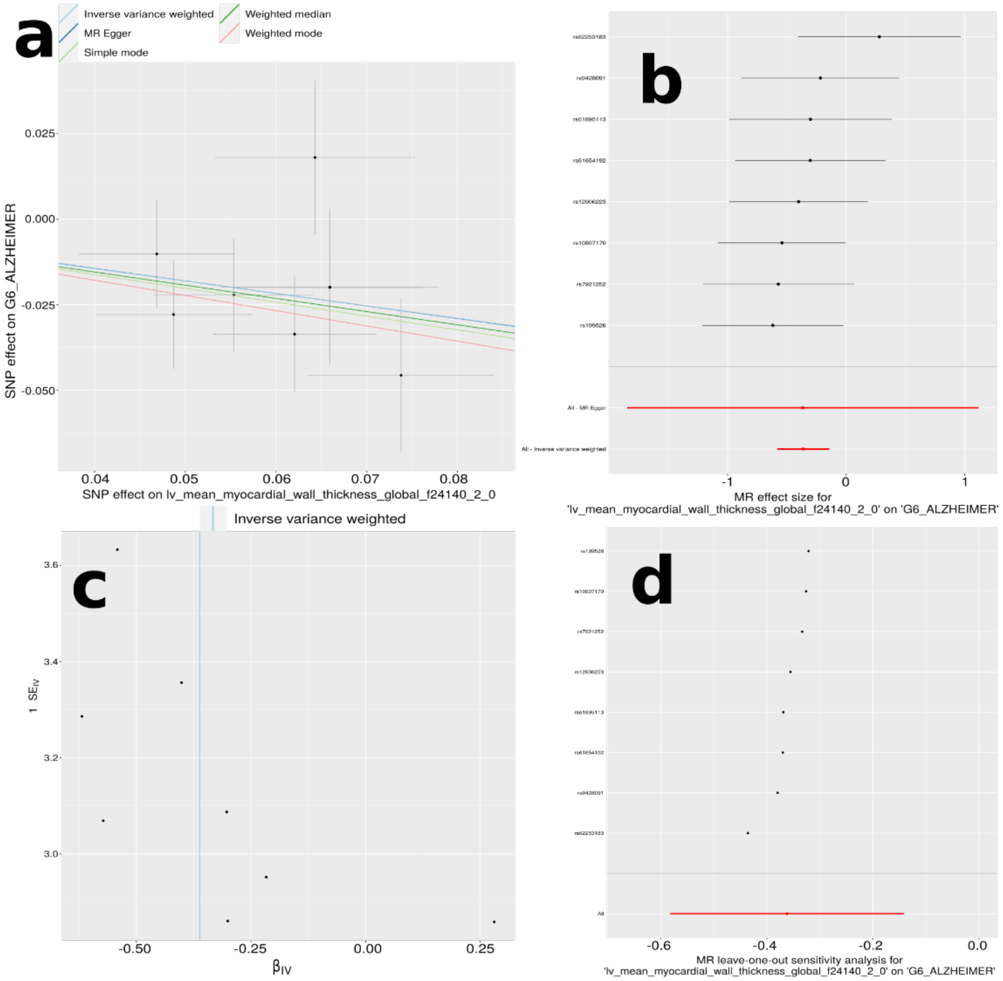


**a**) Scatter plot for the MR effect sizes of the exposure variable (*x*-axis, SD units) and the outcome variable (*y*-axis, log OR) with standard error bars. The slopes of the regression line correspond to the causal effect sizes estimated by the IVW estimator. **b**) Forest plot for the single-SNP MR results. Each line represents the MR effect (log OR) for the exposure variable on the outcome variable using only one SNP; the red line shows the MR effect using all SNPs together. **c**) Funnel plot for the relationship between the causal effect of the exposure variable on the outcome variable. Each dot represents MR effect sizes estimated using each SNP as a separate instrument against the inverse of the standard error of the causal estimate. **d**) Leave-one-out analysis of the exposure variable on the outcome variable. Each row represents the MR effect (log OR) and the 95% CI by excluding that SNP from the analysis. The red line depicts the IVW estimator using all SNPs.

**eFigure 12: Sensitivity check analysis for causal relationship from C512_479 to H7 glaucoma**

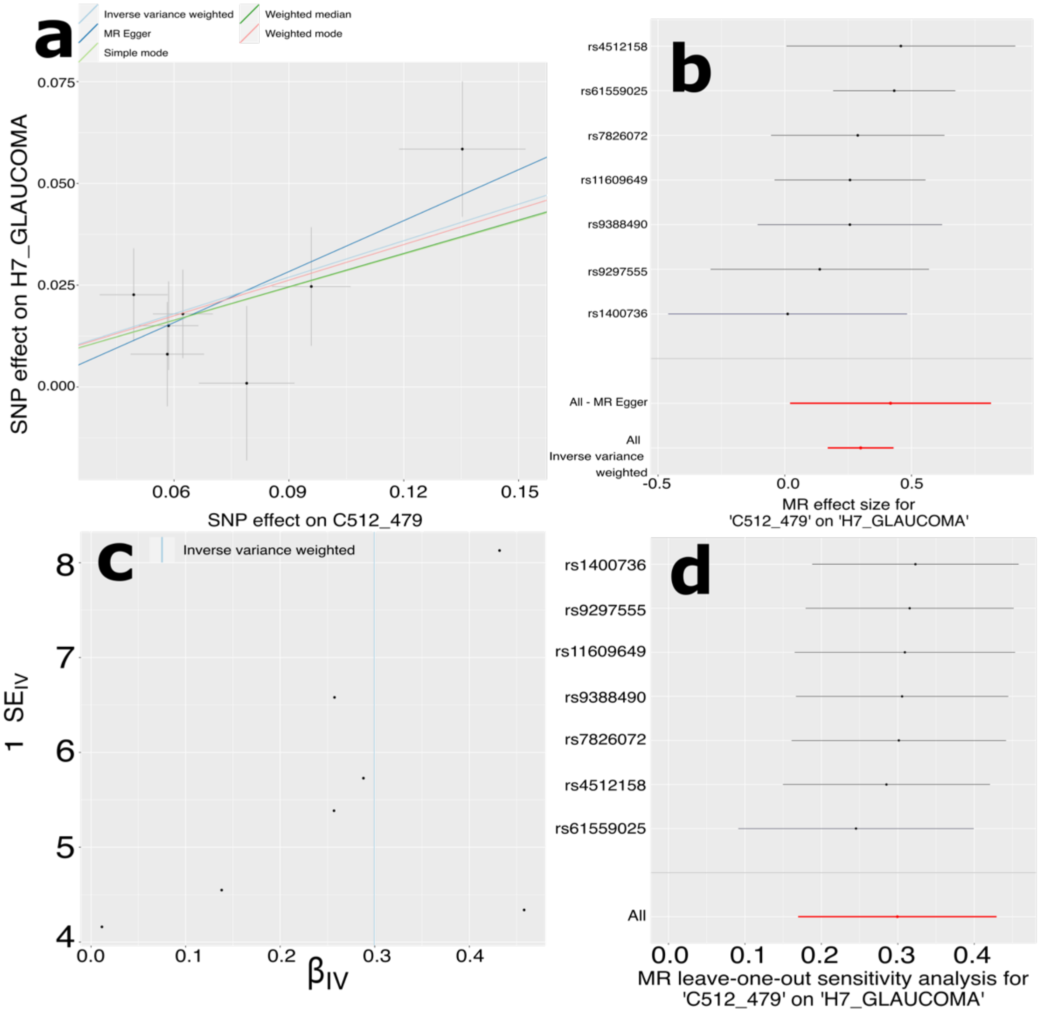


**a**) Scatter plot for the MR effect sizes of the exposure variable (*x*-axis, SD units) and the outcome variable (*y*-axis, log OR) with standard error bars. The slopes of the regression line correspond to the causal effect sizes estimated by the IVW estimator. **b**) Forest plot for the single-SNP MR results. Each line represents the MR effect (log OR) for the exposure variable on the outcome variable using only one SNP; the red line shows the MR effect using all SNPs together. **c**) Funnel plot for the relationship between the causal effect of the exposure variable on the outcome variable. Each dot represents MR effect sizes estimated using each SNP as a separate instrument against the inverse of the standard error of the causal estimate. **d**) Leave-one-out analysis of the exposure variable on the outcome variable. Each row represents the MR effect (log OR) and the 95% CI by excluding that SNP from the analysis. The red line depicts the IVW estimator using all SNPs.

**eFigure 13: Sensitivity check analysis for causal relationship from C512_479 to H7 POAG**

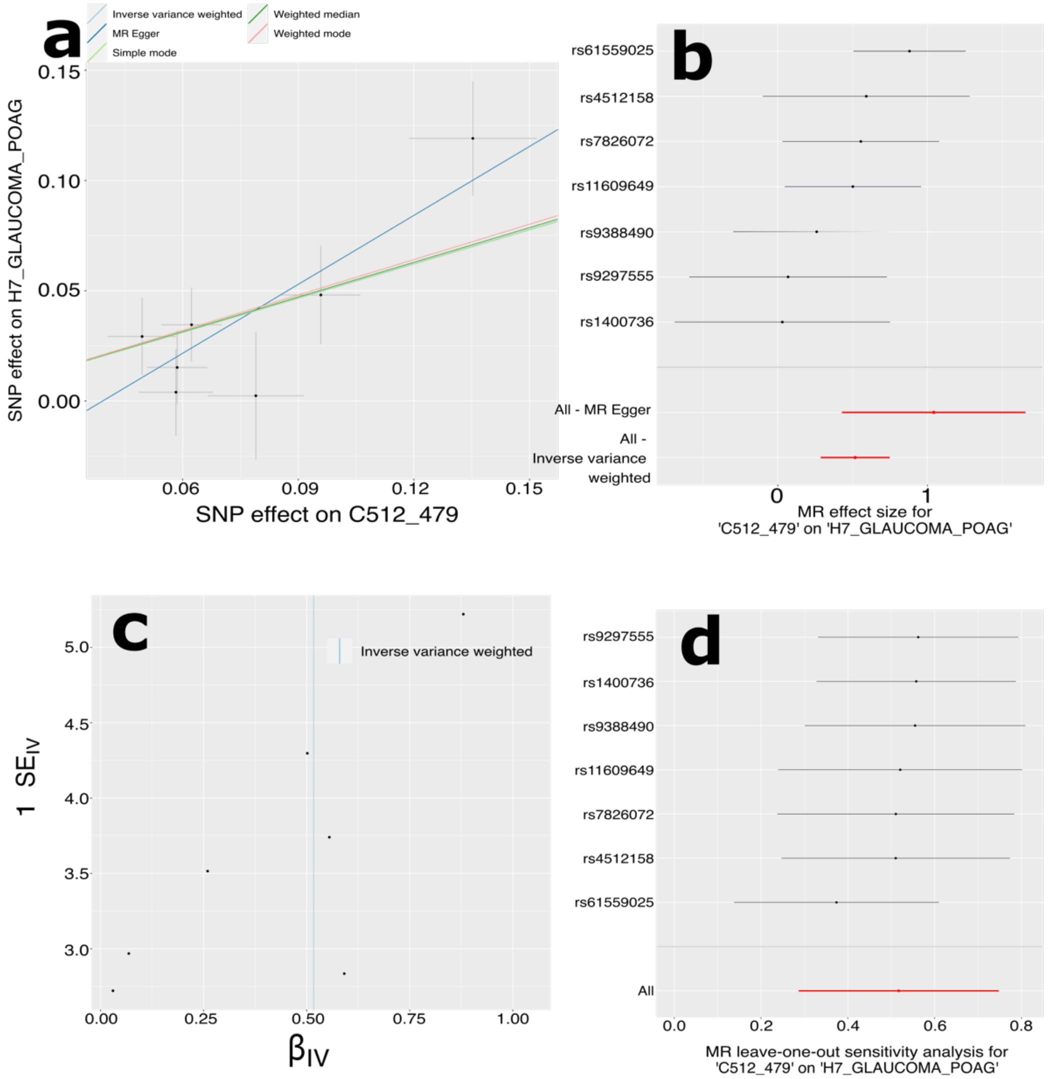


**a**) Scatter plot for the MR effect sizes of the exposure variable (*x*-axis, SD units) and the outcome variable (*y*-axis, log OR) with standard error bars. The slopes of the regression line correspond to the causal effect sizes estimated by the IVW estimator. **b**) Forest plot for the single-SNP MR results. Each line represents the MR effect (log OR) for the exposure variable on the outcome variable using only one SNP; the red line shows the MR effect using all SNPs together. **c**) Funnel plot for the relationship between the causal effect of the exposure variable on the outcome variable. Each dot represents MR effect sizes estimated using each SNP as a separate instrument against the inverse of the standard error of the causal estimate. **d**) Leave-one-out analysis of the exposure variable on the outcome variable. Each row represents the MR effect (log OR) and the 95% CI by excluding that SNP from the analysis. The red line depicts the IVW estimator using all SNPs.

**eFigure 14: Sensitivity check analysis for causal relationship from INLEMLiL to F5 AD**

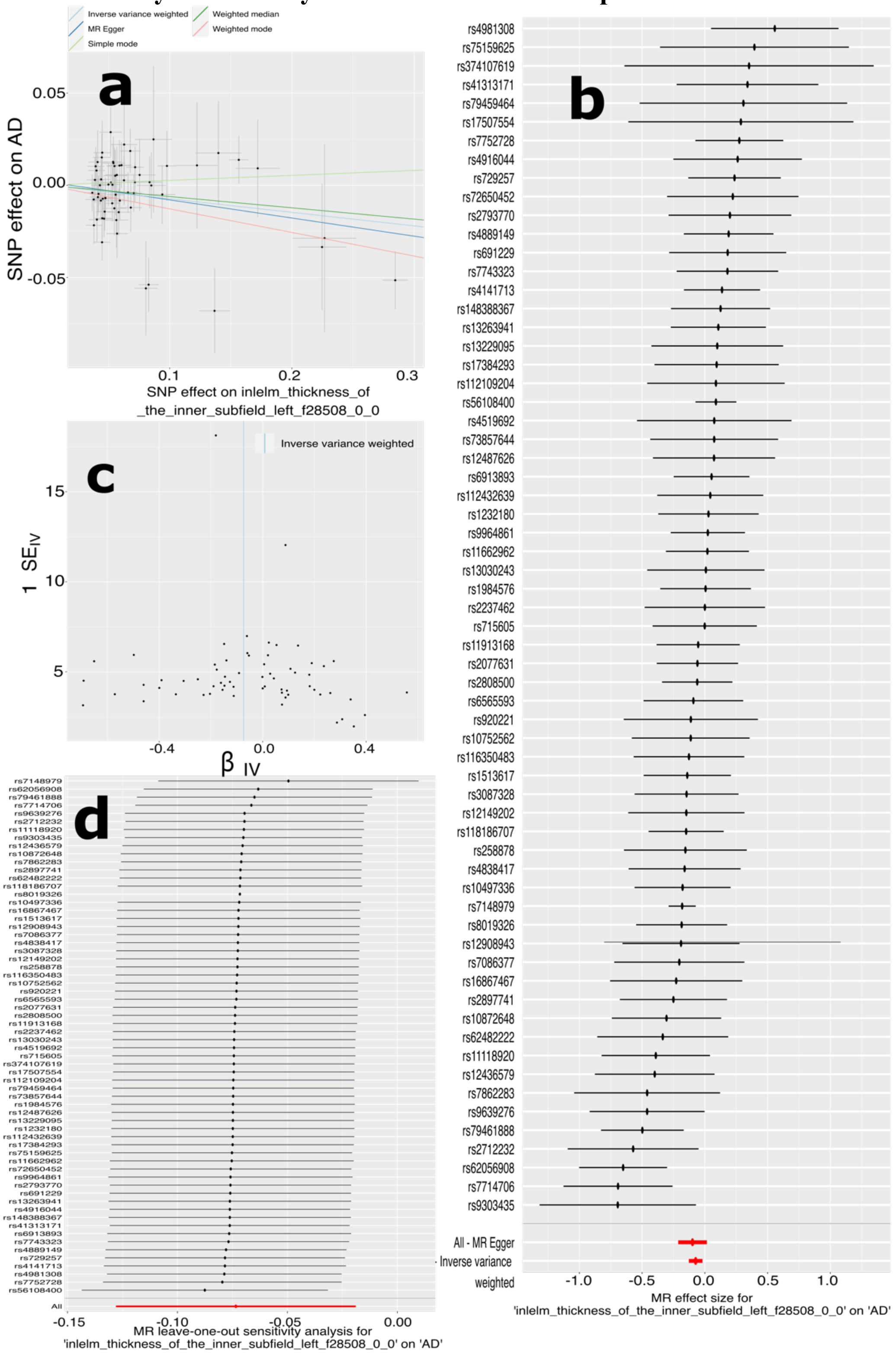


**a**) Scatter plot for the MR effect sizes of the exposure variable (*x*-axis, SD units) and the outcome variable (*y*-axis, log OR) with standard error bars. The slopes of the regression line correspond to the causal effect sizes estimated by the IVW estimator. **b**) Forest plot for the single-SNP MR results. Each line represents the MR effect (log OR) for the exposure variable on the outcome variable using only one SNP; the red line shows the MR effect using all SNPs together. **c**) Funnel plot for the relationship between the causal effect of the exposure variable on the outcome variable. Each dot represents MR effect sizes estimated using each SNP as a separate instrument against the inverse of the standard error of the causal estimate. **d**) Leave-one-out analysis of the exposure variable on the outcome variable. Each row represents the MR effect (log OR) and the 95% CI by excluding that SNP from the analysis. The red line depicts the IVW estimator using all SNPs.

**eFigure 15: Sensitivity check analysis for causal relationship from RpeOsR to F5 the use of hypnotics and sedatives (RX_N05C)**

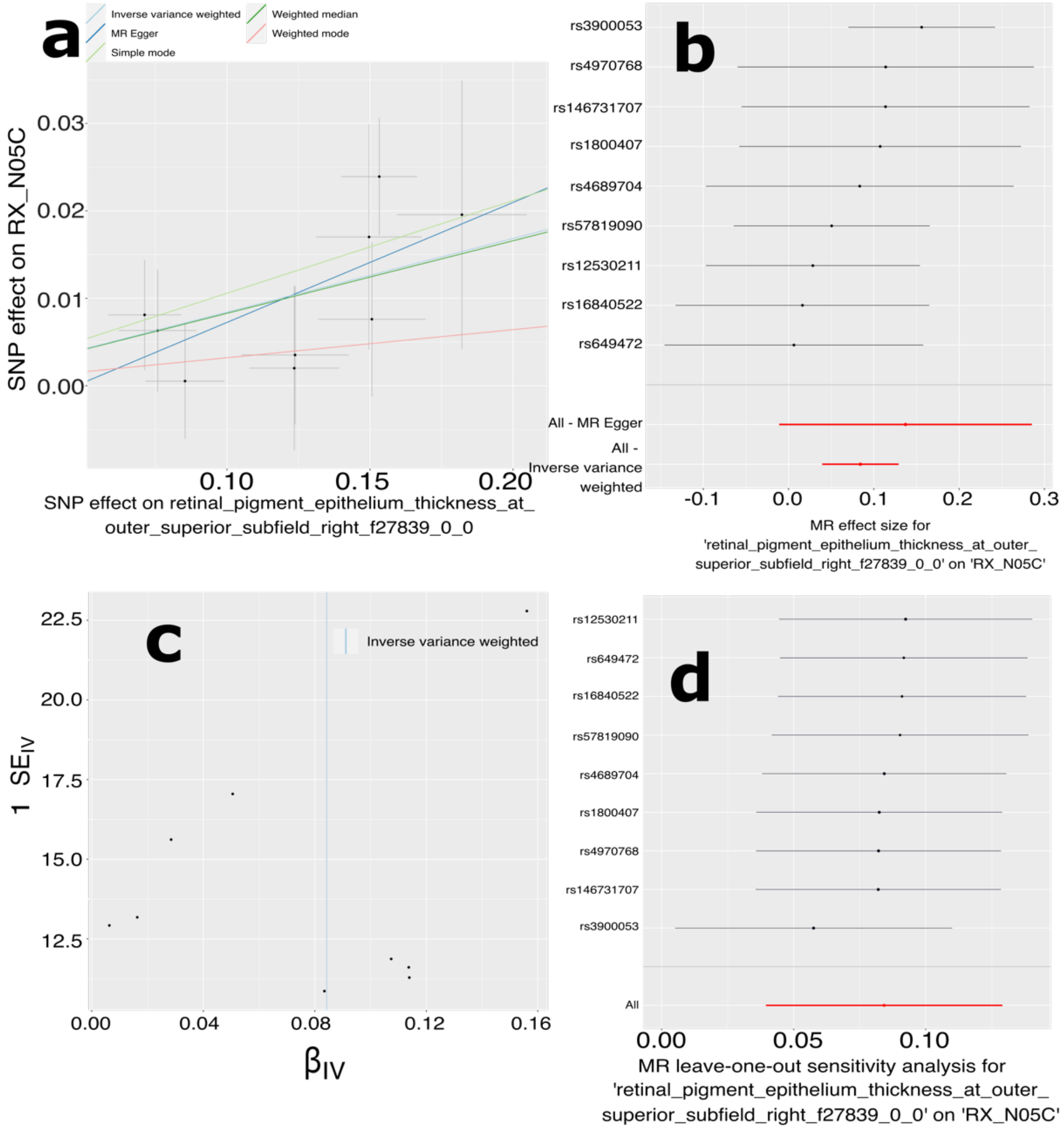


**a**) Scatter plot for the MR effect sizes of the exposure variable (*x*-axis, SD units) and the outcome variable (*y*-axis, log OR) with standard error bars. The slopes of the regression line correspond to the causal effect sizes estimated by the IVW estimator. **b**) Forest plot for the single-SNP MR results. Each line represents the MR effect (log OR) for the exposure variable on the outcome variable using only one SNP; the red line shows the MR effect using all SNPs together. **c**) Funnel plot for the relationship between the causal effect of the exposure variable on the outcome variable. Each dot represents MR effect sizes estimated using each SNP as a separate instrument against the inverse of the standard error of the causal estimate. **d**) Leave-one-out analysis of the exposure variable on the outcome variable. Each row represents the MR effect (log OR) and the 95% CI by excluding that SNP from the analysis. The red line depicts the IVW estimator using all SNPs.

**eFigure 16: Sensitivity check analysis for causal relationship from MisL to F5 depression**

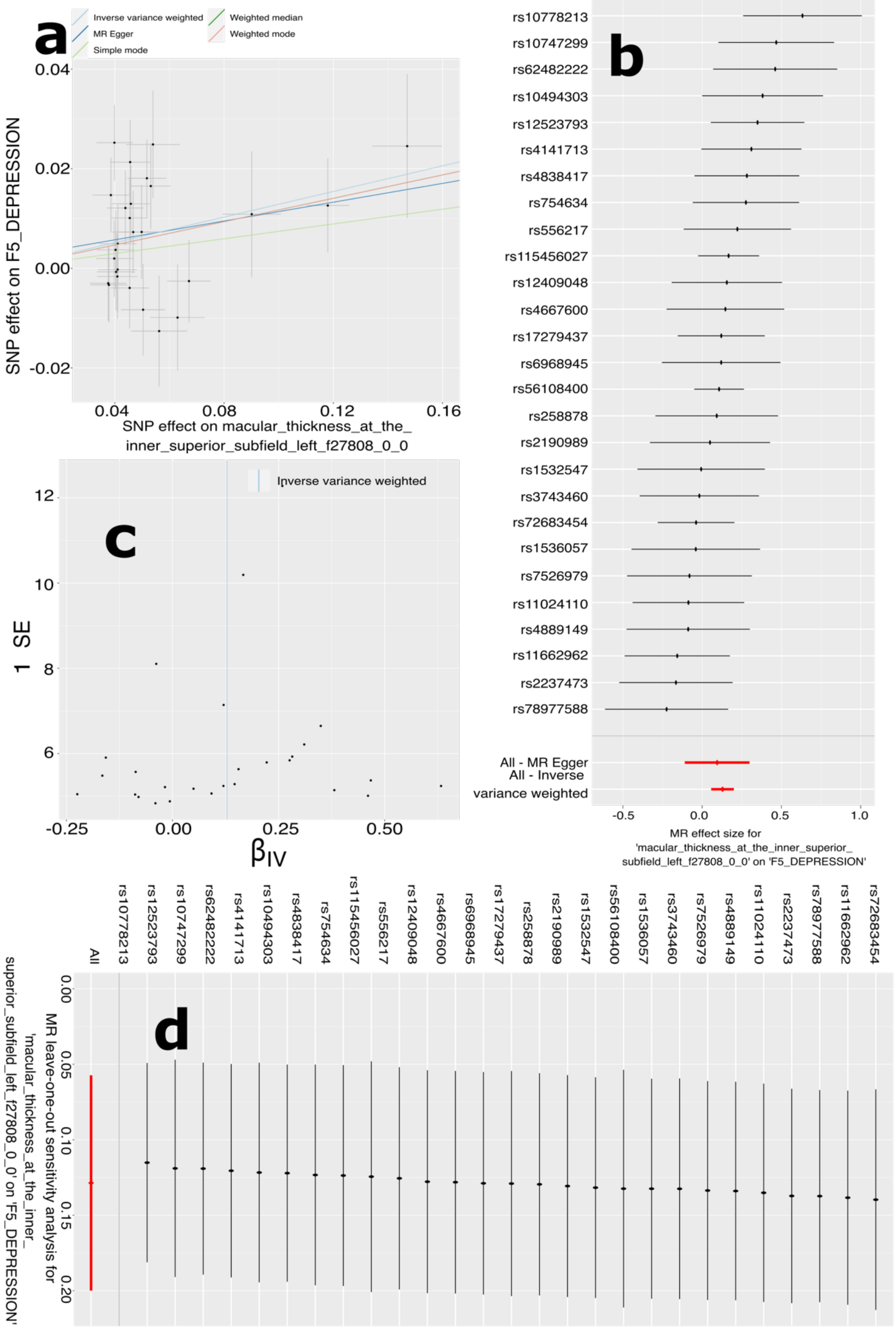


**a**) Scatter plot for the MR effect sizes of the exposure variable (*x*-axis, SD units) and the outcome variable (*y*-axis, log OR) with standard error bars. The slopes of the regression line correspond to the causal effect sizes estimated by the IVW estimator. **b**) Forest plot for the single-SNP MR results. Each line represents the MR effect (log OR) for the exposure variable on the outcome variable using only one SNP; the red line shows the MR effect using all SNPs together. **c**) Funnel plot for the relationship between the causal effect of the exposure variable on the outcome variable. Each dot represents MR effect sizes estimated using each SNP as a separate instrument against the inverse of the standard error of the causal estimate. **d**) Leave-one-out analysis of the exposure variable on the outcome variable. Each row represents the MR effect (log OR) and the 95% CI by excluding that SNP from the analysis. The red line depicts the IVW estimator using all SNPs.

**eFigure 17: Sensitivity check analysis for causal relationship from MiiL to F5 sleep apnea**

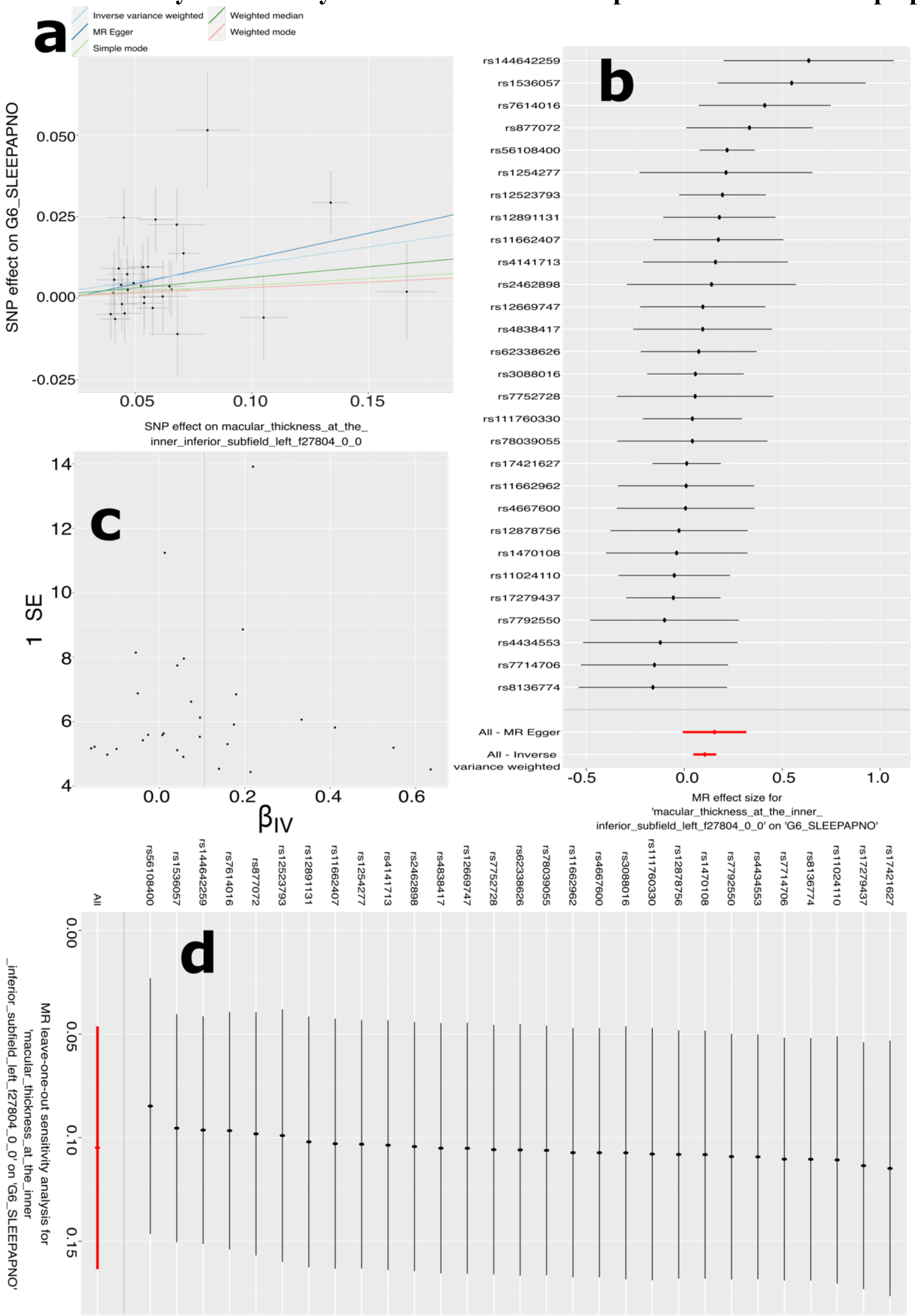


**a**) Scatter plot for the MR effect sizes of the exposure variable (*x*-axis, SD units) and the outcome variable (*y*-axis, log OR) with standard error bars. The slopes of the regression line correspond to the causal effect sizes estimated by the IVW estimator. **b**) Forest plot for the single-SNP MR results. Each line represents the MR effect (log OR) for the exposure variable on the outcome variable using only one SNP; the red line shows the MR effect using all SNPs together. **c**) Funnel plot for the relationship between the causal effect of the exposure variable on the outcome variable. Each dot represents MR effect sizes estimated using each SNP as a separate instrument against the inverse of the standard error of the causal estimate. **d**) Leave-one-out analysis of the exposure variable on the outcome variable. Each row represents the MR effect (log OR) and the 95% CI by excluding that SNP from the analysis. The red line depicts the IVW estimator using all SNPs.

**eFigure 18: Sensitivity check analysis for causal relationship from Dao_max to H7 STRABOTH**

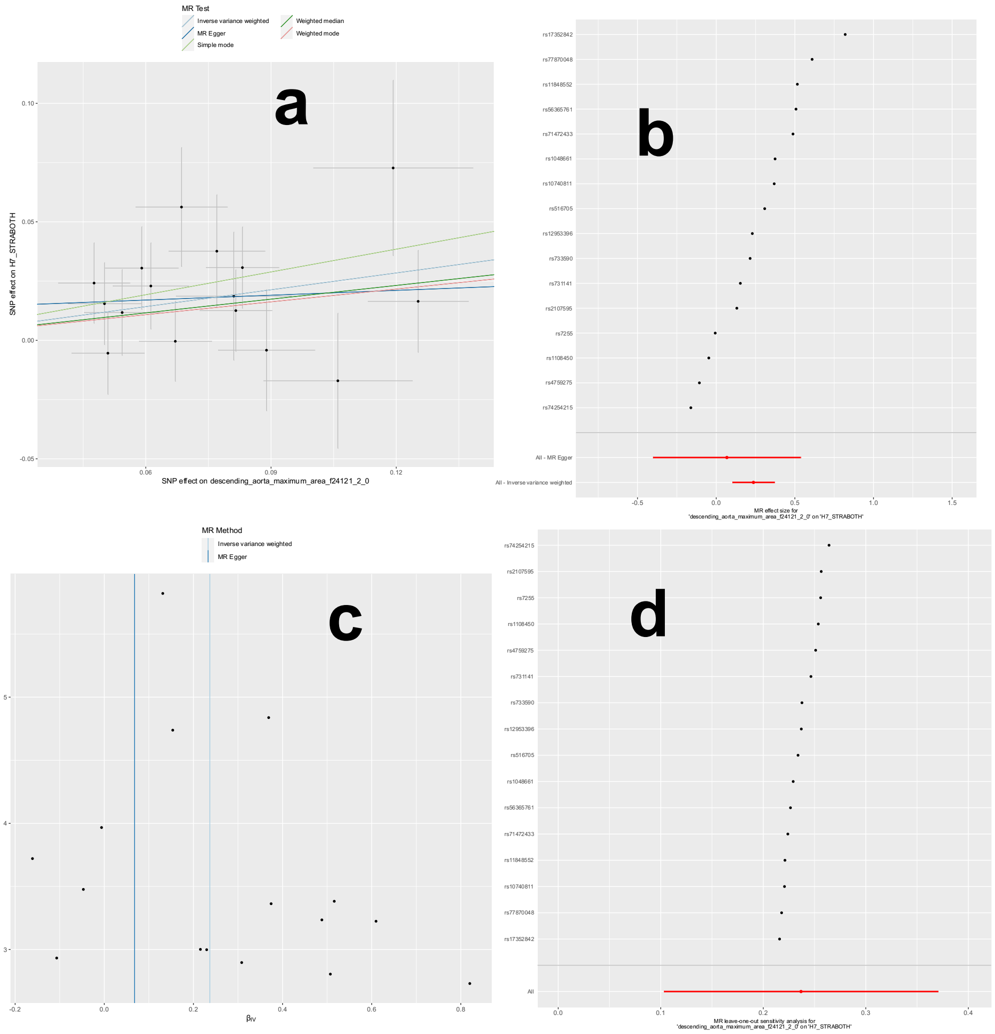


**a**) Scatter plot for the MR effect sizes of the exposure variable (*x*-axis, SD units) and the outcome variable (*y*-axis, log OR) with standard error bars. The slopes of the regression line correspond to the causal effect sizes estimated by the IVW estimator. **b**) Forest plot for the single-SNP MR results. Each line represents the MR effect (log OR) for the exposure variable on the outcome variable using only one SNP; the red line shows the MR effect using all SNPs together. **c**) Funnel plot for the relationship between the causal effect of the exposure variable on the outcome variable. Each dot represents MR effect sizes estimated using each SNP as a separate instrument against the inverse of the standard error of the causal estimate. **d**) Leave-one-out analysis of the exposure variable on the outcome variable. Each row represents the MR effect (log OR) and the 95% CI by excluding that SNP from the analysis. The red line depicts the IVW estimator using all SNPs.

**eFigure 19: Sensitivity check analysis for causal relationship from Dao_max to H7 OCUMUSCLE**

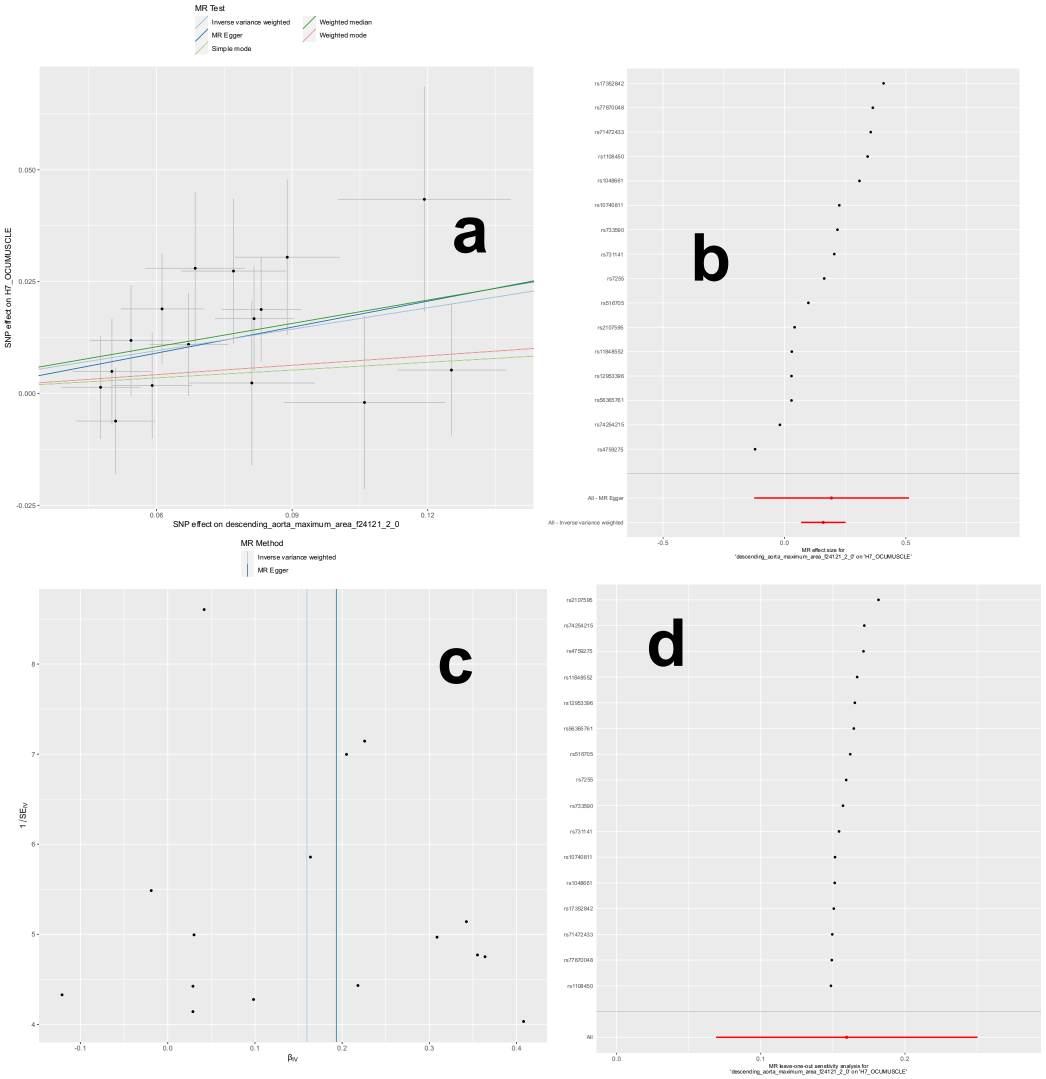


**a**) Scatter plot for the MR effect sizes of the exposure variable (*x*-axis, SD units) and the outcome variable (*y*-axis, log OR) with standard error bars. The slopes of the regression line correspond to the causal effect sizes estimated by the IVW estimator. **b**) Forest plot for the single-SNP MR results. Each line represents the MR effect (log OR) for the exposure variable on the outcome variable using only one SNP; the red line shows the MR effect using all SNPs together. **c**) Funnel plot for the relationship between the causal effect of the exposure variable on the outcome variable. Each dot represents MR effect sizes estimated using each SNP as a separate instrument against the inverse of the standard error of the causal estimate. **d**) Leave-one-out analysis of the exposure variable on the outcome variable. Each row represents the MR effect (log OR) and the 95% CI by excluding that SNP from the analysis. The red line depicts the IVW estimator using all SNPs.

**eFigure 20: Sensitivity check analysis for causal relationship from Dao_min to H7 STRABOTH**

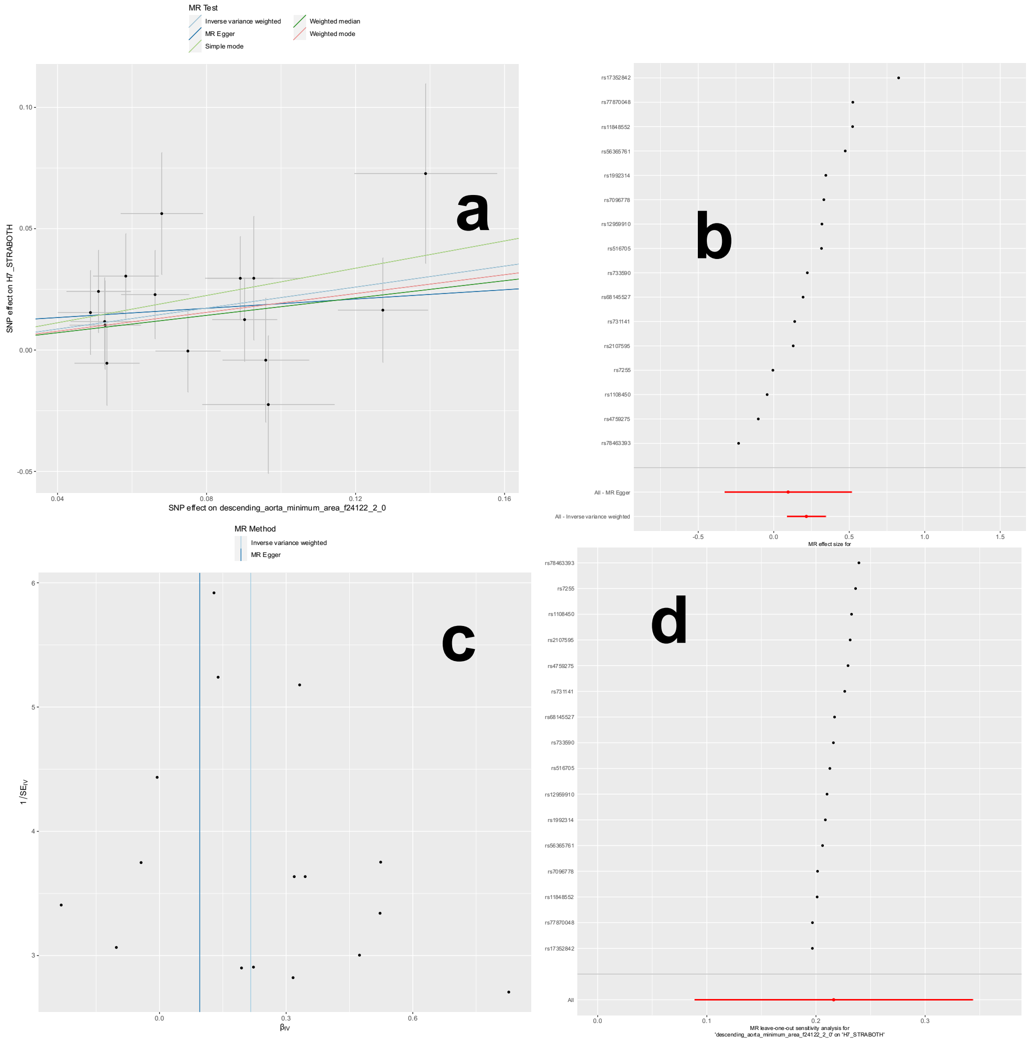


**a**) Scatter plot for the MR effect sizes of the exposure variable (*x*-axis, SD units) and the outcome variable (*y*-axis, log OR) with standard error bars. The slopes of the regression line correspond to the causal effect sizes estimated by the IVW estimator. **b**) Forest plot for the single-SNP MR results. Each line represents the MR effect (log OR) for the exposure variable on the outcome variable using only one SNP; the red line shows the MR effect using all SNPs together. **c**) Funnel plot for the relationship between the causal effect of the exposure variable on the outcome variable. Each dot represents MR effect sizes estimated using each SNP as a separate instrument against the inverse of the standard error of the causal estimate. **d**) Leave-one-out analysis of the exposure variable on the outcome variable. Each row represents the MR effect (log OR) and the 95% CI by excluding that SNP from the analysis. The red line depicts the IVW estimator using all SNPs.

**eFigure 21: Sensitivity check analysis for causal relationship from Dao_min to H7 OCUMUSCLE**

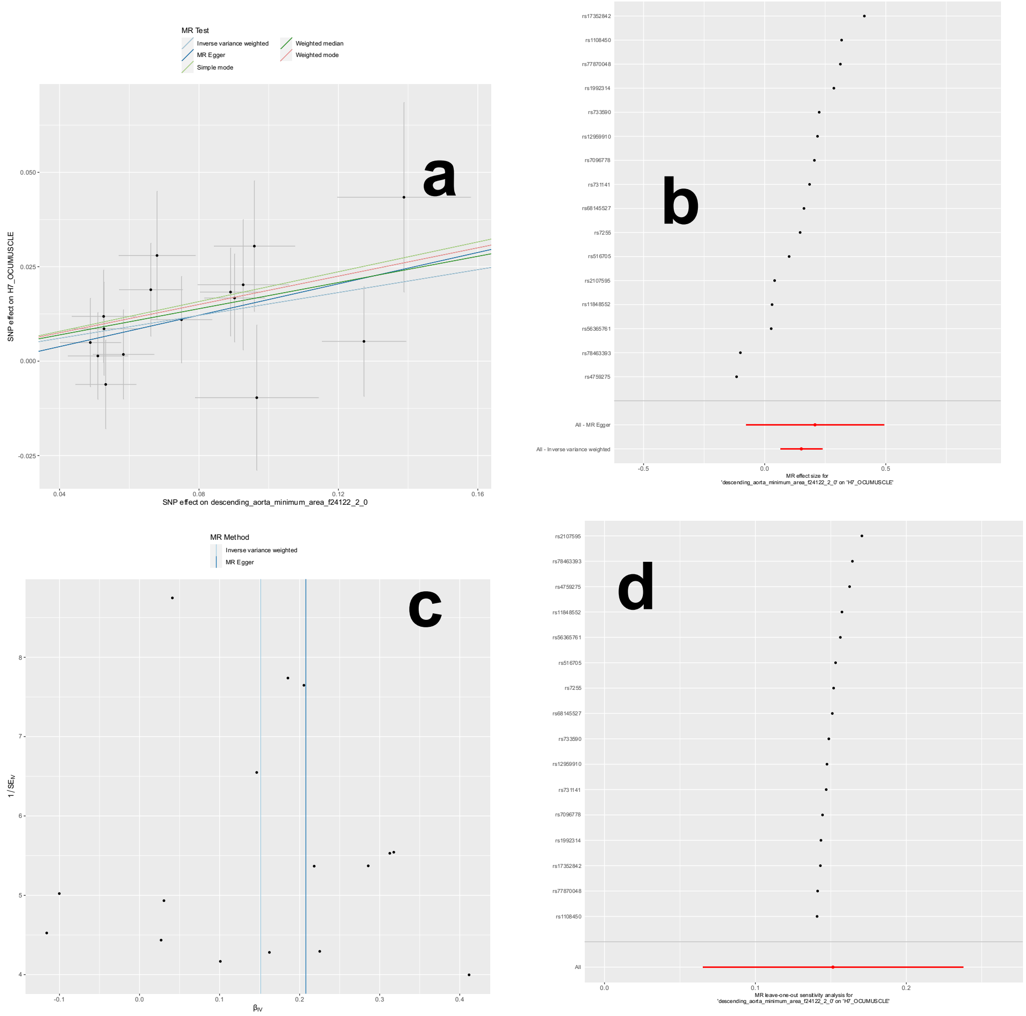


**a**) Scatter plot for the MR effect sizes of the exposure variable (*x*-axis, SD units) and the outcome variable (*y*-axis, log OR) with standard error bars. The slopes of the regression line correspond to the causal effect sizes estimated by the IVW estimator. **b**) Forest plot for the single-SNP MR results. Each line represents the MR effect (log OR) for the exposure variable on the outcome variable using only one SNP; the red line shows the MR effect using all SNPs together. **c**) Funnel plot for the relationship between the causal effect of the exposure variable on the outcome variable. Each dot represents MR effect sizes estimated using each SNP as a separate instrument against the inverse of the standard error of the causal estimate. **d**) Leave-one-out analysis of the exposure variable on the outcome variable. Each row represents the MR effect (log OR) and the 95% CI by excluding that SNP from the analysis. The red line depicts the IVW estimator using all SNPs.

**eFigure 22: Sensitivity check analysis for causal relationship from Aao_max to H7 GLAUCOMA**

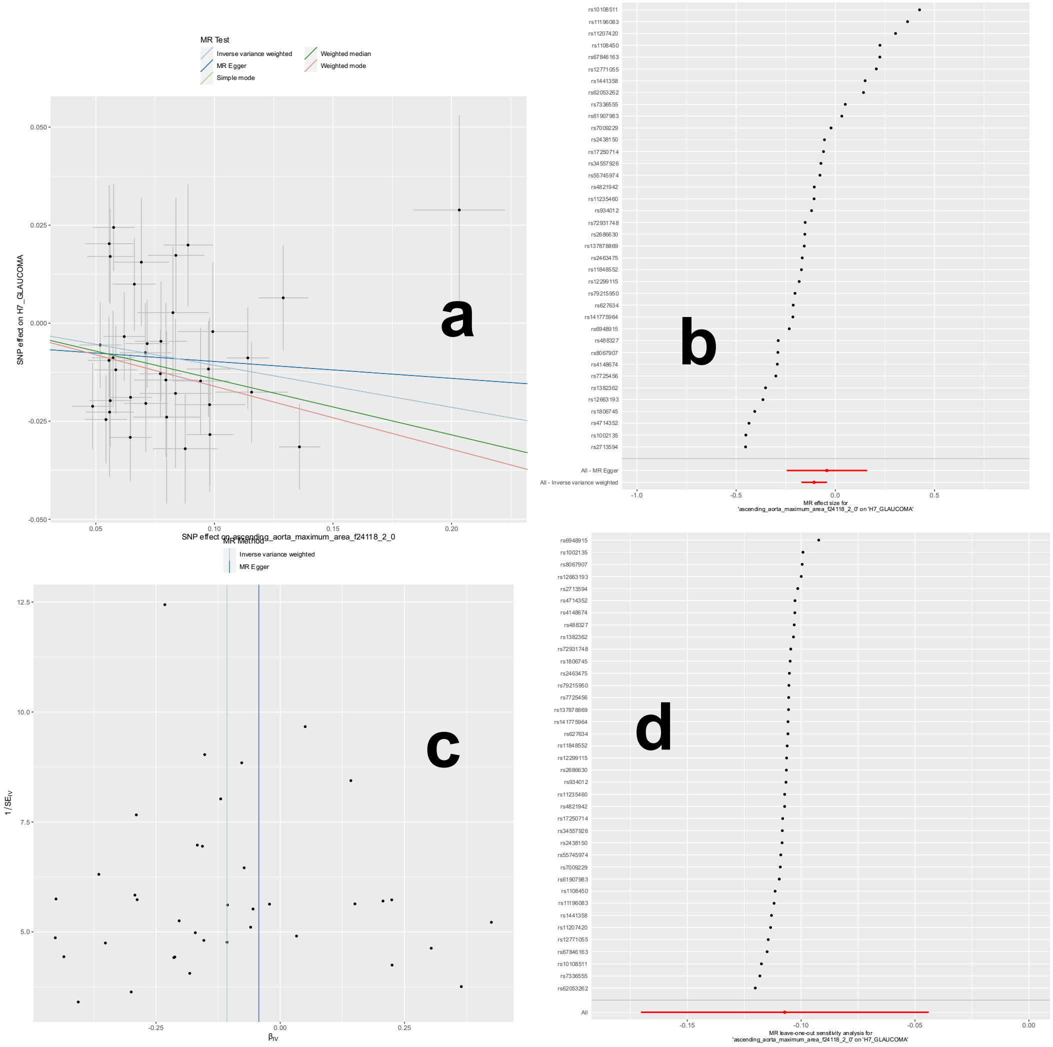


**a**) Scatter plot for the MR effect sizes of the exposure variable (*x*-axis, SD units) and the outcome variable (*y*-axis, log OR) with standard error bars. The slopes of the regression line correspond to the causal effect sizes estimated by the IVW estimator. **b**) Forest plot for the single-SNP MR results. Each line represents the MR effect (log OR) for the exposure variable on the outcome variable using only one SNP; the red line shows the MR effect using all SNPs together. **c**) Funnel plot for the relationship between the causal effect of the exposure variable on the outcome variable. Each dot represents MR effect sizes estimated using each SNP as a separate instrument against the inverse of the standard error of the causal estimate. **d**) Leave-one-out analysis of the exposure variable on the outcome variable. Each row represents the MR effect (log OR) and the 95% CI by excluding that SNP from the analysis. The red line depicts the IVW estimator using all SNPs.

**eFigure 23: Sensitivity check analysis for causal relationship from ElmIsosL to I9 CORATHER**

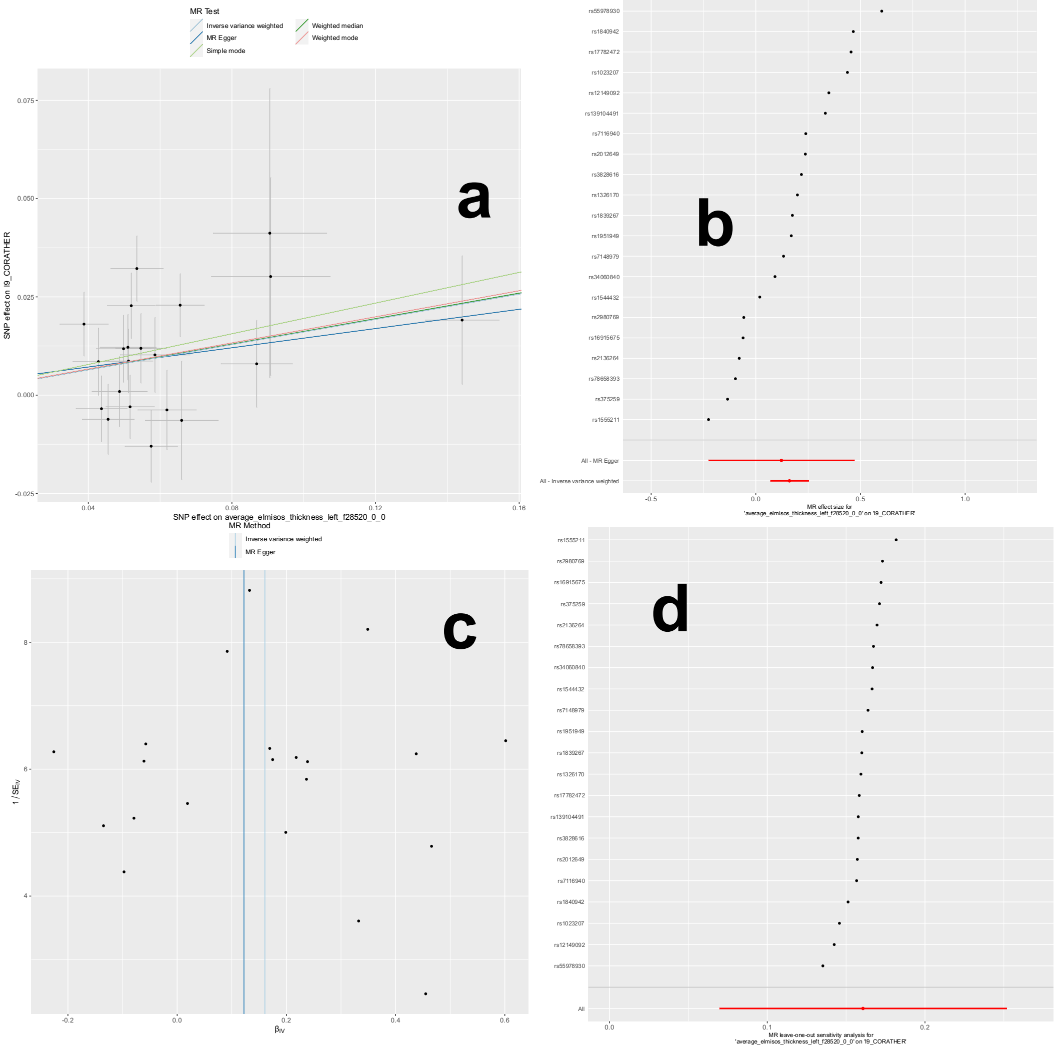


**a**) Scatter plot for the MR effect sizes of the exposure variable (*x*-axis, SD units) and the outcome variable (*y*-axis, log OR) with standard error bars. The slopes of the regression line correspond to the causal effect sizes estimated by the IVW estimator. **b**) Forest plot for the single-SNP MR results. Each line represents the MR effect (log OR) for the exposure variable on the outcome variable using only one SNP; the red line shows the MR effect using all SNPs together. **c**) Funnel plot for the relationship between the causal effect of the exposure variable on the outcome variable. Each dot represents MR effect sizes estimated using each SNP as a separate instrument against the inverse of the standard error of the causal estimate. **d**) Leave-one-out analysis of the exposure variable on the outcome variable. Each row represents the MR effect (log OR) and the 95% CI by excluding that SNP from the analysis. The red line depicts the IVW estimator using all SNPs.

**eFigure 24: Incremental R2 for individual-level imaging features and their respective PRS for predicting the 14 systemic disease categories.**

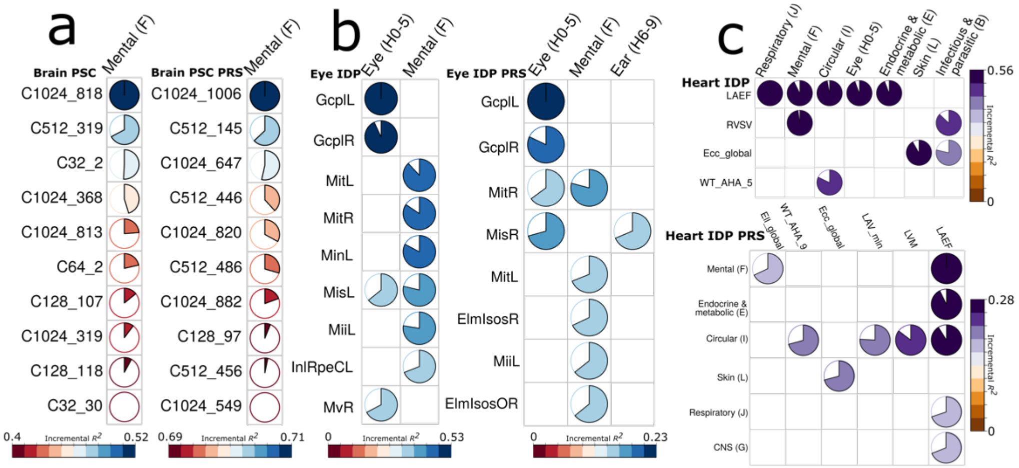


We present the representative features with the 10 highest incremental $R^2$ for each organ.

**eFigure 25: Classification performance for predicting the 14 systemic disease categories using brain PSCs and conventional MUSE ROIs.**

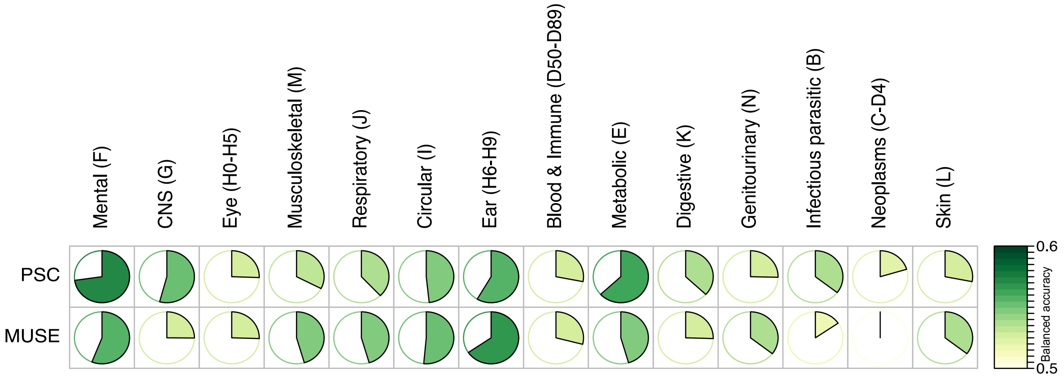


Classification performance to predict the 14 disease categories using data-driven PSCs and conventional MUSE GM ROIs as features.

**eFigure 26: The CV performance from the training/validation/test datasets for predicting the 8 cognitive scores**

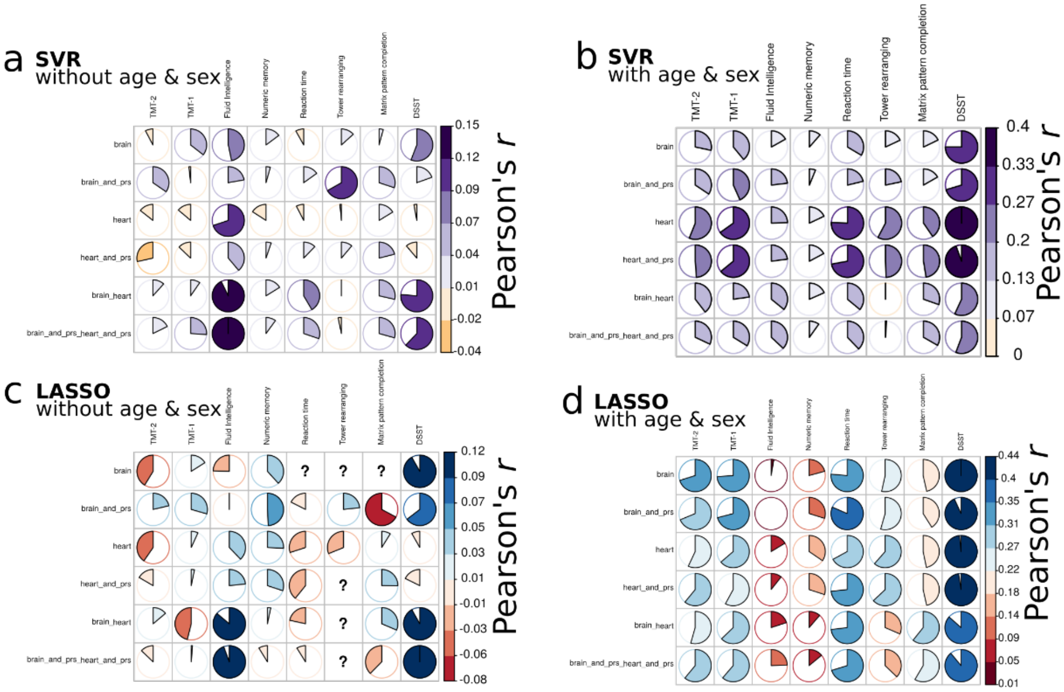


Regression performance using Pearson's *r* in the nested cross-validated training/validation/test datasets for SVR without age and sex as additional features (**a**), SVR with age and sex as additional features (**b**), Lasso regression without age and sex as additional features (**c**), Lasso with age and sex as additional features (**d**). In Figure **c**, the symbol "?" denotes that the model could not predict well for the cognitive scores, resulting in the same predicted values for all instances.

**eFigure 27: The expression analyses of the MOG protein in the HPA**

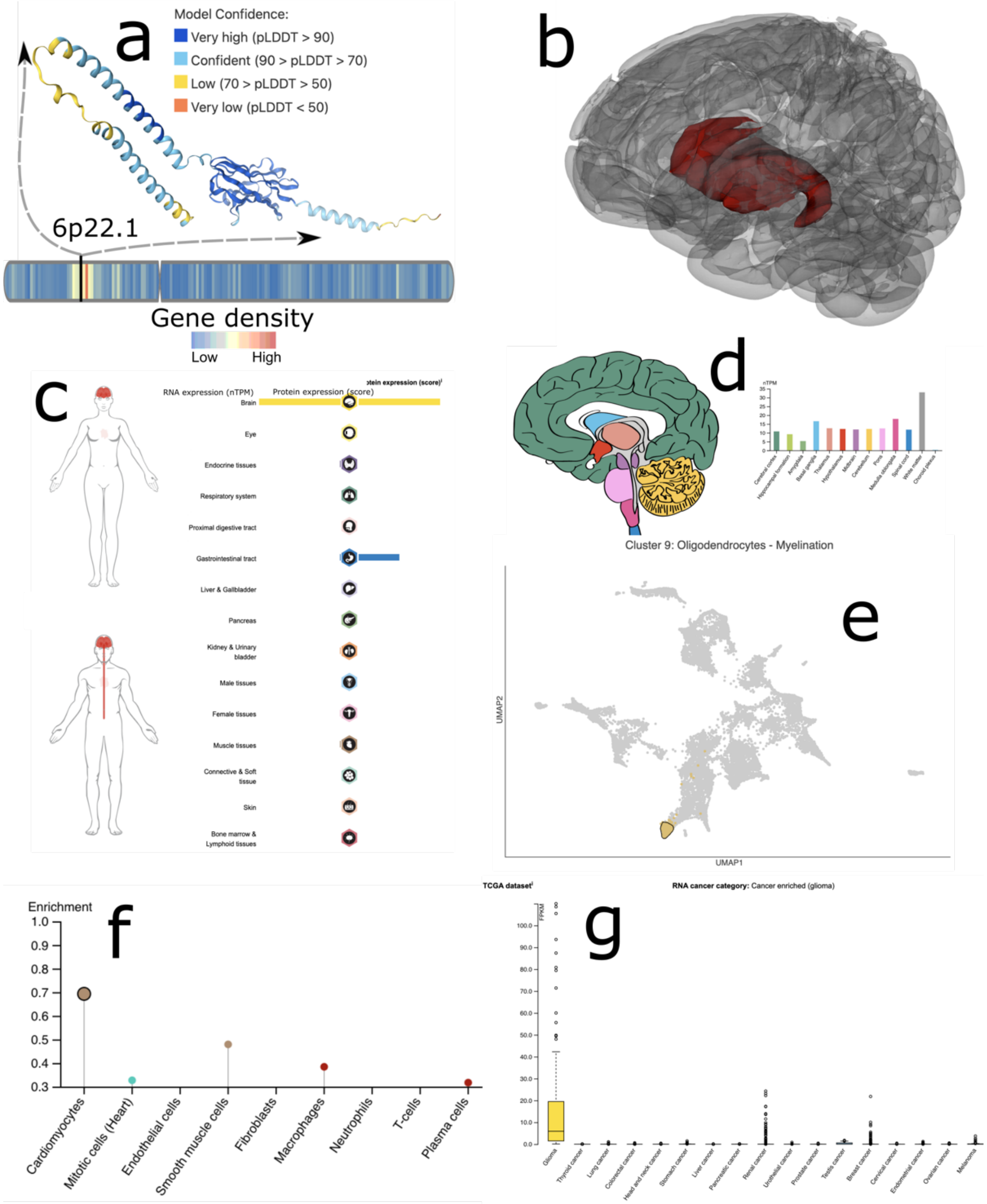


(**a**) Protein structure. (**b**) The brain PSC visualization for C32_1. (**c**) The summary of expression level of the protein in whole-body tissues using RNA and protein data. (**d**) Brain-tissue specificity of the protein expression. (**e**) The visualization of the protein cluster 9: Oligodendrocytes – Myelination. (**f**) Protein cell type-specific enrichment. (**g**) Protein cancer type-specific enrichment.

**eFigure 28: The expression analyses of the PLTP protein in the HPA**

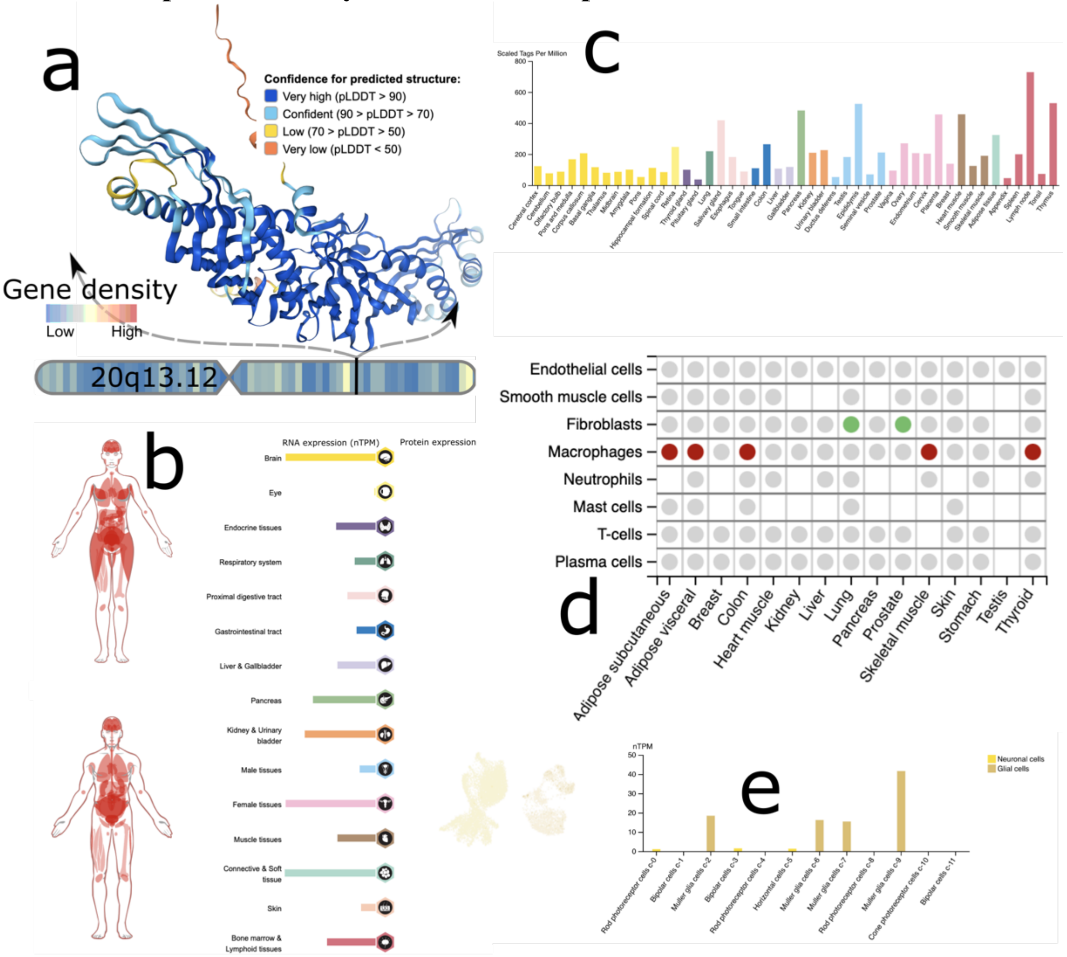


(**a**) Protein structure. (**b**) The summary of expression level of the protein in whole-body tissues using RNA and protein data. (**c**) Detailed expression level of the protein in different tissues/organs. (**d**) Protein tissue cell-type-specific enrichment. (**e**) Protein single-cell tissue-type-specific enrichment.

1047 **eFigure 29: The expression analyses of the TGFA protein in the HPA**

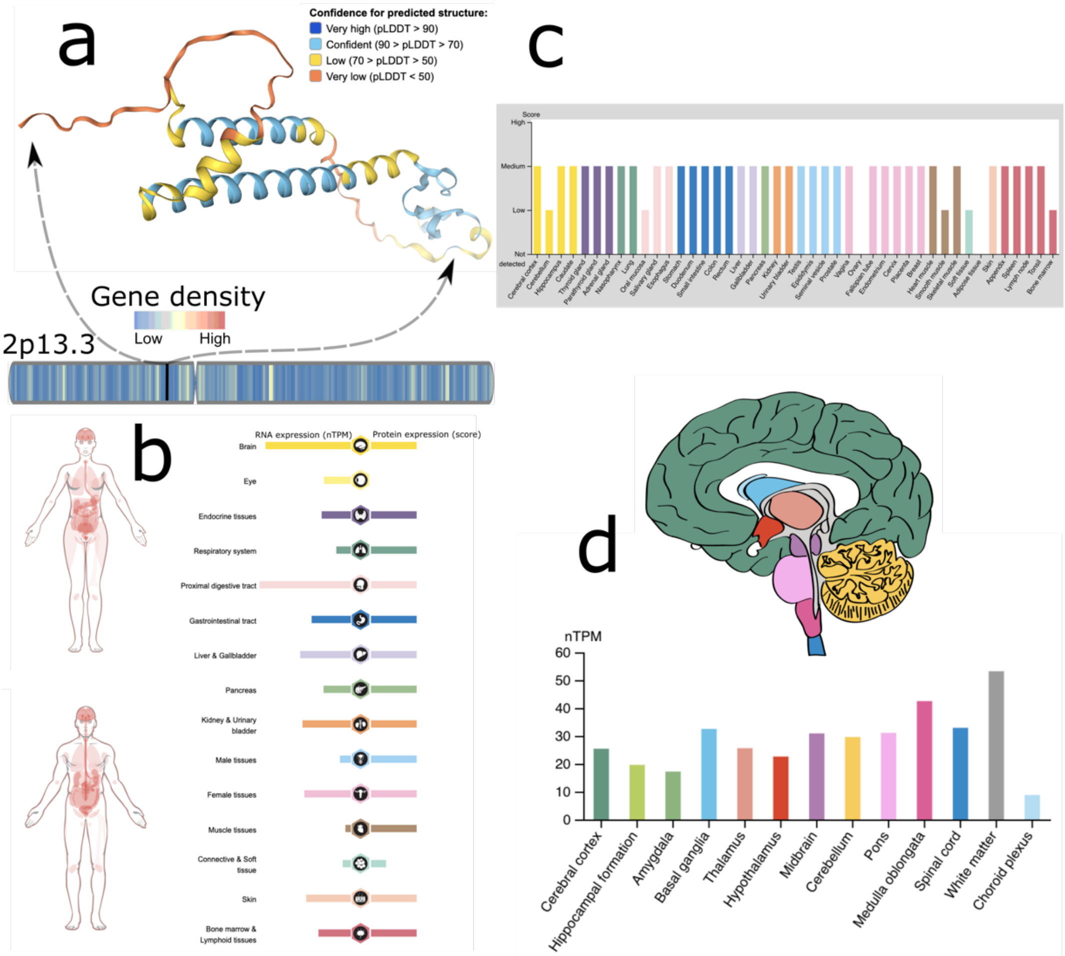


1048

1049 (**a**) Protein structure. (**b**) The summary of expression level of the protein in whole-body tissues
1050 using RNA and protein data. (**c**) Detailed expression level of the protein in different
1051 tissues/organs. (**d**) Brain-tissue specificity of the protein expression.

1052

**eFigure 30**: **Phenotypic correlations between the 119 MUSE ROIs and 128 brain PSCs at the scale of C=128**.

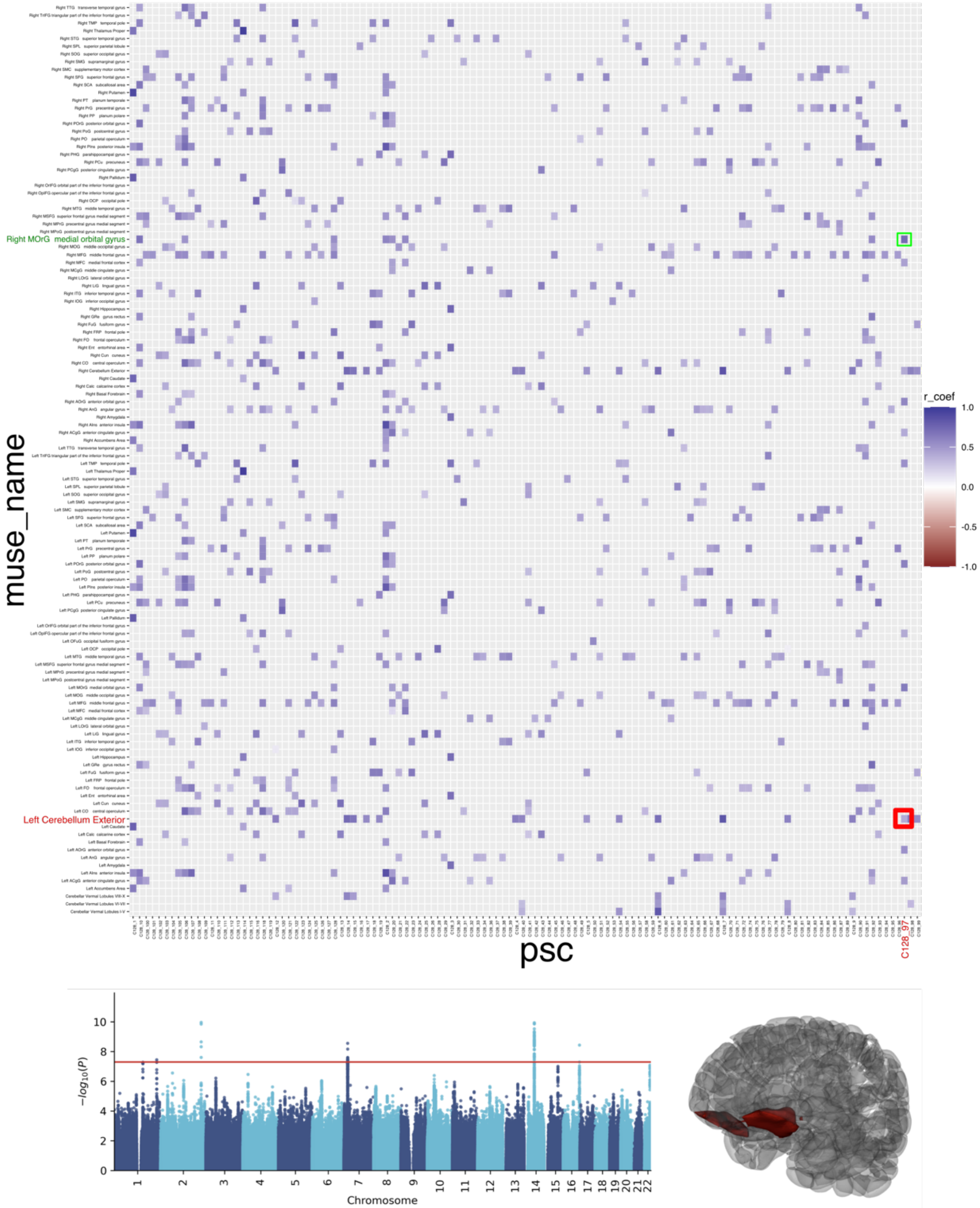


We highlighted one example (red rectangle) of a positive PSC-MUSE association, demonstrated by the phenotypic correlation between C128_97 (https://labs-laboratory.com/bridgeport/music/C128_97, which includes the frontal poles) and the MUSE ROI (left cerebellum exterior). Another example is to highlight the other MUSE ROI (middle orbital gyrus) that spatially overlaps with C128_97 (green rectangle).

**eFigure 31**: **Genetic correlations between the 119 MUSE ROIs and 128 brain PSCs at the scale of C=128**.

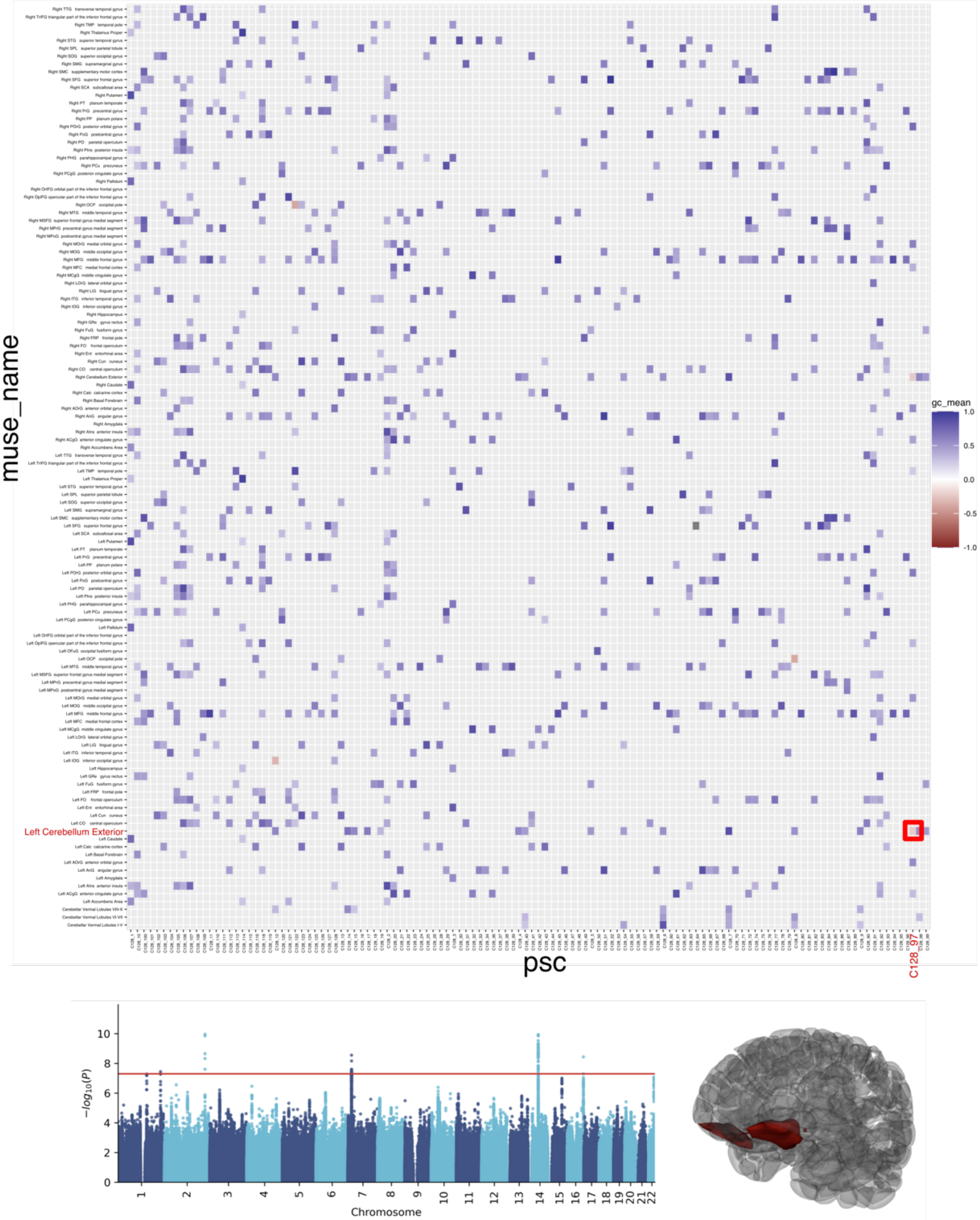


We highlighted one example of a negative PSC-MUSE association, demonstrated by the phenotypic correlation between C128_97 (https://labs-laboratory.com/bridgeport/music/C128_97, which includes the frontal poles) and the MUSE ROI (left cerebellum exterior).

**eFigure 32**: **Spatially overlap between C128_97 and the right orbital gyrus from the MUSE atlas**.

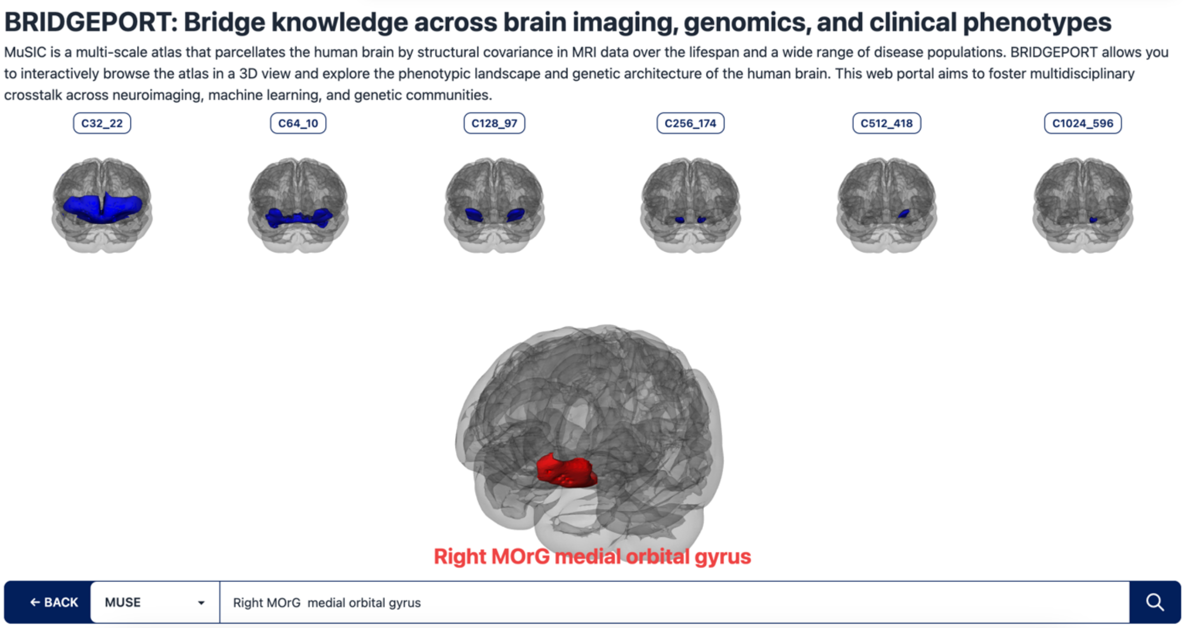


This screenshot demonstrates the functionality of our online BRIDGEPORT portal, which maps our brain PSCs to the conventional MUSE atlas ROIs based on their spatial overlaps. The original link is here: https://labs-laboratory.com/bridgeport/music/right_medial_orbital_gyrus.

**eFigure 33**: **Genetic correlation between the 2003 brain PSCs and ~4000 brain IDPs from Smith et al**.

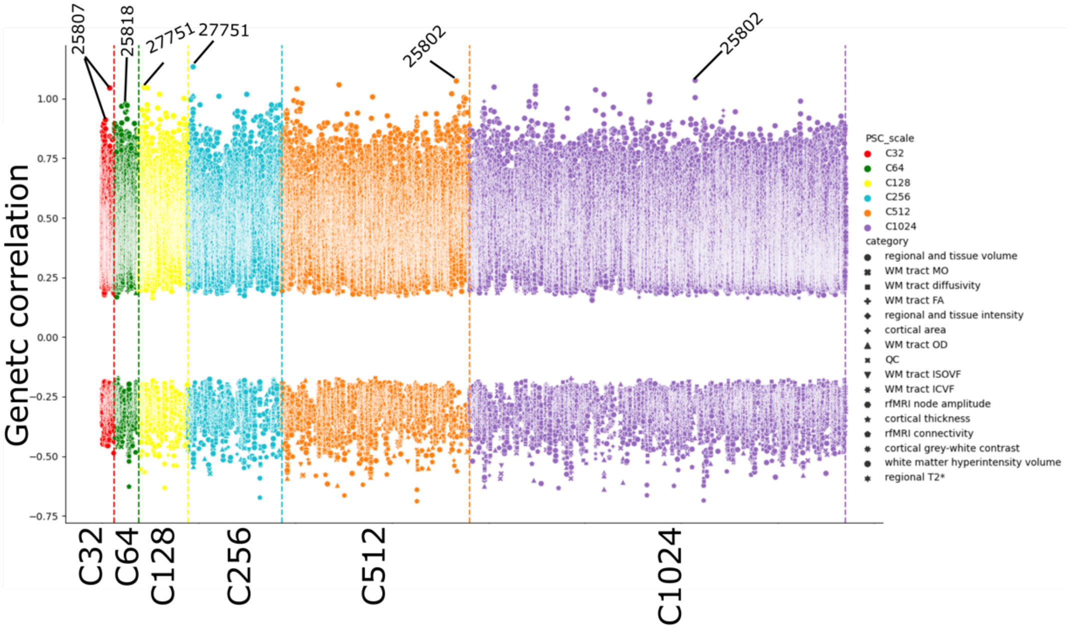


At each level of the brain PSCs, we report significant associations between our brain PSCs and the 3936 conventional multi-modal brain IDPs from the BIG-40 portal. These 3936 brain IDPs are categorized into different high-level features based on imaging modality. As expected, our GM T1-weighted-derived brain PSCs were primarily associated with regional and tissue volume IDPs derived from T1-weighted brain MRI (indicated by round dots in the figure). We have annotated the most prominent associations at each scale with searchable Field IDs from the UKBB website: https://biobank.ndph.ox.ac.uk/showcase/search.cgi. The anatomical correspondence was further discussed in **Supplementary eText 2**.

**eFigure 34: The correlation of the incremental $R^2$ of the heart IDP PRS between the methods and studies**

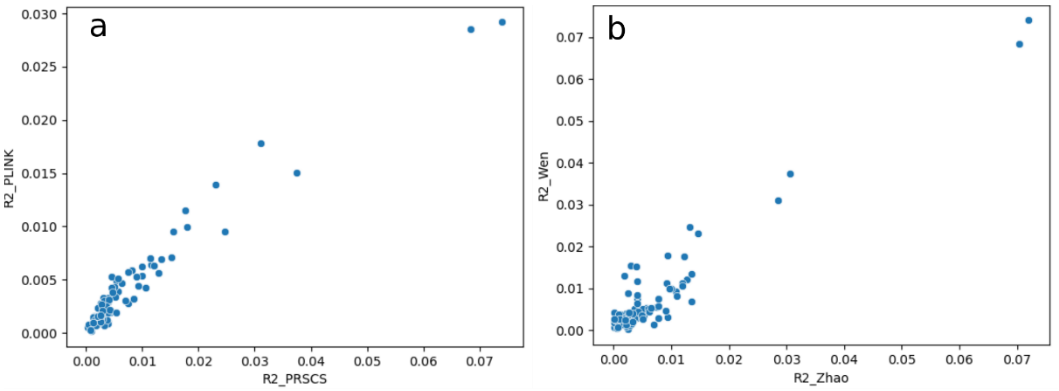


**a)** The PRS calculation (incremental $R^2$) from the PLINK and PRS-CS were highly correlated. **b**) For the 82 heart IDPs between our GWAS and the GWAS conducted by Zhao et al.[4], the two sets of PRS calculations were highly correlated.

**eFigure 35: ProWAS coefficients between the two sets of beta values of the brain ProWAS in the UKBB and the BLSA studies.**

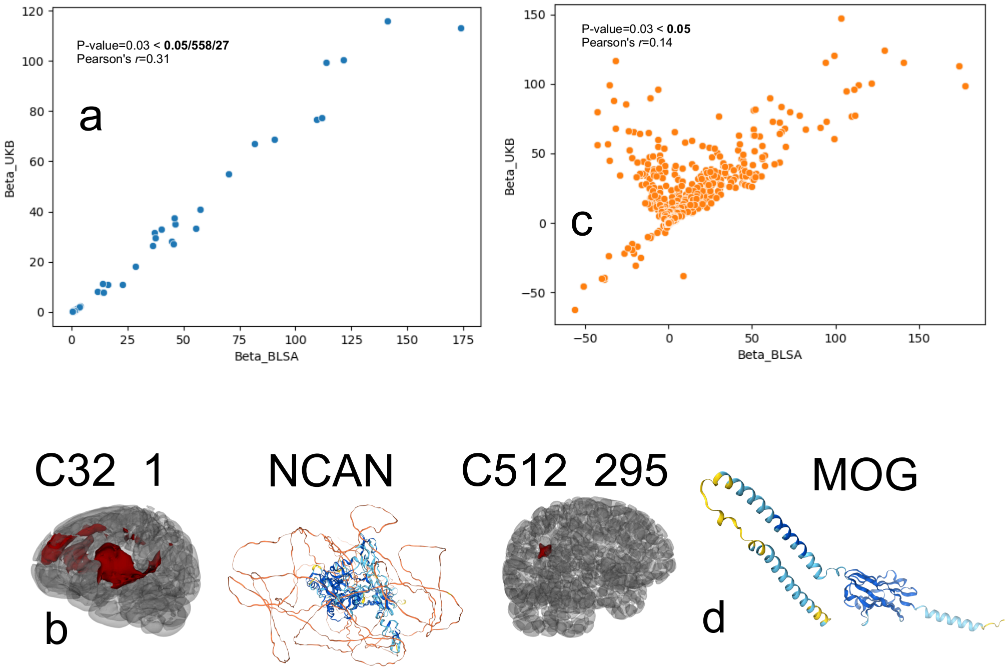


We replicated the significant PSC-protein pairs identified in the UKBB using the BLSA data. For the common PSC-protein pairs that passed the Bonferroni correction, we computed the Pearson's *r* coefficient: **a**) replication at a stringent P-value threshold (0.05/558/27), and **c**) replication at a nominal P-value threshold (0.05). **b**) and **d**) display the visualization of the example PSC-protein associations for **a**) and **c**).

**eTable 1: The 2003 brain PSCs, 82 heart IDPs, and 84 eye IDPs**

a) **Brain PSCs**: The 2003 brain PSCs originated from our prior investigation utilizing six scales of the MuSIC atlas[15] (C=32, 64, 128, 256, 512, and 1024, with 13 PSCs at C1024 omitted during optimization).

| Scale (C) | Number of PSCs | Name | N |
|---|---|---|---|
| 32 | 32 | C32_*N* | 39,567 |
| 64 | 64 | C64_*N* | 39,567 |
| 128 | 128 | C128_*N* | 39,567 |
| 256 | 256 | C256_*N* | 39,567 |
| 512 | 512 | C512_*N* | 39,567 |
| 1024 | 1011 | C1024_*N* | 39,567 |

b) **Heart IDPs**: The 82 heart IDPs were downloaded directly from UKBB. We present the full name of the heart IDPs, Field ID on the UKBB showcase website, abbreviations, high-level groups.

| IDP | Abb | Group | Field ID | Heart field | N |
|---|---|---|---|---|---|
| lv_end_diastolic_volume_f24100_2_0 | LVEDV | LV | 24100 | left_ventricle | 39286 |
| lv_end_systolic_volume_f24101_2_0 | LVESV | LV | 24101 | left_ventricle | 39286 |
| lv_stroke_volume_f24102_2_0 | LVSV | LV | 24102 | left_ventricle | 39286 |
| lv_ejection_fraction_f24103_2_0 | LVEF | LV | 24103 | left_ventricle | 39286 |
| lv_cardiac_output_f24104_2_0 | LVCO | LV | 24104 | left_ventricle | 39286 |
| lv_myocardial_mass_f24105_2_0 | LVM | LV | 24105 | left_ventricle | 39286 |
| rv_end_diastolic_volume_f24106_2_0 | RVEDV | RV | 24106 | right_ventricle | 39286 |
| rv_end_systolic_volume_f24107_2_0 | RVESV | RV | 24107 | right_ventricle | 39286 |
| rv_stroke_volume_f24108_2_0 | RVSV | RV | 24108 | right_ventricle | 39286 |
| rv_ejection_fraction_f24109_2_0 | RVEF | RV | 24109 | right_ventricle | 39286 |
| la_maximum_volume_f24110_2_0 | LAV_max | LA | 24110 | left_atrium | 38863 |
| la_minimum_volume_f24111_2_0 | LAV_min | LA | 24111 | left_atrium | 38863 |
| la_stroke_volume_f24112_2_0 | LASV | LA | 24112 | left_atrium | 38863 |
| la_ejection_fraction_f24113_2_0 | LAEF | LA | 24113 | left_atrium | 38863 |
| ra_maximum_volume_f24114_2_0 | RAV_max | RA | 24114 | left_atrium | 38863 |
| ra_minimum_volume_f24115_2_0 | RAV_min | RA | 24115 | left_atrium | 38863 |
| ra_stroke_volume_f24116_2_0 | RASV | RA | 24116 | left_atrium | 38863 |
| ra_ejection_fraction_f24117_2_0 | RAEF | RA | 24117 | left_atrium | 38863 |
| ascending_aorta_maximum_area_f24118_2_0 | Aao_max | AA | 24118 | ascending_aorta | 37097 |
| ascending_aorta_minimum_area_f24119_2_0 | Aao_min | AA | 24119 | ascending_aorta | 37097 |
| ascending_aorta_distensibility_f24120_2_0 | Aao_AD | AA | 24120 | ascending_aorta | 33866 |

| | | | | | |
|---|---|---|---|---|---|
| descending_aorta_maximum_area_f24121_2_0 | Dao_max | DA | 24121 | descending_aorta | 37097 |
| descending_aorta_minimum_area_f24122_2_0 | Dao_min | DA | 24122 | descending_aorta | 37097 |
| descending_aorta_distensibility_f24123_2_0 | Dao_AD | DA | 24123 | descending_aorta | 33866 |
| lv_mean_myocardial_wall_thickness_aha_1_f24124_2_0 | WT_AHA_1 | LV | 24124 | left_ventricle | 39241 |
| lv_mean_myocardial_wall_thickness_aha_2_f24125_2_0 | WT_AHA_2 | LV | 24125 | left_ventricle | 39241 |
| lv_mean_myocardial_wall_thickness_aha_3_f24126_2_0 | WT_AHA_3 | LV | 24126 | left_ventricle | 39241 |
| lv_mean_myocardial_wall_thickness_aha_4_f24127_2_0 | WT_AHA_4 | LV | 24127 | left_ventricle | 39241 |
| lv_mean_myocardial_wall_thickness_aha_5_f24128_2_0 | WT_AHA_5 | LV | 24128 | left_ventricle | 39241 |
| lv_mean_myocardial_wall_thickness_aha_6_f24129_2_0 | WT_AHA_6 | LV | 24129 | left_ventricle | 39241 |
| lv_mean_myocardial_wall_thickness_aha_7_f24130_2_0 | WT_AHA_7 | LV | 24130 | left_ventricle | 39241 |
| lv_mean_myocardial_wall_thickness_aha_8_f24131_2_0 | WT_AHA_8 | LV | 24131 | left_ventricle | 39241 |
| lv_mean_myocardial_wall_thickness_aha_9_f24132_2_0 | WT_AHA_9 | LV | 24132 | left_ventricle | 39241 |
| lv_mean_myocardial_wall_thickness_aha_10_f24133_2_0 | WT_AHA_10 | LV | 24133 | left_ventricle | 39241 |
| lv_mean_myocardial_wall_thickness_aha_11_f24134_2_0 | WT_AHA_11 | LV | 24134 | left_ventricle | 39241 |
| lv_mean_myocardial_wall_thickness_aha_12_f24135_2_0 | WT_AHA_12 | LV | 24135 | left_ventricle | 39241 |
| lv_mean_myocardial_wall_thickness_aha_13_f24136_2_0 | WT_AHA_13 | LV | 24136 | left_ventricle | 39241 |
| lv_mean_myocardial_wall_thickness_aha_14_f24137_2_0 | WT_AHA_14 | LV | 24137 | left_ventricle | 39241 |
| lv_mean_myocardial_wall_thickness_aha_15_f24138_2_0 | WT_AHA_15 | LV | 24138 | left_ventricle | 39241 |
| lv_mean_myocardial_wall_thickness_aha_16_f24139_2_0 | WT_AHA_16 | LV | 24139 | left_ventricle | 39241 |
| lv_mean_myocardial_wall_thickness_global_f24140_2_0 | WT_global | LV | 24140 | left_ventricle | 39241 |
| lv_circumferential_strain_aha_1_f24141_2_0 | Ecc_AHA_1 | LV | 24141 | left_ventricle | 39217 |
| lv_circumferential_strain_aha_2_f24142_2_0 | Ecc_AHA_2 | LV | 24142 | left_ventricle | 39217 |
| lv_circumferential_strain_aha_3_f24143_2_0 | Ecc_AHA_3 | LV | 24143 | left_ventricle | 39217 |
| lv_circumferential_strain_aha_4_f24144_2_0 | Ecc_AHA_4 | LV | 24144 | left_ventricle | 39217 |
| lv_circumferential_strain_aha_5_f24145_2_0 | Ecc_AHA_5 | LV | 24145 | left_ventricle | 39217 |
| lv_circumferential_strain_aha_6_f24146_2_0 | Ecc_AHA_6 | LV | 24146 | left_ventricle | 39217 |
| lv_circumferential_strain_aha_7_f24147_2_0 | Ecc_AHA_7 | LV | 24147 | left_ventricle | 39217 |
| lv_circumferential_strain_aha_8_f24148_2_0 | Ecc_AHA_8 | LV | 24148 | left_ventricle | 39217 |
| lv_circumferential_strain_aha_9_f24149_2_0 | Ecc_AHA_9 | LV | 24149 | left_ventricle | 39217 |
| lv_circumferential_strain_aha_10_f24150_2_0 | Ecc_AHA_10 | LV | 24150 | left_ventricle | 39217 |
| lv_circumferential_strain_aha_11_f24151_2_0 | Ecc_AHA_11 | LV | 24151 | left_ventricle | 39217 |
| lv_circumferential_strain_aha_12_f24152_2_0 | Ecc_AHA_12 | LV | 24152 | left_ventricle | 39217 |
| lv_circumferential_strain_aha_13_f24153_2_0 | Ecc_AHA_13 | LV | 24153 | left_ventricle | 39217 |
| lv_circumferential_strain_aha_14_f24154_2_0 | Ecc_AHA_14 | LV | 24154 | left_ventricle | 39217 |
| lv_circumferential_strain_aha_15_f24155_2_0 | Ecc_AHA_15 | LV | 24155 | left_ventricle | 39217 |

| | | | | | |
|---|---|---|---|---|---|
| lv_circumferential_strain_aha_16_f24156_2_0 | Ecc_AHA_16 | LV | 24156 | left_ventricle | 39217 |
| lv_circumferential_strain_global_f24157_2_0 | Ecc_global | LV | 24157 | left_ventricle | 39217 |
| lv_radial_strain_aha_1_f24158_2_0 | Err_AHA_1 | LV | 24158 | left_ventricle | 39217 |
| lv_radial_strain_aha_2_f24159_2_0 | Err_AHA_2 | LV | 24159 | left_ventricle | 39217 |
| lv_radial_strain_aha_3_f24160_2_0 | Err_AHA_3 | LV | 24160 | left_ventricle | 39217 |
| lv_radial_strain_aha_4_f24161_2_0 | Err_AHA_4 | LV | 24161 | left_ventricle | 39217 |
| lv_radial_strain_aha_5_f24162_2_0 | Err_AHA_5 | LV | 24162 | left_ventricle | 39217 |
| lv_radial_strain_aha_6_f24163_2_0 | Err_AHA_6 | LV | 24163 | left_ventricle | 39217 |
| lv_radial_strain_aha_7_f24164_2_0 | Err_AHA_7 | LV | 24164 | left_ventricle | 39217 |
| lv_radial_strain_aha_8_f24165_2_0 | Err_AHA_8 | LV | 24165 | left_ventricle | 39217 |
| lv_radial_strain_aha_9_f24166_2_0 | Err_AHA_9 | LV | 24166 | left_ventricle | 39217 |
| lv_radial_strain_aha_10_f24167_2_0 | Err_AHA_10 | LV | 24167 | left_ventricle | 39217 |
| lv_radial_strain_aha_11_f24168_2_0 | Err_AHA_11 | LV | 24168 | left_ventricle | 39217 |
| lv_radial_strain_aha_12_f24169_2_0 | Err_AHA_12 | LV | 24169 | left_ventricle | 39217 |
| lv_radial_strain_aha_13_f24170_2_0 | Err_AHA_13 | LV | 24170 | left_ventricle | 39217 |
| lv_radial_strain_aha_14_f24171_2_0 | Err_AHA_14 | LV | 24171 | left_ventricle | 39217 |
| lv_radial_strain_aha_15_f24172_2_0 | Err_AHA_15 | LV | 24172 | left_ventricle | 39217 |
| lv_radial_strain_aha_16_f24173_2_0 | Err_AHA_16 | LV | 24173 | left_ventricle | 39217 |
| lv_radial_strain_global_f24174_2_0 | Err_global | LV | 24174 | left_ventricle | 39217 |
| lv_longitudinal_strain_segment_1_f24175_2_0 | Ell_1 | LV | 24175 | left_ventricle | 38134 |
| lv_longitudinal_strain_segment_2_f24176_2_0 | Ell_2 | LV | 24176 | left_ventricle | 38134 |
| lv_longitudinal_strain_segment_3_f24177_2_0 | Ell_3 | LV | 24177 | left_ventricle | 38140 |
| lv_longitudinal_strain_segment_4_f24178_2_0 | Ell_4 | LV | 24178 | left_ventricle | 38140 |
| lv_longitudinal_strain_segment_5_f24179_2_0 | Ell_5 | LV | 24179 | left_ventricle | 38139 |
| lv_longitudinal_strain_segment_6_f24180_2_0 | Ell_6 | LV | 24180 | left_ventricle | 38139 |
| lv_longitudinal_strain_global_f24181_2_0 | Ell_global | LV | 24181 | left_ventricle | 38141 |

c) **Eye IDPs**: The 84 eye IDPs were downloaded directly from UKBB. We present the full name of the eye IDPs, Field ID on the UKBB showcase website, abbreviations, high-level groups, and PubMed ID.

| **Eye IDP** | **Field ID** | **Abb** | **Group** | **Pubmed** | **N** |
|---|---|---|---|---|---|
| overall_macular_thickness_left_f27800_0_0 | 27800 | ML | MT | 26746598 | 60335 |
| overall_macular_thickness_right_f27801_0_0 | 27801 | MR | MT | 26746598 | 59823 |
| macular_thickness_at_the_central_subfield_left_f27802_0_0 | 27802 | McL | MT | 26746598 | 61404 |
| macular_thickness_at_the_central_subfield_right_f27803_0_0 | 27803 | McR | MT | 26746598 | 60457 |

| | | | | | |
|---|---|---|---|---|---|
| macular_thickness_at_the_inner_inferior_subfield_left_f27804_0_0 | 27804 | MiiL | MT | 26746598 | 60681 |
| macular_thickness_at_the_inner_inferior_subfield_right_f27805_0_0 | 27805 | MiiR | MT | 26746598 | 60157 |
| macular_thickness_at_the_inner_nasal_subfield_left_f27806_0_0 | 27806 | MinL | MT | 26746598 | 61351 |
| macular_thickness_at_the_inner_nasal_subfield_right_f27807_0_0 | 27807 | MinR | MT | 26746598 | 60456 |
| macular_thickness_at_the_inner_superior_subfield_left_f27808_0_0 | 27808 | MisL | MT | 26746598 | 61404 |
| macular_thickness_at_the_inner_superior_subfield_right_f27809_0_0 | 27809 | MisR | MT | 26746598 | 60457 |
| macular_thickness_at_the_inner_temporal_subfield_left_f27810_0_0 | 27810 | MitL | MT | 26746598 | 61404 |
| macular_thickness_at_the_inner_temporal_subfield_right_f27811_0_0 | 27811 | MitR | MT | 26746598 | 60407 |
| macular_thickness_at_the_outer_inferior_subfield_left_f27812_0_0 | 27812 | MoiL | MT | 26746598 | 60500 |
| macular_thickness_at_the_outer_inferior_subfield_right_f27813_0_0 | 27813 | MoiR | MT | 26746598 | 59989 |
| macular_thickness_at_the_outer_nasal_subfield_left_f27814_0_0 | 27814 | MonL | MT | 26746598 | 61259 |
| macular_thickness_at_the_outer_nasal_subfield_right_f27815_0_0 | 27815 | MonR | MT | 26746598 | 60232 |
| macular_thickness_at_the_outer_superior_subfield_left_f27816_0_0 | 27816 | MosL | MT | 26746598 | 61202 |
| macular_thickness_at_the_outer_superior_subfield_right_f27817_0_0 | 27817 | MosR | MT | 26746598 | 60231 |
| macular_thickness_at_the_outer_temporal_subfield_left_f27818_0_0 | 27818 | MotL | MT | 26746598 | 61185 |
| macular_thickness_at_the_outer_temporal_subfield_right_f27819_0_0 | 27819 | MotR | MT | 26746598 | 60330 |
| average_retinal_nerve_fibre_layer_thickness_left_f28500_0_0 | 28500 | RnfL | RNF | 26746598 | 60335 |
| average_retinal_nerve_fibre_layer_thickness_right_f28501_0_0 | 28501 | RnfR | RNF | 26746598 | 59823 |
| average_inner_nuclear_layer_thickness_left_f28502_0_0 | 28502 | InlL | INL | 26746598 | 60335 |
| average_inner_nuclear_layer_thickness_right_f28503_0_0 | 28503 | InlR | INL | 26746598 | 59823 |
| average_ganglion_cellinner_plexiform_layer_thickness_left_f28504_0_0 | 28504 | GcplL | GCPL | 26746598 | 60335 |
| average_ganglion_cellinner_plexiform_layer_thickness_right_f28505_0_0 | 28505 | GcplR | GCPL | 26746598 | 59823 |
| inlelm_thickness_of_the_central_subfield_left_f28506_0_0 | 28506 | InlElmCL | INL/ELM | 26746598 | 61732 |
| inlelm_thickness_of_the_central_subfield_right_f28507_0_0 | 28507 | InlElmCR | INL/ELM | 26746598 | 60777 |
| inlelm_thickness_of_the_inner_subfield_left_f28508_0_0 | 28508 | InlElmIL | INL/ELM | 26746598 | 60998 |
| inlelm_thickness_of_the_inner_subfield_right_f28509_0_0 | 28509 | InlElmIR | INL/ELM | 26746598 | 60475 |
| inlelm_thickness_of_the_outer_subfield_left_f28510_0_0 | 28510 | InlElmOL | INL/ELM | 26746598 | 60335 |
| inlelm_thickness_of_the_outer_subfield_right_f28511_0_0 | 28511 | InlElmOR | INL/ELM | 26746598 | 59823 |
| average_inlelm_thickness_left_f28512_0_0 | 28512 | InlElmL | INL/ELM | 26746598 | 60335 |
| average_inlelm_thickness_right_f28513_0_0 | 28513 | InlElmR | INL/ELM | 26746598 | 59823 |
| elmisos_thickness_of_central_subfield_left_f28514_0_0 | 28514 | ElmIsosCL | ELM/ISOS | 26746598 | 61732 |
| elmisos_thickness_of_central_subfield_right_f28515_0_0 | 28515 | ElmIsosCR | ELM/ISOS | 26746598 | 60777 |
| elmisos_thickness_of_inner_subfield_left_f28516_0_0 | 28516 | ElmIsosIL | ELM/ISOS | 26746598 | 60998 |
| elmisos_thickness_of_inner_subfield_right_f28517_0_0 | 28517 | ElmIsosIR | ELM/ISOS | 26746598 | 60475 |
| elmisos_thickness_of_outer_subfield_left_f28518_0_0 | 28518 | ElmIsosOL | ELM/ISOS | 26746598 | 60335 |

| | | | | | |
|---|---|---|---|---|---|
| elmisos_thickness_of_outer_subfield_right_f28519_0_0 | 28519 | ElmIsosOR | ELM/ISOS | 26746598 | 59823 |
| average_elmisos_thickness_left_f28520_0_0 | 28520 | ElmIsosL | ELM/ISOS | 26746598 | 60335 |
| average_elmisos_thickness_right_f28521_0_0 | 28521 | ElmIsosR | ELM/ISOS | 26746598 | 59823 |
| isosrpe_thickness_of_central_subfield_left_f28522_0_0 | 28522 | IsosRpeCL | ISOS/RPE | 26746598 | 61732 |
| isosrpe_thickness_of_central_subfield_right_f28523_0_0 | 28523 | IsosRpeCR | ISOS/RPE | 26746598 | 60777 |
| isosrpe_thickness_of_inner_subfield_left_f28524_0_0 | 28524 | IsosRpeIL | ISOS/RPE | 26746598 | 60998 |
| isosrpe_thickness_of_inner_subfield_right_f28525_0_0 | 28525 | IsosRpeIR | ISOS/RPE | 26746598 | 60475 |
| isosrpe_thickness_of_outer_subfield_left_f28526_0_0 | 28526 | IsosRpeOL | ISOS/RPE | 26746598 | 60335 |
| isosrpe_thickness_of_outer_subfield_right_f28527_0_0 | 28527 | IsosRpeOR | ISOS/RPE | 26746598 | 59823 |
| average_isosrpe_thickness_left_f28528_0_0 | 28528 | IsosRpeL | ISOS/RPE | 26746598 | 60335 |
| average_isosrpe_thickness_right_f28529_0_0 | 28529 | IsosRpeR | ISOS/RPE | 26746598 | 59823 |
| inlrpe_thickness_of_central_subfield_left_f28530_0_0 | 28530 | InlRpeCL | INL/RPE | 26746598 | 61732 |
| inlrpe_thickness_of_central_subfield_right_f28531_0_0 | 28531 | InlRpeCR | INL/RPE | 26746598 | 60777 |
| inlrpe_thickness_of_inner_subfield_left_f28532_0_0 | 28532 | InlRpeIL | INL/RPE | 26746598 | 60998 |
| inlrpe_thickness_of_inner_subfield_right_f28533_0_0 | 28533 | InlRpeIR | INL/RPE | 26746598 | 60475 |
| inlrpe_thickness_of_outer_subfield_left_f28534_0_0 | 28534 | InlRpeOL | INL/RPE | 26746598 | 60335 |
| inlrpe_thickness_of_outer_subfield_right_f28535_0_0 | 28535 | InlRpeOR | INL/RPE | 26746598 | 59823 |
| average_inlrpe_thickness_left_f28536_0_0 | 28536 | InlRpeL | INL/RPE | 26746598 | 60335 |
| average_inlrpe_thickness_right_f28537_0_0 | 28537 | InlRpeR | INL/RPE | 26746598 | 59823 |
| overall_average_retinal_pigment_epithelium_thickness_left_f27822_0_0 | 27822 | RpeL | RPE | 26746598 | 60335 |
| overall_average_retinal_pigment_epithelium_thickness_right_f27823_0_0 | 27823 | RpeR | RPE | 26746598 | 59823 |
| retinal_pigment_epithelium_thickness_at_central_subfield_left_f27824_0_0 | 27824 | RpeCL | RPE | 26746598 | 17408 |
| retinal_pigment_epithelium_thickness_at_central_subfield_right_f27825_0_0 | 27825 | RpeCR | RPE | 26746598 | 17038 |
| retinal_pigment_epithelium_thickness_at_inner_inferior_subfield_left_f27826_0_0 | 27826 | RpeIiL | RPE | 26746598 | 17408 |
| retinal_pigment_epithelium_thickness_at_inner_inferior_subfield_right_f27827_0_0 | 27827 | RpeIiR | RPE | 26746598 | 17038 |
| retinal_pigment_epithelium_thickness_at_inner_nasal_subfield_left_f27828_0_0 | 27828 | RpeInL | RPE | 26746598 | 17408 |
| retinal_pigment_epithelium_thickness_at_inner_nasal_subfield_right_f27829_0_0 | 27829 | RpeInR | RPE | 26746598 | 17038 |
| retinal_pigment_epithelium_thickness_at_inner_superior_subfield_left_f27830_0_0 | 27830 | RpeIsL | RPE | 26746598 | 17408 |
| retinal_pigment_epithelium_thickness_at_inner_superior_subfield_right_f27831_0_0 | 27831 | RpeIsR | RPE | 26746598 | 17038 |
| retinal_pigment_epithelium_thickness_at_inner_temporal_subfield_left_f27832_0_0 | 27832 | RpeItL | RPE | 26746598 | 17408 |
| retinal_pigment_epithelium_thickness_at_inner_temporal_subfield_right_f27833_0_0 | 27833 | RpeItR | RPE | 26746598 | 17038 |
| retinal_pigment_epithelium_thickness_at_outer_inferior_subfield_left_f27834_0_0 | 27834 | RpeOiL | RPE | 26746598 | 17408 |
| retinal_pigment_epithelium_thickness_at_outer_inferior_subfield_right_f27835_0_0 | 27835 | RpeOiR | RPE | 26746598 | 17038 |
| retinal_pigment_epithelium_thickness_at_outer_nasal_subfield_left_f27836_0_0 | 27836 | RpeOnL | RPE | 26746598 | 17408 |
| retinal_pigment_epithelium_thickness_at_outer_nasal_subfield_right_f27837_0_0 | 27837 | RpeOnR | RPE | 26746598 | 17038 |

| | | | | | |
|---|---|---|---|---|---|
| retinal_pigment_epithelium_thickness_at_outer_superior_subfield_left_f27838_0_0 | 27838 | RpeOsL | RPE | 26746598 | 17408 |
| retinal_pigment_epithelium_thickness_at_outer_superior_subfield_right_f27839_0_0 | 27839 | RpeOsR | RPE | 26746598 | 17038 |
| retinal_pigment_epithelium_thickness_at_outer_temporal_subfield_left_f27840_0_0 | 27840 | RpeOtL | RPE | 26746598 | 17408 |
| retinal_pigment_epithelium_thickness_at_outer_temporal_subfield_right_f27841_0_0 | 27841 | RpeOtR | RPE | 26746598 | 17038 |
| disc_diameter_after_inverse_rank_normal_transformation_left_f27851_0_0 | 27851 | DdL | DISC | 31809533 | 45522 |
| mean_of_vertical_disc_diameter_left_f27853_0_0 | 27853 | VdiscL | DISC | 31809533 | 45522 |
| vertical_cup_to_disc_ratio_vcdr_regressed_and_transformed_left_f27855_0_0 | 27855 | VcdrTL | DISC | 31809533 | 44171 |
| vertical_cup_to_disc_ratio_vcdr_left_f27857_0_0 | 27857 | VcdrL | DISC | 31809533 | 44171 |
| total_macular_volume_left_f27820_0_0 | 27820 | MvL | MV | 26746598 | 16049 |
| total_macular_volume_right_f27821_0_0 | 27821 | MvR | MV | 26746598 | 15997 |

**eTable 2: The 14 systemic disease categories and 8 cognitive scores**

**a) 14 systemic disease categories:**

| Disease | ICD-10 |
|---|---|
| infectious_parasitic_disease_diagnosis | B |
| neoplasms_diagnosis | C-D4 |
| blood_and_immune_system_diagnosis | D50-D89 |
| endocrine_nutritional_metabolic_disease_diagnosis | E |
| mental_behavioural_disorder_diagnosis | F |
| nerve_system_diagnosis | G |
| eye_diagnosis | H0-H5 |
| ear_diagnosis | H6-H9 |
| circular_system_diagnosis | I |
| respiratory_system_diagnosis | J |
| digestive_system_diagnosis | K |
| skin_system_diagnosis | L |
| musculoskeletal_system_diagnosis | M |
| genitourinary_system_diagnosis | N |

**b) 8 cognitive scores:**

| Cognition | Filed_ID |
|---|---|
| number_of_puzzles_correct_f21004_2_0 | 21004 |
| number_of_symbol_digit_matches_made_correctly_f23324_2_0 | 23324 |
| number_of_puzzles_correctly_solved_f6373_2_0 | 6373 |
| duration_to_complete_numeric_path_trail_1_f6348_2_0 | 6348 |
| duration_to_complete_alphanumeric_path_trail_2_f6350_2_0 | 6350 |
| fluid_intelligence_score_f20016_2_0 | 20016 |
| maximum_digits_remembered_correctly_f4282_2_0 | 4282 |
| mean_time_to_correctly_identify_matches_f20023_0_0 | 20023 |

**eFile 1-64: The Online Supplementary files contain large tables**

Due to the large size, we share these online data files via Google Drive during peer review. Reviewers and editors can use the following links to download the data: https://drive.google.com/drive/folders/1_piv-9hrtLWv34Ja9DzMBftKzoif8NAS?usp=sharing.
